# Actinic keratosis labeling and single-band instability in near-infrared spectroscopy of skin cancer: a nested, patient-level re-analysis

**DOI:** 10.64898/2026.10.25.26366432

**Authors:** Seyedeh Rahemeh Amiri, Ali Khoshnami

**Affiliations:** University at Albany, State University of New York (SUNY), Albany, NY, USA; Shiraz University, Shiraz, Iran

**Author notes:** Corresponding author: Seyedeh Rahemeh Amiri.

**Keywords:** near-infrared spectroscopy, skin cancer, actinic keratosis, nested cross-validation, patient-level validation, feature-importance stability, machine learning

## Abstract

**Background:** Portable near-infrared (NIR) spectroscopy has been proposed as an adjunct for skin-lesion triage, and the public NIR-SC-UFES dataset has made in vivo NIR spectra of skin lesions available for machine-learning benchmarks. Two questions that bear on how such benchmarks should be read have received little attention: how results depend on the labeling of actinic keratosis (ACK), a pre-malignant lesion that the dataset convention places in the non-cancer group, and whether the wavelengths that classifiers rank as important are stable enough to support spectroscopic interpretation.

**Methods:** We analyzed the original NIR-SC-UFES file (714 in vivo spectra, 331 patients, 908–1676 nm, 125 points; MicroNIR Pro 1700). After standard normal variate (SNV) normalization, five classifiers (RBF support vector machine, logistic regression, random forest, PLS-DA and a one-dimensional convolutional neural network) were evaluated with nested cross-validation in which every patient was confined to a single outer fold (5 outer × 3 inner folds; hyperparameters selected on inner folds only). We compared four labelings of ACK and two pairwise tasks, measured the stability of band importance over 60 patient-level bootstrap resamples, and tested the effect of removing top-ranked, random or contiguous bands.

**Results:** On the standard binary task (647 spectra, 319 patients) the SVM reached macro-F1 0.764 ± 0.005, balanced accuracy 0.772 ± 0.006 and ROC-AUC 0.842 ± 0.006 (mean ± SD over five repetitions of the outer split); sensitivity was 0.72 and specificity 0.83. Logistic regression, random forest and the 1D-CNN were within the fold-to-fold variability of the SVM (paired Wilcoxon test, SVM vs. CNN, p = 0.63; patient-bootstrap 95% CI of the AUC difference −0.006 to 0.064). The labeling of ACK changed performance more than the choice of classifier did: macro-F1 was 0.764 with ACK as non-cancer, 0.743 with ACK excluded and 0.677 with ACK counted as positive. ACK was more separable from malignant lesions (AUC 0.876) than benign lesions were (AUC 0.813), while benign lesions and ACK were only moderately separable (AUC 0.794). Band importance was unstable at the level of single bands (mean Jaccard overlap of the top-10 bands 0.34; mean Spearman correlation 0.51), yet the top-ranked band fell within 908–1051 nm in 59 of 60 resamples. Removing the top 5–20 ranked bands lowered macro-F1 by at most 0.016, and removing 20 contiguous bands around the top band by 0.028; removing random bands had no effect.

**Conclusions:** On this cohort, classical and deep classifiers perform similarly under nested patient-level validation, while the definition of the non-cancer class has a larger effect. The informative spectral region is reproducible, but individual band rankings are not, so single-wavelength chemical assignments should be avoided. Studies on NIR-SC-UFES should state how ACK is labeled and should report band importance across resamples.

## 1. Introduction

Skin cancer is among the most frequently diagnosed cancers worldwide. The Global Burden of Disease study estimated 6.64 million new cases in 2021, with age-standardized incidence rising between 1990 and 2021 [1]. Visual and dermoscopic examination remain the clinical standard; optical spectroscopy has been studied as an objective, non-invasive adjunct because it reports on tissue composition rather than surface morphology. In the short-wave NIR range (roughly 900–1700 nm), overtone and combination bands of O–H, C–H and N–H bonds give broad absorption features sensitive to the water, lipid and protein content of tissue [2]. In vivo NIR spectra of skin lesions were shown to differ between benign, pre-malignant and malignant lesions more than two decades ago [3].

The NIR-SC-UFES dataset [4] made this approach accessible to the machine-learning community: 714 in vivo spectra of six lesion types acquired with a portable MicroNIR Pro 1700 spectrometer within the Dermatological Assistance Program (PAD) of the Federal University of Espírito Santo. The dataset’s authors benchmarked gradient-boosting models, a 1D-CNN, an SVM and PLS-DA, with SNV, feature extraction and SMOTE- or GAN-based augmentation, on an 80/20 split stratified by lesion type [5]. A further preprint classified 107-spectrum subsets of ACK, SEK and basal cell carcinoma from a public NIR database, whose class sizes match NIR-SC-UFES, using pretrained image networks on spectrum plots with a fixed spectrum-level split [14]. A concurrent study (in revision) reports a patient-level benchmark of classical and deep models on the same 647-spectrum binary subset, with nested cross-validation, a three-class analysis, calibration and external validation on a second cohort [6]. Whether classical models or CNNs are preferable under patient-level validation is therefore already being addressed, and we do not frame our work around that comparison.

Two further questions affect how any classifier trained on these data should be interpreted, and, as far as we could find in the published dataset papers and the concurrent study’s public materials, they have not been examined directly.

First, the dataset convention groups actinic keratosis (ACK) with nevi (NEV) and seborrheic keratoses (SEK) as “non-cancer”. ACK is a pre-malignant keratinocytic lesion that can progress to squamous cell carcinoma, and it makes up almost half of the dataset (296 of 714 spectra). If ACK is spectrally distinct from both benign and malignant lesions, the reported performance partly reflects a labeling decision, and “non-cancer” should not be read as “benign”.

Second, NIR classifiers are routinely interpreted by ranking wavelengths (for example with SHAP) and assigning chemical meaning to the top bands; the dataset paper identified 939–994 nm as most important for a LightGBM model [5]. Adjacent NIR bands are strongly correlated, and with a few hundred patients a ranking may change when the training patients change. If it does, single-band assignments are not supported by the data.

We address both questions with one nested, patient-level pipeline on the original data file. We report the classifier comparison as context, quantify the effect of four ACK labelings and two pairwise tasks, and measure the stability and necessity of top-ranked bands.

## 2. Materials and methods

### 2.1 Dataset

We used the original NIR-SC-UFES data file (Databrief.xls, sheet “Base”; Mendeley Data, doi:10.17632/j9773cyr3k.1, CC BY 4.0) [4]. It contains 714 spectra with 125 wavelength points between 908.1 and 1676.2 nm and seven labels: ACK (n = 296), basal cell carcinoma (CBC, n = 159), SEK (n = 107), NEV (n = 41), squamous cell carcinoma (CEC, n = 38), melanoma (MEL, n = 6) and “Others” (n = 67). “Others” was excluded because its composition is not specified, leaving 647 spectra.

According to the data providers, spectra were collected between March 2021 and December 2022; malignant diagnoses were confirmed by biopsy, and benign diagnoses by a committee of three dermatologists [5]. The file carries no unit labels; we rely on the data descriptor for the nanometer axis and plot intensities as supplied.

The file does not define the “Sample” field. Its values have the form PPP_LL (for example 331_02). Taking the prefix PPP as the patient identifier yields 331 groups, which matches the 331 patients reported in the data descriptor [4], so we used it as the patient key. Under this key the 647 retained spectra come from 319 patients (median 1, maximum 16 spectra per patient); 18 patients have both cancer and non-cancer lesions.

Provenance note. We first encountered these data as data/Databrief.csv in a public GAN-augmentation repository that describes them as Raman spectra [7]. A value-by-value comparison showed that file to be the NIR-SC-UFES file minus one nevus spectrum (sample 073_01). All analyses reported here use the original 714-row file.

### 2.2 Preprocessing

Each spectrum was transformed with the standard normal variate (SNV) [8], which removes per-spectrum offset and multiplicative scaling arising from probe contact, scattering and pigmentation. Because SNV operates on each spectrum independently it cannot leak information across folds. Features were then standardized with a scaler fitted on the training portion of each fold only.

### 2.3 Classification tasks

The standard binary task (A) follows the dataset convention: cancer = CBC, CEC, MEL (203 spectra); non-cancer = ACK, SEK, NEV (444 spectra). To study ACK labeling we defined three alternatives: (B) ACK excluded, i.e. benign (NEV, SEK) vs. malignant (351 spectra, 225 patients); (C) ACK counted as positive with the malignant lesions, against NEV and SEK (647 spectra); and (D) three classes, benign / ACK / malignant (647 spectra). Two pairwise tasks completed the picture: benign vs. ACK (444 spectra, 186 patients) and ACK vs. malignant (499 spectra, 250 patients). A six-class task over the individual lesion types was run for reference.

### 2.4 Validation protocol

All performance estimates come from nested cross-validation. In the outer loop, patients were assigned to five folds, stratified by the patient’s label (the first label listed for patients with more than one), so that every spectrum of a patient lies in exactly one outer test fold; we verified programmatically that no patient appears in both the training and test partitions of any fold. Within each outer training set, hyperparameters were selected by a 3-fold patient-grouped inner search maximizing macro-F1, the selected model was refitted on the full outer training set and evaluated once on the outer test fold. The main comparison used outer-split seed 42 for all five models, so all models were evaluated on identical folds. To separate the variability due to the random outer partition, the SVM (all tasks) and logistic regression (task A) were repeated with five outer-split seeds (42, 1, 2, 3, 4), and we report the mean ± SD over repetitions of the pooled out-of-fold metric.

### 2.5 Classifiers

- **SVM:** RBF kernel, balanced class weights [9]; inner grid C ∈ {0.1, 1, 10, 50, 100} × γ ∈ {scale, 0.001, 0.01, 0.05, 0.1} (25 configurations). Probabilities for ROC analysis came from Platt scaling.
- **Logistic regression:** L2 penalty, balanced class weights; C ∈ {0.001, 0.01, 0.1, 1, 10}.
- **Random forest:** 200 trees, balanced class weights; maximum depth ∈ {3, 5, 8, unlimited}.
- **PLS-DA:** PLS regression on the 0/1 label, thresholded at 0.5; number of latent variables ∈ {2, 4, 6, 8, 12}.
- **1D-CNN** [10]: two convolutional blocks (kernel sizes 7 and 5; batch normalization, ReLU, max pooling), global average pooling, a hidden fully connected layer with dropout and a two-unit output; Adam, cosine-annealed learning rate, class-weighted cross-entropy, batch size 32, 40 epochs. Eight configurations were searched in the inner loop (filters 8/16, 16/32 or 32/64; dropout 0.3–0.5; learning rate 5×10−^4^ or 10−^3^; weight decay 10−^3^ or 10−^2^). The final outer-fold prediction averaged three networks trained with different random seeds.

The search budgets differ between models (25 SVM vs. 8 CNN configurations), so the comparison is not budget-matched.

### 2.6 Metrics and statistics

Reporting follows TRIPOD+AI [12] where applicable. We report accuracy, balanced accuracy, macro-F1, ROC-AUC (macro one-vs-rest for multi-class tasks), sensitivity and specificity. With 31% positives, a majority-class predictor reaches 0.686 accuracy but a macro-F1 of 0.407, so macro-F1 and balanced accuracy are the primary metrics. Pairwise model comparisons used (i) a two-sided Wilcoxon signed-rank test on the five paired outer-fold macro-F1 values and (ii) a patient-level bootstrap (2,000 resamples of patients) of the difference in pooled out-of-fold ROC-AUC, reported as a 95% percentile interval. With five folds the smallest attainable two-sided Wilcoxon p-value is 0.0625, so this test can only rule out large, consistent differences.

### 2.7 Stability of band importance

Band importance was taken as the absolute coefficient of an L2 logistic regression (C = 0.1, balanced weights) fitted to standardized SNV spectra. We used a linear model because it is fast enough to refit on many resamples and its coefficients map directly onto wavelengths. We drew 60 bootstrap resamples of patients (sampling patients with replacement and keeping all their spectra) and computed (i) the mean pairwise Jaccard overlap of the top-5 and top-10 bands, (ii) the mean pairwise Spearman correlation of the full importance vectors and (iii) the distribution of the top-ranked band.

### 2.8 Band occlusion

To test whether the top-ranked bands are needed, bands were ranked within each outer training fold only (seed-42 folds), the top k (k = 5, 10, 20) were removed, and an SVM with fixed hyperparameters (C = 50, γ = scale, balanced weights; the most frequently selected C in the nested search) was retrained and tested on the remaining bands. Two controls were run with the same k: removal of k randomly chosen bands (10 draws per fold, averaged) and removal of k contiguous bands centered on the top-ranked band. We report the change in macro-F1 and ROC-AUC relative to the same SVM with all 125 bands (mean ± SD over the five folds).

### 2.9 Software and reproducibility

Analyses used Python 3 with scikit-learn 1.9, SciPy and PyTorch 2 (CPU). All random seeds are fixed in the code. The complete code, the result files (JSON, including out-of-fold predictions) and the figure scripts are provided as Supplementary Material; running three scripts on the public data file reproduces every number and figure in this paper.

## 3. Results

### 3.1 Spectra and unsupervised structure

The pooled mean spectrum shows a weak feature near 958 nm, a band near 1174 nm and a broad dominant band near 1497 nm (Figure 1a). Following published tissue band assignments [2], these lie in the regions of the second O–H overtone of water, the second C–H overtone (lipids and proteins), and the overlapping first O– H (water) and N–H (protein) overtones, respectively; given the width of NIR bands, these assignments are tentative. Before SNV, the malignant classes (CBC, MEL) lie above ACK and SEK across most of the range. After SNV the six class means nearly coincide (Figure 1b), so the classifiers rely on small differences in spectral shape rather than overall level. Level differences in raw spectra can arise from probe contact, pigmentation or hydration and are not interpreted further.

**Figure 1.**
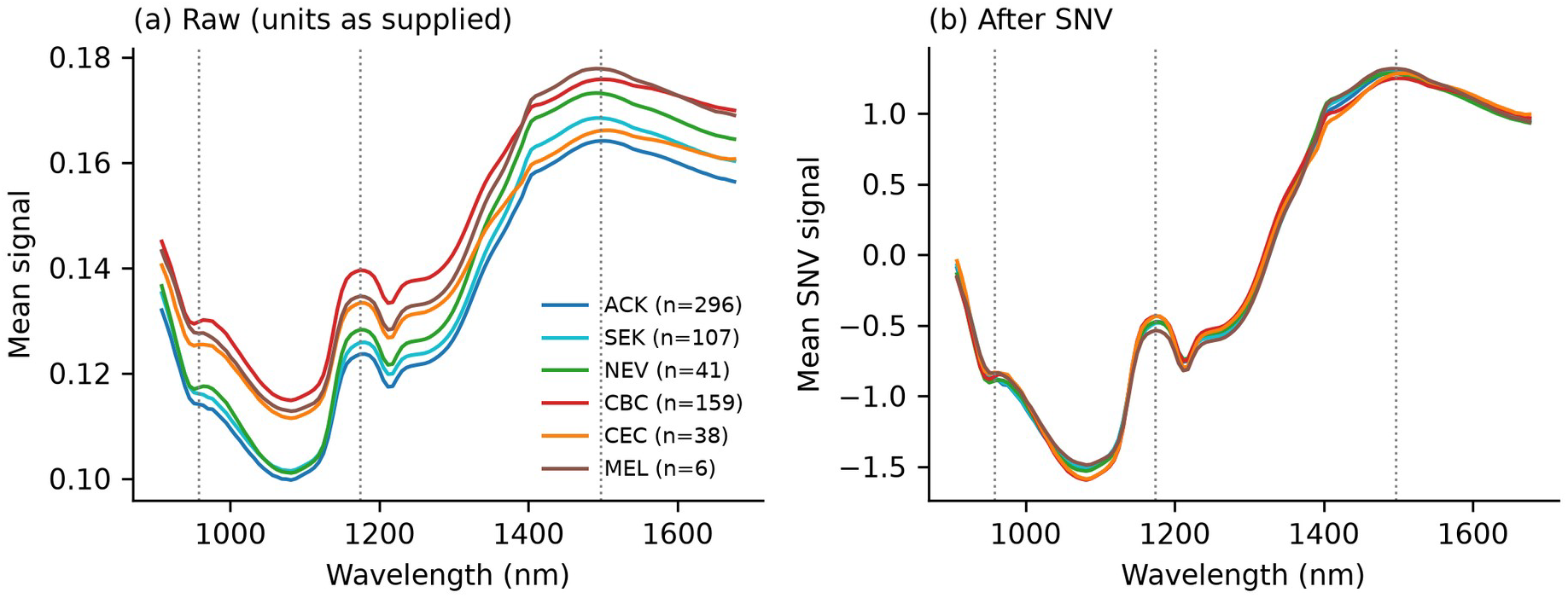
Mean NIR spectrum per lesion class, (a) as supplied in the data file and (b) after SNV. Dotted lines mark 958, 1174 and 1497 nm. Original 714-row file, “Others” excluded.

In a principal component analysis of the SNV spectra, the first two components explained 48.6% and 19.5% of the variance. Cancer and non-cancer spectra overlapped heavily, with the cancer group more dispersed (Figure 2), consistent with it pooling three distinct lesion types.

**Figure 2.**
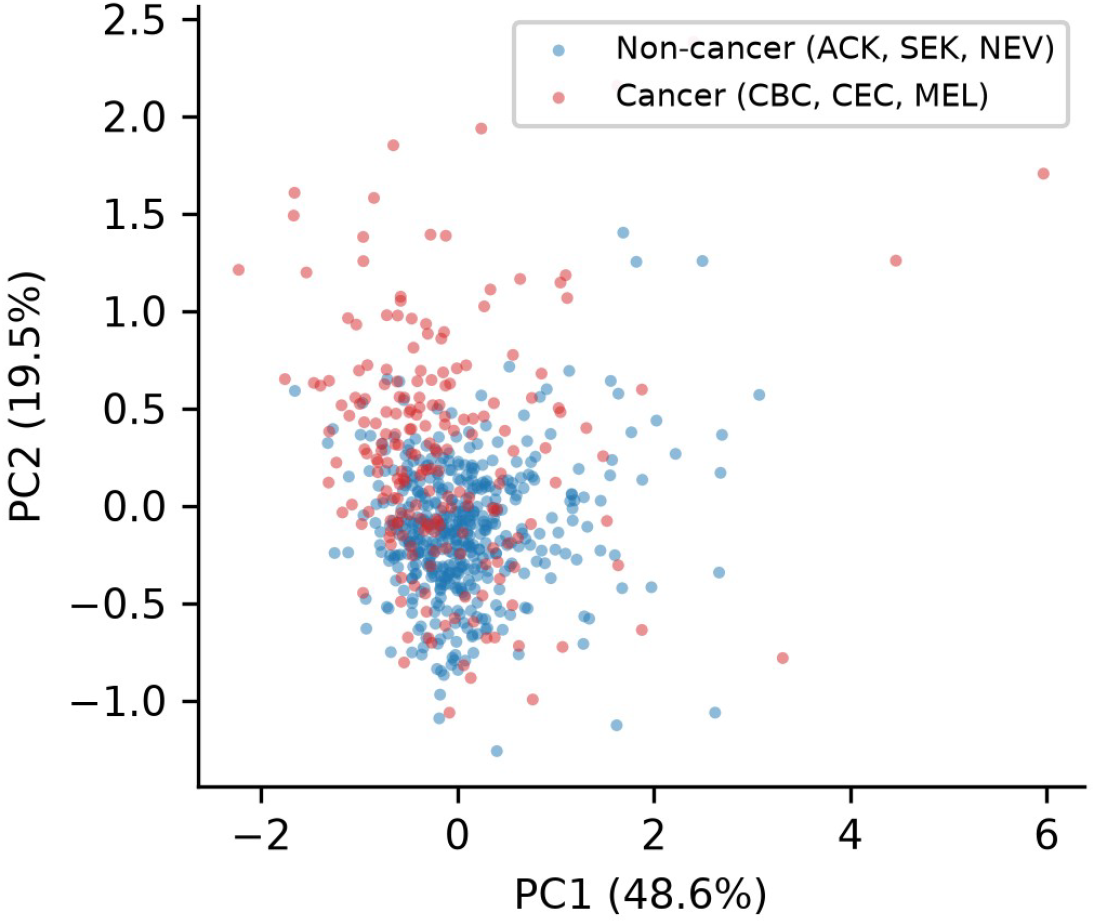
First two principal components of SNV-normalized spectra (task A, 647 spectra).

### 3.2 Classifier comparison under nested patient-level validation

Table 1 summarizes the five classifiers on the standard binary task, all evaluated on identical outer folds. All models were well above the majority-class baseline (macro-F1 0.407). The SVM had the highest mean macro-F1 and AUC, but the differences to logistic regression, random forest and the CNN were smaller than the fold-to-fold SD (0.04–0.07). PLS-DA had the highest specificity (0.95) at the cost of low sensitivity (0.42) at its default 0.5 threshold, which lowered its macro-F1.

**Table 1.** Binary task A (cancer vs. non-cancer, ACK as non-cancer; 647 spectra, 319 patients). Nested cross-validation, outer seed 42; mean ± SD over the five outer folds. Sensitivity (Sens.), specificity (Spec.) and pooled AUC are computed from the pooled out-of-fold predictions. Acc., accuracy; Bal. acc., balanced accuracy.

| Model | Acc. | Bal. acc. | Macro-F1 | AUC (per fold) | Pooled AUC | Sens. | Spec. |
| --- | --- | --- | --- | --- | --- | --- | --- |
| SVM (RBF) | 0.787 ± 0.044 | 0.764 ± 0.043 | 0.758 ± 0.038 | 0.842 ± 0.049 | 0.843 | 0.700 | 0.831 |
| Logistic regression | 0.776 ± 0.066 | 0.765 ± 0.072 | 0.752 ± 0.066 | 0.834 ± 0.092 | 0.834 | 0.729 | 0.802 |
| Random forest | 0.776 ± 0.046 | 0.739 ± 0.039 | 0.739 ± 0.038 | 0.822 ± 0.072 | 0.823 | 0.635 | 0.845 |
| 1D-CNN | 0.770 ± 0.040 | 0.730 ± 0.045 | 0.731 ± 0.044 | 0.814 ± 0.048 | 0.813 | 0.631 | 0.838 |
| PLS-DA | 0.781 ± 0.046 | 0.684 ± 0.033 | 0.702 ± 0.041 | 0.810 ± 0.087 | 0.807 | 0.424 | 0.948 |
| Majority class | 0.686 | 0.500 | 0.407 | – | – | 0.000 | 1.000 |

Selected hyperparameters varied across outer folds (SVM: C = 50 in four folds and C = 1 in one, with γ = 0.001 in four; logistic regression: C between 0.01 and 1; random forest: depth 3 to unlimited), whereas the CNN selected the same configuration (32/64 filters, dropout 0.4, learning rate 10−^3^, weight decay 10−^3^) in all five folds.

The SVM exceeded the CNN in three of five folds (per-fold macro-F1 differences −0.007, +0.140, +0.010, +0.025, −0.029; Wilcoxon W = 5, p = 0.63).

The patient-bootstrap 95% intervals of the pooled-AUC difference (SVM minus comparator) included zero for logistic regression (0.008; −0.022 to 0.041), random forest (0.020; −0.008 to 0.049) and the CNN (0.029; −0.006 to 0.064), and excluded it narrowly for PLS-DA (0.036; 0.003 to 0.070). Pooled ROC curves are shown in Figure 3. Repeating the outer split with five seeds gave macro-F1 0.764 ± 0.005, balanced accuracy 0.772 ± 0.006 and AUC 0.842 ± 0.006 for the SVM (sensitivity 0.718, specificity 0.825), and 0.755 ± 0.013, 0.769 ± 0.014 and 0.840 ± 0.011 for logistic regression.

**Figure 3.**
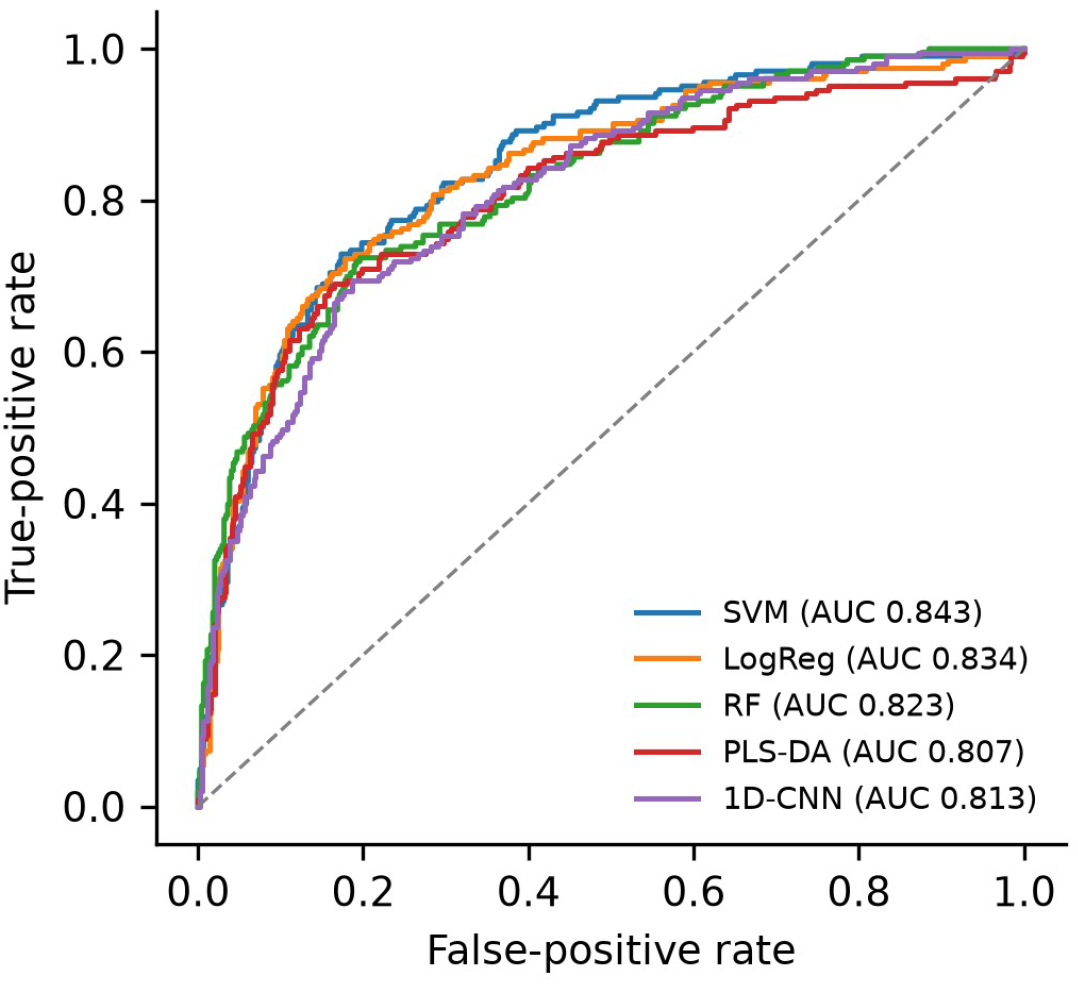
ROC curves from pooled nested out-of-fold predictions, task A, identical outer folds for all models.

For reference, the six-class SVM reached macro-F1 0.395, balanced accuracy 0.431 (chance 0.167) and macro one-vs-rest AUC 0.801. Recall was 0.70 for ACK, 0.53 for CBC, 0.40 for SEK, 0.37 for NEV, 0.26 for CEC and 2 of 6 spectra for MEL. The most frequent confusions were CBC predicted as CEC (33 of 159), SEK predicted as ACK (34 of 107) and ACK predicted as SEK (31 of 296). With six melanoma spectra no melanoma-specific conclusion is possible.

### 3.3 The labeling of actinic keratosis

The way ACK is labeled changed performance by more than the spread between classifiers (Table 2, Figure 4). Counting ACK as positive (task C) lowered macro-F1 from 0.764 to 0.677 and AUC from 0.842 to 0.766. Excluding ACK (task B) lowered macro-F1 to 0.743 and AUC to 0.813; this task is smaller (351 spectra), so part of the drop may reflect fewer training patients.

**Table 2.** SVM, nested patient-level cross-validation, mean ± SD over five outer-split seeds. Benign = NEV, SEK; malignant = CBC, CEC, MEL. For task D, the AUC is the macro one-vs-rest average.

| Task | Spectra | Patients | Macro-F1 | Bal. acc. | ROC-AUC |
| --- | --- | --- | --- | --- | --- |
| A: ACK as non-cancer (standard) | 647 | 319 | 0.764 ± 0.005 | 0.772 ± 0.006 | 0.842 ± 0.006 |
| B: ACK excluded (benign vs. malignant) | 351 | 225 | 0.743 ± 0.013 | 0.747 ± 0.014 | 0.813 ± 0.010 |
| C: ACK as positive | 647 | 319 | 0.677 ± 0.011 | 0.710 ± 0.012 | 0.766 ± 0.012 |
| D: benign / ACK / malignant | 647 | 319 | 0.656 ± 0.009 | 0.661 ± 0.010 | 0.823 ± 0.004 |
| Benign vs. ACK | 444 | 186 | 0.729 ± 0.020 | 0.739 ± 0.021 | 0.794 ± 0.019 |
| ACK vs. malignant | 499 | 250 | 0.802 ± 0.010 | 0.805 ± 0.011 | 0.876 ± 0.007 |

**Figure 4.**
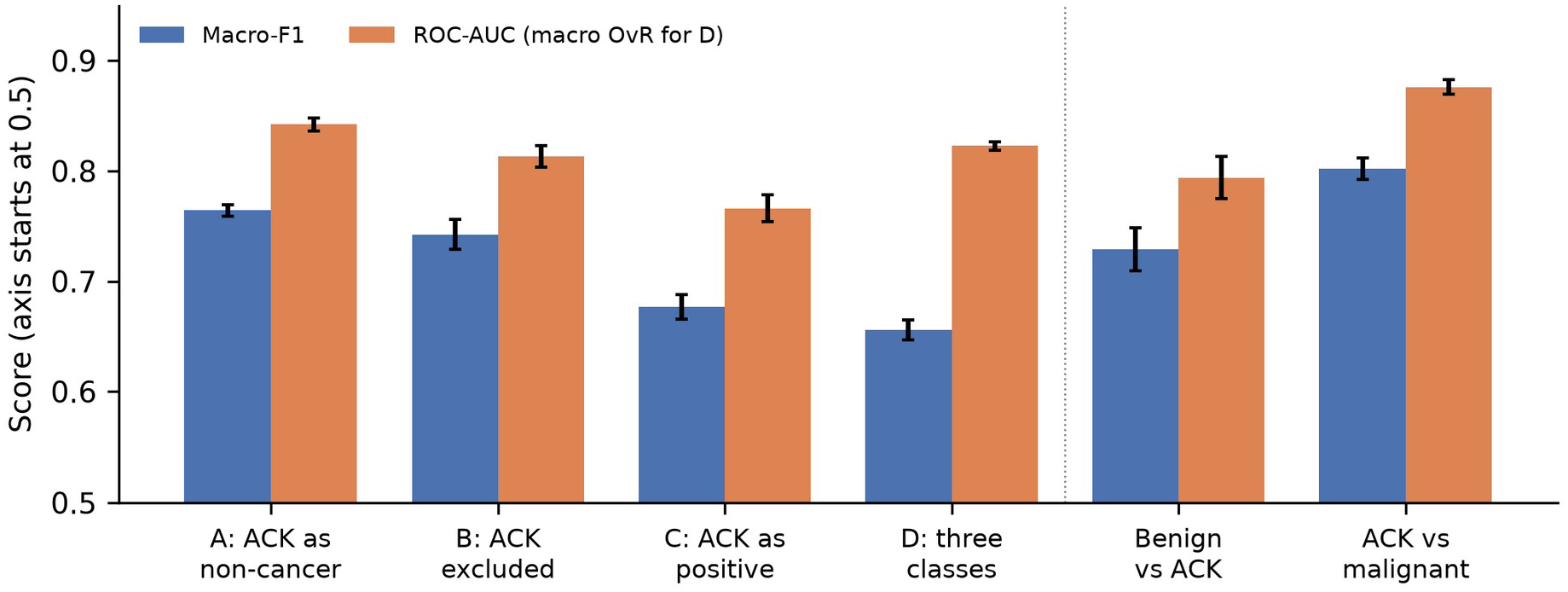
Macro-F1 and ROC-AUC for the four ACK labelings (left) and the two pairwise tasks involving ACK (right). Error bars: SD over five outer-split seeds. The vertical axis starts at 0.5.

The pairwise tasks explain this pattern. ACK was the most separable from the malignant lesions (AUC 0.876), more so than the benign lesions were (benign vs. malignant, AUC 0.813). Benign lesions and ACK were only moderately separable (AUC 0.794). These pairwise results are consistent with an ordering in which the benign lesions lie between ACK and the malignant lesions, rather than ACK lying between benign and malignant lesions as its clinical status might suggest. Including ACK as non-cancer makes the binary task easier, because a large group of non-cancer spectra is far from the malignant group, and counting ACK as positive makes it harder, because it places lesions that were readily separable in this cohort in the same class. The three-class formulation (task D) reached macro-F1 0.656 and balanced accuracy 0.661 (chance 0.333), with a macro one-vs-rest AUC of 0.823.

The SDs in Table 2 reflect the choice of outer partition only. Variation across folds within one partition is larger (about 0.04 in macro-F1, Table 1), so differences of a few hundredths between tasks B and A should not be over-interpreted; the gaps involving task C and the ACK-vs-malignant task are several times larger.

### 3.4 Stability and necessity of top-ranked bands

Under patient-level bootstrap resampling, band rankings were unstable at the level of individual bands (Figure 5a). The mean pairwise Jaccard overlap was 0.24 for the top-5 bands and 0.34 for the top-10 bands, and the mean Spearman correlation between importance vectors was 0.51. The top-ranked band was 1032.0 nm in 24 of 60 resamples and 1050.6 nm in 12, and twelve different bands were top-ranked at least once. The region was much more stable than the band: in 59 of 60 resamples the top-ranked band lay between 908 and 1051 nm. Within the five outer training folds, which share about 80% of their patients, 1032.0 and 1050.6 nm were among the top five bands in every fold. A secondary cluster near 1571–1590 nm was also consistently weighted (Figure 5a).

**Figure 5.**
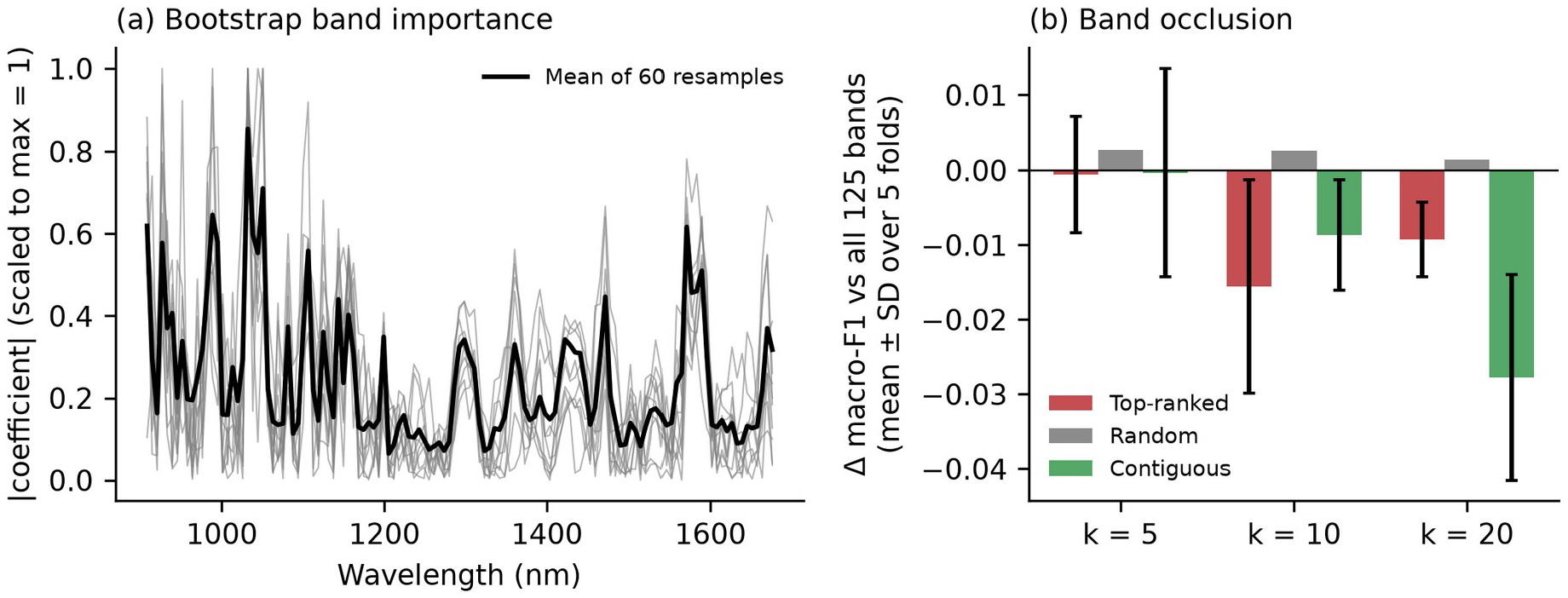
(a) Absolute logistic-regression coefficients for 10 of the 60 patient-bootstrap resamples (grey) and the mean over all 60 (black), each scaled to a maximum of 1. (b) Change in SVM macro-F1 after removing the top-k ranked bands, k random bands or k contiguous bands around the top band; bands ranked on training folds only. Error bars: SD over five folds (random-band removal averaged over 10 draws per fold).

Removing the highest-ranked bands had only a small effect (Figure 5b). With all 125 bands, the fixed-hyperparameter SVM reached macro-F1 0.777 and AUC 0.848 on the seed-42 folds. Removing the top 5, 10 and 20 bands changed macro-F1 by −0.001 ± 0.008, −0.016 ± 0.014 and −0.009 ± 0.005, and AUC by −0.005, −0.011 and −0.016. Removing the same number of random bands changed macro-F1 by +0.003, +0.003 and +0.001 (AUC within ±0.002). Removing a contiguous block of 20 bands around the top band (about 120 nm) produced the largest loss (macro-F1 −0.028 ± 0.014; AUC −0.017). The top-ranked bands therefore carry some information that random bands do not, but most of it is also present in correlated neighboring and distant bands, and the classifier remains within about 0.03 of its full-spectrum performance when they are removed.

## 4. Discussion

### 4.1 Label definition matters more than model choice

Under nested patient-level validation, four of the five classifiers were statistically indistinguishable on this cohort, and the spread between them (about 0.03 in macro-F1 between SVM and CNN, smaller between SVM and logistic regression) was smaller than the effect of how ACK was labeled (0.09 in macro-F1 between tasks A and C). The finding that the classical models are at least as good as a 1D-CNN on a few hundred patients agrees with the dataset authors’ benchmark, in which the SVM had higher balanced accuracy than the 1D-CNN in most settings without feature extraction [5], and with the concurrent patient-level study [6].

Our nested estimates are lower than figures obtained with spectrum-level or non-nested protocols, which is expected and, in our view, closer to performance on new patients.

The pairwise results indicate that the standard “non-cancer” class is heterogeneous: in these data, ACK was separated from the malignant lesions more readily than from the benign lesions (AUC 0.876 vs 0.794). This is consistent with, but does not establish, a closer spectral resemblance between ACK and the benign lesions; ACK was nonetheless distinguishable from NEV and SEK, so the two groups should not be treated as interchangeable. This matters for interpretation in two ways. A reported cancer-vs-non-cancer accuracy on NIR-SC-UFES partly reflects the large, comparatively easy ACK group, and a model that performs well on the standard task is not thereby shown to separate pre-malignant from benign lesions. Early in vivo NIR work already treated ACK as a separate pre-malignant group [3]. We suggest that studies on this dataset report the ACK labeling explicitly and include at least one alternative labeling or the three-class task.

The spectral reason for ACK’s position cannot be established from these data. ACK is a hyperkeratotic lesion of the epidermis, and differences in stratum-corneum thickness, hydration or lipid content could plausibly alter the short-wave NIR shape, but anatomical site, age and Fitzpatrick type, which may also differ between lesion types, are not available in the public file. The ACK effect may therefore partly reflect such covariates rather than lesion biology.

### 4.2 Reproducible regions, unreproducible bands

The bootstrap analysis separates two levels of interpretation. At the level of a spectral region, the result is reproducible: the 908–1051 nm range carried the top-ranked band in 59 of 60 resamples, which is consistent with the dataset authors’ report that 939–994 nm was most important for a different model and feature set [5], and with an independent near-infrared hyperspectral imaging study in which 900–1085 nm was among the regions differing most between non-melanoma skin cancer, actinic keratosis and healthy skin [11]. At the level of a single wavelength it is not: the top band changed across resamples, top-10 sets shared only about a third of their members, and removing the top-ranked bands cost at most about 0.016 in macro-F1. NIR overtone and combination bands are broad and the 125 points of the MicroNIR are strongly correlated, so importance is spread across neighboring channels and can shift between them with small changes in the training patients.

The practical recommendation is that wavelength-importance results on cohorts of this size be reported as distributions over resamples and at the level of regions, with a removal control, before any chemical assignment is attached. The 908 nm channel, the first point of the detector range, was occasionally top-ranked; edge channels may be affected by detector response and should be interpreted with particular caution. Our importance measure was the coefficient magnitude of a linear model; SHAP values for a kernel model may rank bands differently, and we did not test whether they are more stable.

### 4.3 Relation to prior and concurrent work

The dataset paper [5] and its data descriptor [4] introduced NIR-SC-UFES and its first benchmarks; the arXiv version of the dataset paper describes 971 samples, whereas the public file contains 714, so our numbers are not directly comparable with it. The concurrent study [6] covers nested and repeated cross-validation, calibration, an ablation, a three-class analysis and external validation on a second spectral cohort. Its public reproduction package does not describe an ACK-labeling comparison or a band-stability analysis, but we have not seen the full manuscript and parts of our label analysis, in particular the three-class task, may overlap with it. The image-network preprint [14] treats ACK as one of three classes but does not compare labelings or assess importance stability, and its split is not described as patient-level. The contributions we claim are the systematic comparison of ACK labelings with pairwise tasks, and the stability and occlusion analysis of band importance, both under one nested, patient-level protocol with released code.

### 4.4 Limitations

All results come from one cohort in one dermatology program, without external validation. Patients were identified from the sample-code prefix; this assumption reproduces the published patient count but is not documented in the data file.

Melanoma has only six spectra, so nothing can be said about melanoma detection, and the “Others” category was excluded. The outer cross-validation has five folds, which limits the power of paired tests. Model search budgets differ between classifiers. Calibration of predicted probabilities was not assessed, so the reported probabilities should not be read as risks. Hyperparameters of the occlusion SVM were fixed rather than nested, so its absolute performance (0.777) is slightly optimistic; the analysis concerns the differences, which share the same bias. Clinical covariates (age, sex, anatomical site, skin type) were not available for adjustment. A multimodal image-and-NIR dataset from the same program has recently been released [13]; because it was collected within the same program and may share patients, we did not use it as an external test set, as its independence from NIR-SC-UFES could not be established from the public descriptions.

## 5. Conclusions

On the public NIR-SC-UFES cohort, nested patient-level validation gives cancer-vs-non-cancer macro-F1 of about 0.76 and ROC-AUC of about 0.84, with classical models and a 1D-CNN performing similarly. How actinic keratosis is labeled has a larger effect on these figures than the choice of classifier, probably because ACK spectra were separable, to differing degrees, from both benign and malignant lesions in this cohort. The informative spectral region (908–1051 nm) is reproducible across patient resamples, but individual top-ranked bands are not and can be removed with little loss. Reports on this and similar datasets should state the labeling of pre-malignant lesions, use patient-level nested validation, and present band importance as a resampled, region-level result.

## Supporting information

Supplementary code and result files

## Declarations

All data analyzed in this study are publicly available: NIR-SC-UFES, Mendeley Data, version 1, https://doi.org/10.17632/j9773cyr3k.1 (file Databrief.xls, sheet “Base”; CC BY 4.0) [4]. No new data were generated. All code, result files (including out-of-fold predictions) and figure scripts needed to reproduce every number and figure are provided as a supplementary file with this preprint.

## Acknowledgements

We thank the NIR-SC-UFES team and the Dermatological and Surgical Assistance Program (PAD-UFES) for making their data publicly available.

## Ethics statement

This study is a secondary analysis of a fully de-identified dataset that is publicly available under a CC BY 4.0 licence [4]. No new data were collected, the authors had no access to identifiable information and no contact with participants, so no additional ethics committee approval or informed consent was required for this analysis. The original data were collected within the Dermatological and Surgical Assistance Program (PAD) of the Federal University of Espírito Santo, which, according to the data providers, was approved by the university ethics committee (no. 500002/478) and by Plataforma Brasil (no. 4.007.097), with patient consent [4, 13]. All relevant ethical guidelines were followed. The results are a methodological analysis and are not intended to support clinical decisions.

## Clinical trial registration

Not applicable; this is not a clinical trial.

## Reporting guideline

A reporting checklist based on TRIPOD+AI is provided as a supplementary file.

## Author contributions

S.R.A.: conceptualization, methodology, software, formal analysis, visualization, writing – original draft. A.K.: writing – review and editing.

