## Supplementary code and result files for "Actinic keratosis labeling and single-band instability in near-infrared spectroscopy of skin cancer: a nested, patient-level re-analysis"

This document reproduces, as plain text, the complete Python code and all result files used for the numbers and figures in the manuscript. Data: NIR-SC-UFES, Mendeley Data, https://doi.org/10.17632/j9773cyr3k.1 (file Databrief.xls, sheet “Base”).

#### S1. README

```
# Reproduction package

Manuscript: manuscript.md (figures in figures/).
Data: NIR-SC-UFES, Mendeley Data doi:10.17632/j9773cyr3k.1 (Databrief.xls, sheet "Base"). Not included; set
NIR_DATA=/path/to/Databrief.xls (or a CSV export).
Run (from code/): python classical_and_sensitivity.py; python cnn_nested.py; python post_analysis.py; python
make_figures.py
Requirements: Python 3, numpy, pandas, scipy, scikit-learn (1.9 used), torch (2.x CPU), matplotlib, xlrd (for .xls).
Runtime: roughly 15 min (classical) and 25 min (CNN) on 2 CPU cores.
```

#### S2. code/common.py

```python
"""Shared data loading and patient-grouped fold generation for NIR-SC-UFES (Databrief.xls / .csv, 714 rows)."""
import numpy as np, pandas as pd, os
DATA = os.environ.get('NIR_DATA', '/home/user/workspace/nir/Databrief.csv')
RNG = 42
CAN = {'CBC', 'CEC', 'MEL'}; BEN = {'NEV', 'SEK'}; ACK = {'ACK'}
def load_df():
    df = pd.read_csv(DATA) if DATA.endswith('.csv') else pd.read_excel(DATA, sheet_name='Base')
    df['PatientID'] = df['Sample'].astype(str).str.split('_').str[0]
    cols = [c for c in df.columns if c not in ('N', 'Sample', 'Class', 'yClass', 'PatientID')]
    return df, cols
def snv(X):
    X = np.asarray(X, float); return (X - X.mean(1, keepdims=True)) / X.std(1, keepdims=True)
def task(df, cols, name):
    """Return X (SNV), y, groups, class names for a labeling scheme."""
    c = df['Class']
    if name == 'A':   keep, lab = c.isin(CAN | BEN | ACK), c.isin(CAN).astype(int)
    elif name == 'B': keep, lab = c.isin(CAN | BEN), c.isin(CAN).astype(int)
    elif name == 'C': keep, lab = c.isin(CAN | BEN | ACK), c.isin(CAN | ACK).astype(int)
    elif name == 'D': keep, lab = c.isin(CAN | BEN | ACK), c.map(lambda v: 0 if v in BEN else (1 if v in ACK else 2))
    elif name == 'BEN_vs_ACK': keep, lab = c.isin(BEN | ACK), c.isin(ACK).astype(int)
    elif name == 'ACK_vs_MAL': keep, lab = c.isin(ACK | CAN), c.isin(CAN).astype(int)
    elif name == 'SIX':
        order = ['ACK', 'CBC', 'CEC', 'MEL', 'NEV', 'SEK']
        keep, lab = c.isin(order), c.map({k: i for i, k in enumerate(order)})
    d = df[keep]
    return snv(d[cols].values), np.asarray(lab[keep]).astype(int), d['PatientID'].values
def pkfold(groups, y, k, seed):
    """Patient-grouped, label-stratified k-fold: every patient is wholly in one fold."""
    rng = np.random.RandomState(seed)
    pl = pd.Series(y, index=groups).groupby(level=0).first()
    folds = [[] for _ in range(k)]
    for lab in sorted(pl.unique()):
        pats = pl[pl == lab].index.tolist(); rng.shuffle(pats)
        for i, p in enumerate(pats): folds[i % k].append(p)
    for i in range(k):
        te = np.isin(groups, folds[i]); yield ~te, te
```

### S3. code/classical_and_sensitivity.py

```python
"""Nested patient-grouped CV for classical models (task A), 6-class SVM, actinic-keratosis labeling analysis,
and band-importance stability / occlusion. All on the original 714-row NIR-SC-UFES file."""
import numpy as np, json, itertools, warnings; warnings.filterwarnings('ignore')
from common import *
from scipy.stats import spearmanr
from sklearn.preprocessing import StandardScaler
from sklearn.svm import SVC
from sklearn.linear_model import LogisticRegression
from sklearn.ensemble import RandomForestClassifier
from sklearn.cross_decomposition import PLSRegression
from sklearn.metrics import f1_score, accuracy_score, roc_auc_score, balanced_accuracy_score, confusion_matrix
df, cols = load_df(); wl = np.array([float(c) for c in cols])
GRIDS = {'SVM': [dict(C=C, gamma=ga) for C in [0.1, 1, 10, 50, 100] for ga in ['scale', 0.001, 0.01, 0.05, 0.1]],
         'LogReg': [dict(C=C) for C in [0.001, 0.01, 0.1, 1, 10]],
         'RF': [dict(max_depth=d) for d in [3, 5, 8, None]],
         'PLS-DA': [dict(n=n) for n in [2, 4, 6, 8, 12]]}
def fit_score(name, kw, Xtr, ytr, Xte, multiclass=False, need_score=True):
    """Return (pred, score). score: P(class 1) for binary, class-probability matrix for multiclass."""
    sc = StandardScaler().fit(Xtr); A, B = sc.transform(Xtr), sc.transform(Xte)
    if name == 'PLS-DA':
        s = PLSRegression(n_components=kw['n']).fit(A, ytr).predict(B).ravel(); return (s > 0.5).astype(int), s
    if name == 'SVM': m = SVC(kernel='rbf', class_weight='balanced', probability=need_score, random_state=RNG, **kw)
    if name == 'SVM' and not need_score:
        m.fit(A, ytr); return m.predict(B), None
    if name == 'LogReg': m = LogisticRegression(max_iter=3000, class_weight='balanced', random_state=RNG, **kw)
    if name == 'RF': m = RandomForestClassifier(n_estimators=200, class_weight='balanced', random_state=RNG, **kw)
    m.fit(A, ytr); P = m.predict_proba(B)
    return m.predict(B), (P if multiclass else P[:, 1])
def nested(name, X, y, g, seed, multiclass=False):
    pred = np.zeros(len(y), int); score = np.zeros((len(y), len(np.unique(y)))) if multiclass else np.zeros(len(y))
    folds, chosen = [], []
    for tr, te in pkfold(g, y, 5, seed):
        idx = np.where(tr)[0]; best, bk = -1, None
        for kw in GRIDS[name]:
            s = [f1_score(y[idx[ite]], fit_score(name, kw, X[idx[itr]], y[idx[itr]], X[idx[ite]], multiclass,
need_score=False)[0], average='macro')
                 for itr, ite in pkfold(g[tr], y[tr], 3, seed + 1)]
            if np.mean(s) > best: best, bk = np.mean(s), kw
        p, s = fit_score(name, bk, X[tr], y[tr], X[te], multiclass); pred[te] = p; score[te] = s;
chosen.append(str(bk))
        auc = roc_auc_score(y[te], s, multi_class='ovr', average='macro') if multiclass else roc_auc_score(y[te], s)
        folds.append(dict(f1=f1_score(y[te], p, average='macro'), acc=accuracy_score(y[te], p),
                          bacc=balanced_accuracy_score(y[te], p), auc=auc))
    return pred, score, folds, chosen
def pooled(y, pred, score, multiclass=False):
    r = dict(f1=f1_score(y, pred, average='macro'), acc=accuracy_score(y, pred), bacc=balanced_accuracy_score(y,
pred),
             auc=roc_auc_score(y, score, multi_class='ovr', average='macro') if multiclass else roc_auc_score(y,
score))
    if not multiclass: r.update(sens=float((pred[y == 1] == 1).mean()), spec=float((pred[y == 0] == 0).mean()))
    return r
R = {}
# ---- 1. Main binary comparison (task A), outer seed 42 (same folds as CNN) ----
X, y, g = task(df, cols, 'A')
for tr, te in pkfold(g, y, 5, RNG): assert not set(g[tr]) & set(g[te])
R['dataset'] = dict(n_spectra=int(len(y)), n_patients=int(len(set(g))), n_cancer=int(y.sum()),
                    majority_f1=float(f1_score(y, np.zeros_like(y), average='macro')), majority_acc=float(1 -
y.mean()))
R['main'] = {}
for name in GRIDS:
    pred, score, folds, chosen = nested(name, X, y, g, RNG)
    R['main'][name] = dict(folds=folds, chosen=chosen, pooled=pooled(y, pred, score), oof=score.tolist())
    print('main', name, {k: round(np.mean([f[k] for f in folds]), 4) for k in folds[0]}, chosen, flush=True)
R['y_A'] = y.tolist()
# ---- 2. Repeated outer splits (5 seeds) for SVM and LogReg ----
R['repeat'] = {}
for name in ['SVM', 'LogReg']:
    R['repeat'][name] = [pooled(y, *nested(name, X, y, g, s)[:2]) for s in [42, 1, 2, 3, 4]]
    print('repeat', name, {k: (round(np.mean([r[k] for r in R['repeat'][name]]), 4), round(np.std([r[k] for r in
R['repeat'][name]]), 4)) for k in R['repeat'][name][0]}, flush=True)
# ---- 3. Six-class task, SVM ----
X6, y6, g6 = task(df, cols, 'SIX')
pred, score, folds, chosen = nested('SVM', X6, y6, g6, RNG, multiclass=True)
cm = confusion_matrix(y6, pred)
R['six'] = dict(pooled=pooled(y6, pred, score, True), folds=folds, chosen=chosen, cm=cm.tolist(),
                recall=(cm.diagonal() / cm.sum(1)).tolist(), classes=['ACK', 'CBC', 'CEC', 'MEL', 'NEV', 'SEK'],
n=int(len(y6)))
print('six', R['six']['pooled'], R['six']['recall'], flush=True)
# ---- 4. Actinic-keratosis labeling analysis (SVM, 5 outer seeds) ----
```

```
R['label'] = {}
for t in ['A', 'B', 'C', 'D', 'BEN_vs_ACK', 'ACK_vs_MAL']:
    Xt, yt, gt = task(df, cols, t); mc = (t == 'D')
    rs = [pooled(yt, *nested('SVM', Xt, yt, gt, s, mc)[:2], mc) for s in [42, 1, 2, 3, 4]]
    R['label'][t] = dict(n=int(len(yt)), patients=int(len(set(gt))),
                         mean={k: float(np.mean([r[k] for r in rs])) for k in ['f1', 'bacc', 'auc']},
                         sd={k: float(np.std([r[k] for r in rs])) for k in ['f1', 'bacc', 'auc']})
    print('label', t, R['label'][t], flush=True)
# ---- 5. Band-importance stability (patient bootstrap, LogReg |coef|) ----
rng = np.random.RandomState(RNG); pats = np.unique(g); imps = []
for b in range(60):
    bp = rng.choice(pats, len(pats), replace=True)
    idx = np.concatenate([np.where(g == p)[0] for p in bp])
    sc = StandardScaler().fit(X[idx])
    m = LogisticRegression(C=0.1, class_weight='balanced', max_iter=3000).fit(sc.transform(X[idx]), y[idx])
    imps.append(np.abs(m.coef_.ravel()))
imps = np.array(imps)
def jac(k):
    tops = [set(np.argsort(-v)[:k]) for v in imps]
    return float(np.mean([len(a & b) / len(a | b) for a, b in itertools.combinations(tops, 2)]))
sp = [spearmanr(a, b)[0] for a, b in itertools.combinations(imps, 2)]
top1 = np.argmax(imps, 1); vals, cnt = np.unique(top1, return_counts=True)
R['stability'] = dict(jaccard5=jac(5), jaccard10=jac(10), spearman=float(np.mean(sp)),
                      top1={f'{wl[v]:.1f}': int(c) for v, c in sorted(zip(vals, cnt), key=lambda t: -t[1])},
                      top1_908_1051=int(np.sum(wl[top1] <= 1051.5)), imps=imps.tolist(), wl=wl.tolist())
print('stability', {k: v for k, v in R['stability'].items() if k not in ('imps', 'wl')}, flush=True)
# ---- 6. Occlusion test (SVM C=50, gamma='scale'; bands ranked on training folds only) ----
def svm_eval(Xtr, ytr, Xte, yte):
    sc = StandardScaler().fit(Xtr); m = SVC(kernel='rbf', C=50, gamma='scale', class_weight='balanced',
random_state=RNG).fit(sc.transform(Xtr), ytr)
    return f1_score(yte, m.predict(sc.transform(Xte)), average='macro'), roc_auc_score(yte,
m.decision_function(sc.transform(Xte)))
occ = {'all': [], **{f'{kind}{k}': [] for kind in ['top', 'rand', 'contig'] for k in [5, 10, 20]}}
rr = np.random.RandomState(7)
for tr, te in pkfold(g, y, 5, RNG):
    occ['all'].append(svm_eval(X[tr], y[tr], X[te], y[te]))
    sc = StandardScaler().fit(X[tr]); imp = np.abs(LogisticRegression(C=0.1, class_weight='balanced',
max_iter=3000).fit(sc.transform(X[tr]), y[tr]).coef_.ravel())
    order = np.argsort(-imp); n = X.shape[1]
    for k in [5, 10, 20]:
        keep = np.setdiff1d(np.arange(n), order[:k]); occ[f'top{k}'].append(svm_eval(X[tr][:, keep], y[tr], X[te][:,
keep], y[te]))
        rs = []
        for _ in range(10):
            keep = np.setdiff1d(np.arange(n), rr.choice(n, k, replace=False)); rs.append(svm_eval(X[tr][:, keep],
y[tr], X[te][:, keep], y[te]))
        occ[f'rand{k}'].append(tuple(np.mean(rs, 0)))
        c0 = int(np.clip(order[0] - k // 2, 0, n - k)); keep = np.setdiff1d(np.arange(n), np.arange(c0, c0 + k))
        occ[f'contig{k}'].append(svm_eval(X[tr][:, keep], y[tr], X[te][:, keep], y[te]))
base = np.mean(occ['all'], 0)
R['occlusion'] = dict(base_f1=float(base[0]), base_auc=float(base[1]),
                      delta={k: [float(np.mean(v, 0)[0] - base[0]), float(np.mean(v, 0)[1] - base[1])] for k, v in
occ.items() if k != 'all'})
print('occlusion', R['occlusion'], flush=True)
json.dump(R, open('../results/classical_and_sensitivity.json', 'w'))
print('done')
```

### S4. code/cnn_nested.py

```
"""Nested patient-grouped CV for the 1D-CNN (binary task A); saves per-fold metrics and out-of-fold probabilities."""
import numpy as np, json, warnings; warnings.filterwarnings('ignore')
import torch, torch.nn as nn, torch.optim as optim
from torch.utils.data import TensorDataset, DataLoader
from common import *
from sklearn.preprocessing import StandardScaler
from sklearn.metrics import f1_score, accuracy_score, roc_auc_score, balanced_accuracy_score
torch.set_num_threads(1)
df, cols = load_df(); X, y, g = task(df, cols, 'A')
class Net(nn.Module):
    def __init__(s, f1, f2, dr):
        super().__init__()
        s.n = nn.Sequential(nn.Conv1d(1, f1, 7, padding=3), nn.BatchNorm1d(f1), nn.ReLU(), nn.MaxPool1d(2),
                            nn.Conv1d(f1, f2, 5, padding=2), nn.BatchNorm1d(f2), nn.ReLU(), nn.MaxPool1d(2),
nn.AdaptiveAvgPool1d(1))
        s.f = nn.Sequential(nn.Flatten(), nn.Linear(f2, f2 // 2), nn.ReLU(), nn.Dropout(dr), nn.Linear(f2 // 2, 2))
    def forward(s, x): return s.f(s.n(x))
def train_pred(cfg, Xtr, ytr, Xte, seed, epochs=40):
    torch.manual_seed(seed); np.random.seed(seed)
    sc = StandardScaler().fit(Xtr)
    A = torch.tensor(sc.transform(Xtr), dtype=torch.float32).unsqueeze(1); B = torch.tensor(sc.transform(Xte),
dtype=torch.float32).unsqueeze(1)
```

```
    cnt = np.bincount(ytr, minlength=2); w = torch.tensor(cnt.sum() / (2 * np.maximum(cnt, 1)), dtype=torch.float32)
    m = Net(cfg['f1'], cfg['f2'], cfg['dr']); opt = optim.Adam(m.parameters(), lr=cfg['lr'], weight_decay=cfg['wd'])
    sch = optim.lr_scheduler.CosineAnnealingLR(opt, T_max=epochs); cr = nn.CrossEntropyLoss(weight=w)
    dl = DataLoader(TensorDataset(A, torch.tensor(ytr, dtype=torch.long)), batch_size=32, shuffle=True)
    for _ in range(epochs):
        m.train()
        for xb, yb in dl: opt.zero_grad(); cr(m(xb), yb).backward(); opt.step()
        sch.step()
    m.eval()
    with torch.no_grad(): return torch.softmax(m(B), 1)[:, 1].numpy()
cfgs = [dict(f1=16, f2=32, dr=0.3, lr=1e-3, wd=1e-3), dict(f1=16, f2=32, dr=0.5, lr=1e-3, wd=1e-3), dict(f1=16, f2=32,
dr=0.4, lr=5e-4, wd=1e-3),
        dict(f1=16, f2=32, dr=0.4, lr=1e-3, wd=1e-2), dict(f1=32, f2=64, dr=0.4, lr=1e-3, wd=1e-3), dict(f1=32, f2=64,
dr=0.5, lr=5e-4, wd=1e-2),
        dict(f1=8, f2=16, dr=0.3, lr=1e-3, wd=1e-3), dict(f1=16, f2=32, dr=0.4, lr=1e-3, wd=1e-3)]
out = dict(f1=[], acc=[], bacc=[], auc=[], chosen=[]); oof = np.zeros(len(y))
for k, (tr, te) in enumerate(pkfold(g, y, 5, RNG)):
    idx = np.where(tr)[0]; best, bi = -1, 0
    for ci, cfg in enumerate(cfgs):
        s = [f1_score(y[idx[ite]], (train_pred(cfg, X[idx[itr]], y[idx[itr]], X[idx[ite]], 0) > 0.5).astype(int),
average='macro')
             for itr, ite in pkfold(g[tr], y[tr], 3, RNG + 1)]
        if np.mean(s) > best: best, bi = np.mean(s), ci
    pr = np.mean([train_pred(cfgs[bi], X[tr], y[tr], X[te], seed) for seed in range(3)], 0)
    p = (pr > 0.5).astype(int); oof[te] = pr
    for key, v in [('f1', f1_score(y[te], p, average='macro')), ('acc', accuracy_score(y[te], p)),
                   ('bacc', balanced_accuracy_score(y[te], p)), ('auc', roc_auc_score(y[te], pr))]:
out[key].append(float(v))
    out['chosen'].append(bi); print('fold', k + 1, 'cfg', bi, {kk: round(out[kk][-1], 4) for kk in ['f1', 'acc',
'bacc', 'auc']}, flush=True)
out['oof'] = oof.tolist()
json.dump(out, open('../results/cnn_nested.json', 'w'))
print('CNN', {kk: (round(np.mean(out[kk]), 4), round(np.std(out[kk]), 4)) for kk in ['f1', 'acc', 'bacc', 'auc']})
```

### S5. code/post_analysis.py

```
"""Paired SVM-vs-CNN comparison on identical outer folds; per-fold occlusion deltas; patient-bootstrap CI of pooled
differences."""
import numpy as np, json, warnings; warnings.filterwarnings('ignore')
from scipy.stats import wilcoxon
from sklearn.metrics import f1_score, roc_auc_score
from sklearn.preprocessing import StandardScaler; from sklearn.svm import SVC; from sklearn.linear_model import
LogisticRegression
from common import *
R = json.load(open('../results/classical_and_sensitivity.json')); C = json.load(open('../results/cnn_nested.json'))
df, cols = load_df(); X, y, g = task(df, cols, 'A'); y = np.array(R['y_A'])
out = {}
svm_f = [f['f1'] for f in R['main']['SVM']['folds']]; cnn_f = C['f1']
w = wilcoxon(svm_f, cnn_f); out['wilcoxon_svm_cnn'] = dict(stat=float(w.statistic), p=float(w.pvalue),
diff=list(np.subtract(svm_f, cnn_f)))
for nm in ['LogReg', 'RF', 'PLS-DA']:
    f = [x['f1'] for x in R['main'][nm]['folds']]; out[f'diff_SVM_{nm}'] = list(np.subtract(svm_f, f))
# patient bootstrap of pooled OOF differences (SVM minus each model)
pats = np.unique(g); rng = np.random.RandomState(0); idxs = {p: np.where(g == p)[0] for p in pats}
scores = {nm: np.array(d['oof']) for nm, d in R['main'].items()}; scores['1D-CNN'] = np.array(C['oof'])
thr = {nm: (0.5 if nm in ('PLS-DA', '1D-CNN') else None) for nm in scores}
preds = {nm: (scores[nm] > 0.5).astype(int) for nm in scores}
# SVM/LogReg/RF pooled preds: use stored folds? recompute from probability>0.5 is not identical to predict(); store
AUC differences only plus F1 of CNN vs SVM via probs
boot = {nm: [] for nm in scores if nm != 'SVM'}
for b in range(2000):
    bi = np.concatenate([idxs[p] for p in rng.choice(pats, len(pats))])
    a_svm = roc_auc_score(y[bi], scores['SVM'][bi])
    for nm in boot: boot[nm].append(a_svm - roc_auc_score(y[bi], scores[nm][bi]))
out['auc_diff_ci'] = {nm: [float(np.mean(v)), float(np.percentile(v, 2.5)), float(np.percentile(v, 97.5))] for nm, v
in boot.items()}
out['pooled_auc'] = {nm: float(roc_auc_score(y, s)) for nm, s in scores.items()}
# per-fold occlusion (same procedure as main script, same RNG)
def ev(Xtr, ytr, Xte, yte):
    sc = StandardScaler().fit(Xtr); m = SVC(kernel='rbf', C=50, gamma='scale', class_weight='balanced',
random_state=RNG).fit(sc.transform(Xtr), ytr)
    return f1_score(yte, m.predict(sc.transform(Xte)), average='macro'), roc_auc_score(yte,
m.decision_function(sc.transform(Xte)))
rr = np.random.RandomState(7); per = {}
for tr, te in pkfold(g, y, 5, RNG):
    base = np.array(ev(X[tr], y[tr], X[te], y[te]))
    sc = StandardScaler().fit(X[tr]); imp = np.abs(LogisticRegression(C=0.1, class_weight='balanced',
max_iter=3000).fit(sc.transform(X[tr]), y[tr]).coef_.ravel())
    order = np.argsort(-imp); n = X.shape[1]
    for k in [5, 10, 20]:
        keep = np.setdiff1d(np.arange(n), order[:k]); per.setdefault(f'top{k}', []).append(np.array(ev(X[tr][:,
```

```
keep], y[tr], X[te][:, keep], y[te])) - base)
        rs = [np.array(ev(X[tr][:, np.setdiff1d(np.arange(n), rr.choice(n, k, replace=False))], y[tr], X[te][:,
np.setdiff1d(np.arange(n), rr.choice(n, k, replace=False))], y[te])) for _ in range(0)]
        c0 = int(np.clip(order[0] - k // 2, 0, n - k)); keep = np.setdiff1d(np.arange(n), np.arange(c0, c0 + k))
        per.setdefault(f'contig{k}', []).append(np.array(ev(X[tr][:, keep], y[tr], X[te][:, keep], y[te])) - base)
        per.setdefault(f'top{k}_bands', []).append([float(v) for v in np.sort(np.array([float(c) for c in cols])
[order[:k]])])
out['occlusion_perfold'] = {k: (np.array(v).tolist() if 'bands' not in k else v) for k, v in per.items()}
out['occlusion_sd'] = {k: np.array(v).std(0).tolist() for k, v in per.items() if 'bands' not in k}
json.dump(out, open('../results/post_analysis.json', 'w'), indent=1)
print(json.dumps({k: v for k, v in out.items() if k not in ('occlusion_perfold',)}, indent=1)[:3000])
print('top5 bands per fold', per['top5_bands'])
```

### S6. code/make_figures.py

```
"""Figures for the manuscript, from the original 714-row file and the JSON results."""
import numpy as np, json, os, matplotlib; matplotlib.use('Agg'); import matplotlib.pyplot as plt
from sklearn.decomposition import PCA; from sklearn.metrics import roc_curve, roc_auc_score
from common import *
plt.rcParams.update({'font.size': 9, 'axes.spines.top': False, 'axes.spines.right': False, 'savefig.dpi': 300})
F = '../figures'; os.makedirs(F, exist_ok=True)
df, cols = load_df(); wl = np.array([float(c) for c in cols])
order = ['ACK', 'SEK', 'NEV', 'CBC', 'CEC', 'MEL']; colors = dict(ACK='#1f77b4', SEK='#17becf', NEV='#2ca02c',
CBC='#d62728', CEC='#ff7f0e', MEL='#8c564b')
# Fig 1
fig, ax = plt.subplots(1, 2, figsize=(7.2, 2.9))
for c in order:
    R = df[df.Class == c][cols].values.astype(float)
    ax[0].plot(wl, R.mean(0), color=colors[c], lw=1.2, label=f'{c} (n={len(R)})'); ax[1].plot(wl, snv(R).mean(0),
color=colors[c], lw=1.2)
for a, t in zip(ax, ['(a) Raw (units as supplied)', '(b) After SNV']):
    for x in (958, 1174, 1497): a.axvline(x, ls=':', c='grey', lw=0.8)
    a.set_xlabel('Wavelength (nm)'); a.set_title(t, loc='left', fontsize=9)
ax[0].set_ylabel('Mean signal'); ax[1].set_ylabel('Mean SNV signal'); ax[0].legend(fontsize=7, frameon=False)
plt.tight_layout(); plt.savefig(f'{F}/fig1_mean_spectra.png'); plt.close()
# Fig 2
X, y, g = task(df, cols, 'A'); Z = PCA(2).fit(X); P = Z.transform(X)
fig, a = plt.subplots(figsize=(3.6, 3.1))
for lab, col, nm in [(0, '#1f77b4', 'Non-cancer (ACK, SEK, NEV)'), (1, '#d62728', 'Cancer (CBC, CEC, MEL)')]:
    a.scatter(P[y == lab, 0], P[y == lab, 1], s=7, alpha=0.5, c=col, label=nm, edgecolors='none')
a.set_xlabel(f'PC1 ({100*Z.explained_variance_ratio_[0]:.1f}%)'); a.set_ylabel(f'PC2
({100*Z.explained_variance_ratio_[1]:.1f}%)')
a.legend(fontsize=7, frameon=True, framealpha=0.9, loc='upper right'); plt.tight_layout();
plt.savefig(f'{F}/fig2_pca.png'); plt.close()
print('PCA var', Z.explained_variance_ratio_[:2])
J = '../results/classical_and_sensitivity.json'; C = '../results/cnn_nested.json'
if os.path.exists(J):
    R = json.load(open(J)); yA = np.array(R['y_A'])
    # Fig 3 ROC
    fig, a = plt.subplots(figsize=(3.6, 3.3))
    models = dict(R['main']);
    if os.path.exists(C): models['1D-CNN'] = dict(oof=json.load(open(C))['oof'])
    for nm, d in models.items():
        s = np.array(d['oof']); fpr, tpr, _ = roc_curve(yA, s); a.plot(fpr, tpr, lw=1.2, label=f'{nm} (AUC
{roc_auc_score(yA, s):.3f})')
    a.plot([0, 1], [0, 1], ls='--', c='grey', lw=0.8); a.set_xlabel('False-positive rate'); a.set_ylabel('True-
positive rate')
    a.legend(fontsize=7, frameon=False, loc='lower right'); plt.tight_layout();
plt.savefig(f'{F}/fig3_roc_nested.png'); plt.close()
    if 'label' in R:
        L = R['label']; names = {'A': 'A: ACK as\nnon-cancer', 'B': 'B: ACK\nexcluded', 'C': 'C: ACK as\npositive',
'D': 'D: three\nclasses',
                                 'BEN_vs_ACK': 'Benign\nvs ACK', 'ACK_vs_MAL': 'ACK vs\nmalignant'}
        keys = list(names); x = np.arange(len(keys)); w = 0.38
        fig, a = plt.subplots(figsize=(7.0, 2.8))
        for i, (m, col) in enumerate([('f1', '#4c72b0'), ('auc', '#dd8452')]):
            a.bar(x + (i - 0.5) * w, [L[k]['mean'][m] for k in keys], w, yerr=[L[k]['sd'][m] for k in keys],
color=col, capsize=2,
                  label='Macro-F1' if m == 'f1' else 'ROC-AUC (macro OvR for D)')
        a.axvline(3.5, c='grey', lw=0.6, ls=':'); a.set_xticks(x); a.set_xticklabels(keys and [names[k] for k in
keys], fontsize=8)
        a.set_ylim(0.5, 0.95); a.set_ylabel('Score (axis starts at 0.5)'); a.legend(fontsize=7, frameon=False,
ncol=2, loc='upper left')
        plt.tight_layout(); plt.savefig(f'{F}/fig4_ack_labeling.png'); plt.close()
    if 'occlusion' in R:
        S = R['stability']; imps = np.array(S['imps']); imps = imps / imps.max(1, keepdims=True)
        fig, ax = plt.subplots(1, 2, figsize=(7.2, 2.9), gridspec_kw=dict(width_ratios=[1.3, 1]))
        for v in imps[:10]: ax[0].plot(wl, v, c='grey', lw=0.5, alpha=0.6)
        ax[0].plot(wl, imps.mean(0), c='k', lw=1.5, label='Mean of 60 resamples'); ax[0].set_xlabel('Wavelength
(nm)')
        ax[0].set_ylabel('|coefficient| (scaled to max = 1)'); ax[0].set_title('(a) Bootstrap band importance',
```

```
loc='left', fontsize=9); ax[0].legend(fontsize=7, frameon=False)
        D = R['occlusion']['delta']; PA = json.load(open('../results/post_analysis.json')); ks = [5, 10, 20]; x = 
np.arange(3); w = 0.26
        for i, (kind, col, lab) in enumerate([('top', '#c44e52', 'Top-ranked'), ('rand', '#8c8c8c', 'Random'), 
('contig', '#55a868', 'Contiguous')]):
            sd = [np.array(PA['occlusion_perfold'][f'{kind}{k}'])[:, 0].std() for k in ks] if kind != 'rand' else None
            ax[1].bar(x + (i - 1) * w, [D[f'{kind}{k}'][0] for k in ks], w, yerr=sd, capsize=2, color=col, label=lab)
        ax[1].axhline(0, c='k', lw=0.6); ax[1].set_xticks(x); ax[1].set_xticklabels([f'k = {k}' for k in ks]); 
ax[1].set_ylabel('Δ macro-F1 vs all 125 bands\n(mean ± SD over 5 folds)')
        ax[1].set_title('(b) Band occlusion', loc='left', fontsize=9); ax[1].legend(fontsize=7, frameon=False)
        plt.tight_layout(); plt.savefig(f'{F}/fig5_band_stability.png'); plt.close()
print('figures done')
```

### S7. results/classical_and_sensitivity.log


main SVM {'f1': np.float64(0.7584), 'acc': np.float64(0.7873), 'bacc': np.float64(0.7642), 'auc': np.float64(0.8417)} ["{'C': 1, 'gamma': 0.001}", "{'C': 50, 'gamma': 0.001}", "{'C': 50, 'gamma': 0.001}", "{'C': 50, 'gamma': 'scale'}", "{'C': 50, 'gamma': 0.001}"]
main LogReg {'f1': np.float64(0.752), 'acc': np.float64(0.7764), 'bacc': np.float64(0.7651), 'auc': np.float64(0.8343)} ["{'C': 0.01}", "{'C': 0.1}", "{'C': 1}", "{'C': 0.01}", "{'C': 1}"]
main RF {'f1': np.float64(0.7394), 'acc': np.float64(0.7759), 'bacc': np.float64(0.7385), 'auc': np.float64(0.8223)} ["{'max_depth': 3}", "{'max_depth': 5}", "{'max_depth': 8}", "{'max_depth': 3}", "{'max_depth': None}"]
main PLS-DA {'f1': np.float64(0.7019), 'acc': np.float64(0.7807), 'bacc': np.float64(0.6836), 'auc': np.float64(0.8101)} ["{'n': 4}", "{'n': 4}", "{'n': 6}", "{'n': 4}", "{'n': 4}"]
repeat SVM {'f1': (np.float64(0.7642), np.float64(0.0051)), 'acc': (np.float64(0.7917), np.float64(0.0045)), 'bacc': (np.float64(0.7717), np.float64(0.0056)), 'auc': (np.float64(0.8422), np.float64(0.0062)), 'sens': (np.float64(0.7182), np.float64(0.0114)), 'spec': (np.float64(0.8252), np.float64(0.0058))}
repeat LogReg {'f1': (np.float64(0.7553), np.float64(0.0129)), 'acc': (np.float64(0.7799), np.float64(0.0114)), 'bacc': (np.float64(0.7685), np.float64(0.014)), 'auc': (np.float64(0.8398), np.float64(0.0107)), 'sens': (np.float64(0.7379), np.float64(0.0217)), 'spec': (np.float64(0.7991), np.float64(0.0082))}
six {'f1': 0.39453775302164895, 'acc': 0.5564142194744977, 'bacc': 0.43141031303727734, 'auc': 0.8009147686941093} [0.6959459459459459, 0.5283018867924528, 0.2631578947368421, 0.3333333333333333, 0.36585365853658536, 0.40186915887850466]
label A {'n': 647, 'patients': 319, 'mean': {'f1': 0.764234302533626, 'bacc': 0.7717259131052234, 'auc': 0.8421614964718414}, 'sd': {'f1': 0.00511078409843758, 'bacc': 0.005603230323562229, 'auc': 0.00618127965293236}}
label B {'n': 351, 'patients': 225, 'mean': {'f1': 0.7425110499044874, 'bacc': 0.747400479297031, 'auc': 0.813400346158967}, 'sd': {'f1': 0.013322632510401366, 'bacc': 0.01370883071353827, 'auc': 0.00958943947891749}}
label C {'n': 647, 'patients': 319, 'mean': {'f1': 0.6767281244852306, 'bacc': 0.7096666305584141, 'auc': 0.7661620538374045}, 'sd': {'f1': 0.010874735318317947, 'bacc': 0.011838498351777079, 'auc': 0.012187347942112242}}
label D {'n': 647, 'patients': 319, 'mean': {'f1': 0.656077419987241, 'bacc': 0.6610238317134869, 'auc': 0.8227227637133655}, 'sd': {'f1': 0.008928594237647799, 'bacc': 0.009643172336541456, 'auc': 0.003959866580338907}}
label BEN_vs_ACK {'n': 444, 'patients': 186, 'mean': {'f1': 0.7290643939171952, 'bacc': 0.7385135135135135, 'auc': 0.7939508765522278}, 'sd': {'f1': 0.019679596988634057, 'bacc': 0.0206440758343949, 'auc': 0.019307427947047748}}
label ACK_vs_MAL {'n': 499, 'patients': 250, 'mean': {'f1': 0.8021852149643731, 'bacc': 0.804871188922913, 'auc': 0.8761632938357078}, 'sd': {'f1': 0.009983308634823725, 'bacc': 0.010687343236279104, 'auc': 0.006856390962880098}}
stability {'jaccard5': 0.24087301587301585, 'jaccard10': 0.3350215750095059, 'spearman': 0.512392782941498, 'top1': {'1032.0': 24, '1050.6': 12, '988.6': 6, '908.1': 4, '926.7': 4, '994.8': 2, '1038.2': 2, '1044.4': 2, '914.3': 1, '939.1': 1, '951.5': 1, '1570.9': 1}, 'top1_908_1051': 59}
occlusion {'base_f1': 0.7766933066847855, 'base_auc': 0.8478519209637033, 'delta': {'top5': [-0.0005453120014990986, -0.004585163172284856], 'top10': [-0.015565614025676022, -0.011140773310169938], 'top20': [-0.00924458537263051, -0.01634755642303365], 'rand5': [0.002723984170910909, -0.00015021326513864341], 'rand10': [0.0026779487032012605, -0.001615620732906664], 'rand20': [0.0014235076059639917, -0.0016261249575864056], 'contig5': [-0.00037069255009891666, 0.0023403804292678743], 'contig10': [-0.008650060745846733, -0.003141427701874866], 'contig20': [-0.02774203097493244, -0.016827280685573376]}}
done


### S8. results/cnn_nested.log


fold 1 cfg 4 {'f1': 0.7708, 'acc': 0.8182, 'bacc': 0.7708, 'auc': 0.8602}
fold 2 cfg 4 {'f1': 0.6839, 'acc': 0.7594, 'bacc': 0.6806, 'auc': 0.812}
fold 3 cfg 4 {'f1': 0.727, 'acc': 0.7578, 'bacc': 0.7316, 'auc': 0.8475}
fold 4 cfg 4 {'f1': 0.6825, 'acc': 0.708, 'bacc': 0.6801, 'auc': 0.725}
fold 5 cfg 4 {'f1': 0.7887, 'acc': 0.8077, 'bacc': 0.7873, 'auc': 0.8243}
CNN {'f1': (np.float64(0.7306), np.float64(0.0436)), 'acc': (np.float64(0.7702), np.float64(0.0396)), 'bacc': (np.float64(0.7301), np.float64(0.0445)), 'auc': (np.float64(0.8138), np.float64(0.0475))}


### S9. results/classical_and_sensitivity.json


{"dataset": {"n_spectra": 647, "n_patients": 319, "n_cancer": 203, "majority_f1": 0.4069660861594867, "majority_acc": 0.6862442040185471}, "main": {"SVM": {"folds": [{"f1": 0.763896532746285, "acc": 0.8111888111888111, "bacc": 0.766025641025641, "auc": 0.8427021696252466}, {"f1": 0.8235951134380454, "acc": 0.8571428571428571, "bacc": 0.8387755102040817, "auc": 0.9244897959183673}, {"f1": 0.7373461012311902, "acc": 0.765625, "bacc": 0.743762265208859, "auc": 0.8458088029156153}, {"f1": 0.707717980809345, "acc": 0.7256637168141593, "bacc": 0.7087525150905433, "auc": 0.7706237424547284}, {"f1": 0.7594896331738437, "acc": 0.7769230769230769, "bacc": 0.7634575569358177, "auc": 0.8250517598343685}], "chosen": ["{'C': 1, 'gamma': 0.001}", "{'C': 50, 'gamma': 0.001}", "{'C': 50, 'gamma': 0.001}", "{'C': 50, 'gamma': 'scale'}", "{'C': 50, 'gamma': 0.001}"], "pooled": {"f1": 0.7602920344339108, "acc": 0.7897990726429676, "bacc": 0.7652942351218213, "auc": 0.8434518262104469, "sens": 0.6995073891625616, "spec": 0.831081081081081}, "oof": [0.8396384298829547,

0.47187841786562523, 0.7015497906623778, 0.6610778661713033, 0.863066082511234, 0.252505811512558,
0.040900427467257014, 0.03016709810778117, 0.12410048354033971, 0.07714878555896297, 0.20035808978165573,
0.2002240436589161, 0.07688956485941617, 0.06536004533907494, 0.028326311139444022, 0.9175358658616072,
0.9898407599074648, 0.5946057618662117, 0.9181745980683678, 0.7545749849954104, 0.5636943040911496,
0.13301661795733308, 0.9238522537292555, 0.8922740060743911, 0.19275543896350647, 0.15406240934705304,
0.08332985841040859, 0.14231655526022366, 0.21040520280431416, 0.23435388081907094, 0.9003390255817294,
0.7111606280982901, 0.23276435705653498, 0.4087699241449856, 0.7710197324673919, 0.9608245100247138,
0.7288919902852367, 0.13222132236343861, 0.14554811732206502, 0.37811576932340535, 0.14595695240729964,
0.0636930956796917, 0.17631547492292232, 0.5552862333273358, 0.2710745299679263, 0.7277824996604654,
0.36214890997532795, 0.7594000652846887, 0.17891364554669362, 0.1983896210495454, 0.19394716584541183,
0.3126887777187432, 0.16699587981239952, 0.36561218491643044, 0.16222864490395167, 0.7791733416805326,
0.09544247401937471, 0.07054364165116786, 0.14354557515469796, 0.08072594321919833, 0.0830187871852787,
0.06625262803417875, 0.05645535447628526, 0.06226759570508152, 0.11960997804958921, 0.16938586841110417, 0.5,
0.6923741019783373, 0.36509899669731727, 0.18581238811733347, 0.5274733283921758, 0.3306246019028128,
0.28576736577493245, 0.9857198206311394, 0.9199091258600479, 0.6910993156572085, 0.10393595545734147,
0.212803877279989, 0.10061200799324979, 0.06076250647589689, 0.1431754976655963, 0.06581777252212392,
0.1620984568846493, 0.06631545407917681, 0.1021702289481436, 0.04739184375122265, 0.16465255927274938,
0.07519484331561226, 0.14918760341192663, 0.07464276117251763, 0.13812721022203064, 0.19362195554490721,
0.34245649820617863, 0.07351114030257534, 0.11730395579649469, 0.7931236628160796, 0.2877811853242814,
0.007347481301860349, 0.1494243804523129, 0.23568671162996552, 0.12119138395991784, 0.15793225100621414, 0.5,
0.16312441565059882, 0.19325811766004874, 0.9875762921164634, 0.4657560726955055, 0.8743720523381376,
0.18912278058540324, 0.30403031132625813, 0.043418308620948025, 0.06548067721308438, 0.06136578405994741,
0.05267435051217814, 0.30368657721193, 0.1140453670707304, 0.19837807879211075, 0.20501554776272246,
0.2775590718906603, 0.15612228062294545, 0.34636789443513144, 0.39551985785489835, 0.13659675732779414,
0.035047211330372634, 0.032999568021394865, 0.05476933239528317, 0.2800310771700616, 0.14576914916691047,
0.09495082349269647, 0.11009506540153052, 0.027100213660299743, 0.060534327806823766, 0.024593659508763206,
0.0320051775781198, 0.01971396177451134, 0.0944304237452291, 0.22669855863470853, 0.10296326780082364,
0.15041360649263835, 0.06145118918127574, 0.11316178906496714, 0.08377858630933467, 0.05389576462158505,
0.2208380431380415, 0.046265097129525305, 0.8349658334764869, 0.9345269712862352, 0.1449824228032662,
0.29731034931672495, 0.46316593808507245, 0.44665944908512495, 0.08718561534289834, 0.15660794001781975,
0.07315912149734674, 0.050298838075060956, 0.05896269810732854, 0.058929148196758416, 0.059863019081955716,
0.050629541712216966, 0.6309197250887163, 0.5364388902645458, 0.35326341965367697, 0.24932156768211056,
0.9876208120950197, 0.23103061279246878, 0.2203404756722516, 0.7915250049075504, 0.18253045126571613,
0.7930103575799177, 0.27667668221696706, 0.08195048754745368, 0.08782750634409811, 0.05733883076863194,
0.09572229934647869, 0.08140802231417962, 0.301542666351199, 0.060668391050332456, 0.051718936543900446,
0.06426739467018307, 0.10960546317433444, 0.6623403047164402, 0.4549012982085836, 0.6513943016248614,
0.4271376108254227, 0.6547003850578637, 0.12845115906723448, 0.10602307054517109, 0.6910666507822315,
0.6427335663544556, 0.19804745870153037, 0.44813485452758967, 0.8192146244671227, 0.4706125854767678,
0.6502070670215395, 0.3783441739840749, 0.1488887602813687l, 0.43257282212796483, 0.28821110594962884,
0.34121825406738, 0.5841923619507462, 0.27097340200445924, 0.7742374087045293, 0.19430520306212265,
0.5618861429386094, 0.6258193275390811, 0.3789734548014687, 0.13427202441193603, 0.1415130200056748,
0.7267580334566052, 0.3685082461062264, 0.8368163055389344, 0.4829330616079089, 0.518026477849448,
0.23070376224406772, 0.33066664965536985, 0.6493689621302708, 0.3539263897399758, 0.21594010595225782,
0.28260780753437886, 0.10460772649642075, 0.1004250532623251, 0.08764341176608637, 0.08738413686359554,
0.13717104590428716, 0.4217776471123171, 0.26443693009026853, 0.2217558279693035, 0.38596929071521274,
0.8694063835560678, 0.06145272793161744, 0.13741926932288653, 0.03260931703741272, 0.05954951384476554,
0.09743267402738995, 0.06101113115933141, 0.39407521623936254, 0.2864119482420888, 0.5561657321547961,
0.36306458640051026, 0.620078450427865, 0.710759783331754, 0.8685758721732304, 0.6633976190765298,
0.14350976802924884, 0.5593046856195277, 0.02231897673341961, 0.08481639187909676, 0.13104209097040886,
0.10106553517191535, 0.6672898434379648, 0.41752358638626763, 0.2988552661884506, 0.31116619952231783, 0.5,
0.25925522663092043, 0.36059684456128144, 0.12457184768953342, 0.174428966041955, 0.14485754137575418,
0.6382832882598249, 0.40774255564336565, 0.28925173922219405, 0.7657350771981176, 0.886976797967463,
0.3114187441399849, 0.36016482040841885, 0.35676110516820386, 0.9621116501085127, 0.43152517399086626,
0.7035820396552607, 0.3092262422496458, 0.438946155045018, 0.15386460416580008, 0.1558342079612405З,
0.10360077587078383, 0.0886146069150802, 0.08752627348421865, 0.25402133023205253, 0.18659601524880517,
0.2114667753033706, 0.09468658493388508, 0.6210188910335068, 0.0711488894652275, 0.05693709556571033,
0.14255415914959477, 0.47914815330096416, 0.18414564887845042, 0.15857406603320062, 0.18399503657229335,
0.1925721036160737, 0.1140681800038534, 0.27298408801094437, 0.054014693007267654, 0.11989244382806871,
0.09568911401997697, 0.09792517559969692, 0.040814543338229294, 0.0777308850166272, 0.31768955112399394,
0.08598508654992634, 0.12349615931932971, 0.07772611843650983, 0.0943580780189221, 0.14909430013644853,
0.16145926987753934, 0.05162756312477686, 0.11864687712054173, 0.09883650774543985, 0.10000676306073715,
0.5926429199699039, 0.28918573735496217, 0.7871263934593817, 0.025646424506668018, 0.0791923162741846,
0.6555742582813495, 0.33746034480867304, 0.8835715698436928, 0.6599200206466801, 0.7849319039387943,
0.8383794465495967, 0.13495433381008493, 0.9718941715228522, 0.7902388802782578, 0.31794147005401485,
0.6056030749155051, 0.1052073729781586, 0.12125692925321807, 0.9180178572855843, 0.711592724509642,
0.3374245241615569, 0.41603779012671044, 0.3144710573075589, 0.296586681602088, 0.015109833659333287,
0.010231480072731745, 0.006465565213016355, 0.024036475859758787, 0.04632621910530514, 0.024584103262085065,
0.02337120995229028, 0.13248916245704084, 0.07235450949794549, 0.3788051749453546, 0.1430990546452058,
0.2949773863812065, 0.25540939236539345, 0.21895307750950052, 0.3583885992165142, 0.28129023694022687,
0.17072985652944248, 0.9328442256339758, 0.462895292061004, 0.086570356166882, 0.0818993186434544,
0.13627390891111682, 0.061635011712670815, 0.13403135910567704, 0.22633392451858078, 0.3137986894943648,
0.5930596985373091, 0.21741927139166012, 0.22796636003614262, 0.1955746718835602, 0.15921949142740438,
0.26708372470690905, 0.177781317840116, 0.21832530904642888, 0.08018429348793678, 0.19327812059383734,
0.20420243754295253, 0.4943794945989867, 0.25768226995326043, 0.17276136834271244, 0.20941072661738225,
0.1097074180812395, 0.40179002036146744, 0.7805637890295396, 0.850980959598425, 0.7648570687903504,
0.46875040783167865, 0.1030770984050488, 0.017301035164126132, 0.024840518812732904, 0.0418388126850268,
0.012017212847342107, 0.01742313869490186, 0.042779537454045356, 0.6655026024008565, 0.2633735269816897,
0.11119222780282836, 0.19258879368225176, 0.7477719794466022, 0.29685498946368366, 0.8320891285990665,
0.15386682529849371, 0.08200357869474631, 0.08720320667034454, 0.13748598209574453, 0.8654634679516802,
0.09396595691400401, 0.1754808283154714, 0.21233532454826667, 0.09457909920054244, 0.09100107366133951,
0.1344428618768941, 0.6699246844553571, 0.17654658077006316, 0.0581105622510948, 0.4372779282044258,
0.7285385259795197, 0.7849458894959959, 0.009237967854750425, 0.23046444865748997, 0.14156228143827304,
0.1708069851852345, 0.02172862912982993, 0.22881216397266393, 0.3660132481650741, 0.9405947568900602,

0.09421062774489398, 0.04824980157911544, 0.8229530723089927, 0.16059528993145375, 0.22529441297376174,
0.035026259622356504, 0.49234656251418973, 0.3164692846966548, 0.46368091227860603, 0.6052570033379221,
0.2699284990587117, 0.542697221762126, 0.8852636176513605, 0.9614414643233307, 0.4136439344325754,
0.8911721308768183, 0.9634207950055964, 0.1568280260412719, 0.9075085713556313, 0.4753140406727045,
0.7219410664777887, 0.8387020116499125, 0.9808936826737447, 0.12468258593233787, 0.096035983746294,
0.11135771852760966, 0.19192867000294228, 0.09970515050766288, 0.7910234133896931, 0.3168201722927597,
0.21530471666621934, 0.632431932780457, 0.5379861448623341, 0.6329162468893683, 0.14148506937108044,
0.14534483827269448, 0.8627487179159518, 0.07057502247451088, 0.48132688552845626, 0.28636520189156167,
0.19821791094965094, 0.2157161147085745, 0.7881687377857849, 0.16506101744428828, 0.12743875608156535,
0.1443110686100057, 0.15345050234674482, 0.1610094200547617, 0.0653221472220947, 0.20247187782754023,
0.12959636196319313, 0.3299934733579068, 0.9805217259869964, 0.4394499590208274, 0.6681182414691405,
0.9732891492428456, 0.8210044233878806, 0.9913650150162111, 0.07093140814222063, 0.03901209536587504,
0.11096697159991276, 0.060387822181637, 0.14323201099393257, 0.08763301263223469, 0.43402311235184143,
0.48410903208721473, 0.20040089624742694, 0.6465273762032016, 0.23323003648105853, 0.6793387539525235,
0.03901639727380669, 0.02487691300838538, 0.15665753413838293, 0.19945356679027815, 0.18409221861301377,
0.13973636441195156, 0.13806961447044497, 0.09240509027409817, 0.1484467141690071, 0.6990723943978957,
0.1099710058444823, 0.1659160957863614, 0.2145224702410773, 0.26816546426488164, 0.8922553081796646,
0.8863397796689574, 0.6798892215055461, 0.45791664331123905, 0.22278308750387618, 0.7630914524549369,
0.2432167194608791, 0.5936489236775537, 0.4600145023145591, 0.19269991030235295, 0.6514317429468341,
0.1421079014653655, 0.07340698506935954, 0.11819053089963028, 0.04543080730285599, 0.7532337944958818,
0.17416418217595134, 0.18633689808349546, 0.20292187480011317, 0.17288953283066905, 0.06859273713180872,
0.23598802450938391, 0.18386819515725222, 0.14346555352547394, 0.08846924016534112, 0.13796822784198765,
0.14243110582952787, 0.1752717318138703, 0.062261867364928544, 0.12776675885580766, 0.06028284048310502,
0.06336847594026121, 0.08652612461947749, 0.08791253414344706, 0.08965283079679165, 0.11225513319221413,
0.22433220464427903, 0.15173752879897126, 0.09752273472511147, 0.0930170102968142, 0.07998236905567854,
0.07985837627762957, 0.07440828507110576, 0.06432476084392949, 0.30252054196879447, 0.6562385419646837,
0.20434705605309647, 0.3172699745122834, 0.4948957306830577, 0.13705140638187863, 0.15594254404545754,
0.3950204774039437, 0.9045141134466931, 0.5413626195390359, 0.4581031377862982, 0.4158804532316711,
0.3511358019781705, 0.3104166717608022, 0.036227366701668186, 0.708253196293291, 0.11929731366087508,
0.09358925918340126, 0.1729195811563776, 0.8728962968540996, 0.24950427506956693, 0.22492335994682858,
0.8557258666260797, 0.34286651589975786, 0.21203485619561968, 0.21323173025585498, 0.18605030384504526,
0.3422131450562747, 0.7670679270605167, 0.6679192239751258, 0.6552226569224279, 0.07187193926860654,
0.24023483815819968, 0.08603569149224524, 0.5906288534241233, 0.2131672431546088, 0.8490738513644179,
0.47553188595849377, 0.1133481046354559, 0.15283564665491386, 0.6682127288222761, 0.07190641019279816,
0.40195365957377016, 0.1536639176199923, 0.2868016336198992, 0.4853673437207175, 0.14240593119021022,
0.6482372628695123, 0.614988402579155, 0.7629497422694865, 0.06906384022178641, 0.36843220088037143,
0.7033882206202663, 0.5547758794070983, 0.18279736264868354, 0.2126927981080039, 0.06948554462819176,
0.03920249895198849, 0.06630450161847366, 0.12361665621556121, 0.24023432752984397, 0.08989066517411634,
0.1101983221980031, 0.06510128294207744, 0.19168184056287083, 0.08663583362720702, 0.1298700385435975,
0.6467618774084931, 0.38269088312172667, 0.05944050794107446, 0.34885501294014415, 0.4659889475192284,
0.23380380518159463, 0.4076209308209966, 0.37204946359157115, 0.8351417669808243, 0.4248105419879985,
0.19901679833311778, 0.6261270086962711, 0.9805773124040648, 0.3566059837107282, 0.5848573032952319,
0.1530689102704907, 0.42888311761017806, 0.10532755826347417, 0.19272929768320116, 0.4527959410305232,
0.22869574009829952, 0.32259511182444034, 0.3834297760210957, 0.2630197783937779, 0.5340782384012781,
0.2847415181207477, 0.426958124487361, 0.5620438258630406, 0.10871672695175821, 0.44489993514457693,
0.2837255946617071, 0.13530089468231618, 0.4064331320440716]}, "LogReg": {"folds": [{"f1": 0.7173255813953487,
"acc": 0.7622377622377622, "bacc": 0.7323717948717949, "auc": 0.8318540433925048}, {"f1": 0.8710526315789473,
"acc": 0.8947368421052632, "bacc": 0.8918367346938776, "auc": 0.9571428571428571}, {"f1": 0.7527805527805528,
"acc": 0.7734375, "bacc": 0.7688533781889544, "auc": 0.847771236333053}, {"f1": 0.6700041718815186, "acc":
0.6902654867256637, "bacc": 0.6708584842387659, "auc": 0.6723675385647218}, {"f1": 0.7490503767357868, "acc":
0.7615384615384615, "bacc": 0.7613871635610766, "auc": 0.8625776397515528}], "chosen": ["{'C': 0.01}", "{'C':
0.1}", "{'C': 1}", "{'C': 0.01}", "{'C': 1}"], "pooled": {"f1": 0.753504109552545, "acc": 0.7789799072642968,
"bacc": 0.7654329206053344, "auc": 0.8342098255891358, "sens": 0.729064039408867, "spec": 0.8018018018018018},
"oof": [0.967072166518199, 0.6451523713254171, 0.40367308636178095, 0.6457162428178786, 0.9806210873111133,
0.47146221549443046, 0.1969704917595437, 0.12537979166055951, 0.28182200985626793, 0.22133063119102406,
0.08774067009246003, 0.19072723789189316, 0.14550006011335095, 0.17544233794737274, 0.22594116049525886,
0.9590771887364629, 0.9946251838898048, 0.5875173266051719, 0.9076288997897088, 0.8844381577437943,
0.3965262230702327, 0.21009529549233466, 0.9753653625278355, 0.869947848649586, 0.28523018349349166,
0.6625862883103765, 0.8376342991087998, 0.17414410376263778, 0.1722346043862102, 0.7105555839232279,
0.9246703302824464, 0.7040580145103648, 0.5795751685182523, 0.7192872148067028, 0.7302291999307687,
0.9819885294710592, 0.9151363684826558, 0.6267859013676327, 0.1937770544410913, 0.39540581291903776,
0.30376572007161246, 0.15374174288621992, 0.4202766533125227, 0.6835682442885631, 0.4292176157391973,
0.7832770313257967, 0.5731977983495692, 0.8141886995915067, 0.4107592571142017, 0.41192970535846907,
0.35413485370959746, 0.13631385535537238, 0.3784926195144577, 0.4798505925444994, 0.16943024003362125,
0.4201955681001231, 0.2569976046761316, 0.18599724367895243, 0.3241155030260259, 0.21524135199010588,
0.24442574607707168, 0.20416076438576158, 0.17564754128871649, 0.22419550707738822, 0.25316749358582613,
0.14580074737297244, 0.513495586733768, 0.5345907646608266, 0.8468997208321519, 0.22897988562341487,
0.6784799498860692, 0.598543666335499, 0.6380547741307924, 0.9971695786599916, 0.9915315151560307,
0.9254779878201057, 0.15999264273017008, 0.338670948629788, 0.2696910299645728, 0.22410564457979798,
0.37579222753771296, 0.2196754182168542, 0.3838938479421375, 0.21763325819756807, 0.28810381920296046,
0.1799005671027999, 0.4001402375588006, 0.2544262755821856, 0.29105453763396766, 0.23628830086463357,
0.3424039682513497, 0.2663961684514493, 0.3786251370930473, 0.20736556442405127, 0.3540352889932819,
0.5280262659424814, 0.3531870219802889, 0.22719747231696408, 0.244214509590662, 0.33186328528375125,
0.15773147733169818, 0.17053257007512793, 0.6057689685079365, 0.20768128250024237, 0.27157803593442,
0.988466943249614, 0.8904257059427205, 0.8532160690399355, 0.22232354848539174, 0.5556614076554817,
0.07640323474184249, 0.057942692979872215, 0.06420259227264324, 0.12032933120399535, 0.13253034227704275,
0.16897728988672242, 0.5631870606132998, 0.49735911314765946, 0.4365853048169579, 0.3207776098986785,
0.6659019653936263, 0.3810970785632269, 0.2548656812471236, 0.5107777282259139, 0.022644804426813582,
0.04824116700835014, 0.4670931397914575, 0.14433557300849342, 0.09463544748145707, 0.1179795512321292,
0.02643494195198137, 0.0925547804717006, 0.06459138410005597, 0.06630698826852462, 0.0433892362669665,
0.1691862272165568, 0.5182294440170111, 0.13679677514329525, 0.2838661112574114, 0.1310119214235005,
0.1434299953447992, 0.07838564299097514, 0.10864062393843993, 0.35515206126447135, 0.07566556264829281,
0.9877952205692629, 0.9921283051930856, 0.27237781205720535, 0.5057814519538079, 0.018515925981712367,

0.6069211293320806, 0.5333518987598989, 0.2637367043205739, 0.18342178923743446, 0.15651262242095992,
0.12854091531335354, 0.1226520754987577, 0.10414333048748754, 0.09730201436706104, 0.8840910952088362,
0.6710077874164662, 0.515791301549348, 0.20051516577724493, 0.9791380066364282, 0.32578968847447737,
0.5139912479901741, 0.9100588652056283, 0.8154211228113442, 0.8186766835262076, 0.0982174907366553,
0.08079588867445356, 0.112135081251312, 0.04680057003591932, 0.1043736505340472, 0.06088687108325312,
0.2809921992822716, 0.053000398668697594, 0.0477492274084705, 0.08845783249986645, 0.08549539197484543,
0.9784355836830594, 0.9815992817785985, 0.6232908691242287, 0.10123121876159881, 0.6554064015553507,
0.1772620181198677, 0.27873954165239156, 0.891354942092520, 0.7064628241169455, 0.3364352010702175,
0.5516687923858877, 0.7771137397958151, 0.7346857246728251, 0.7518279113428047, 0.49967465852079734,
0.37753677257889856, 0.18532115157317156, 0.5118486786542848, 0.4743665359651573, 0.6586076477550125,
0.39808386108479427, 0.8914963557558604, 0.4097576052779511, 0.3928025406714313, 0.4669227660479392,
0.6116252474381298, 0.17541134572750497, 0.1509341985935428, 0.7915115656401116, 0.5937845374851473,
0.7953941443856807, 0.5631245416365844, 0.9619618216160833, 0.5993452962051322, 0.4035946671951817,
0.7639312453170382, 0.4417512227957953, 0.4574651418991164, 0.5047825029187808, 0.1167399269887547,
0.16401392506650336, 0.2249011373241205, 0.31602008534678067, 0.2956034065039028, 0.9664782502868781,
0.40736408037418237, 0.44594430004686747, 0.5495245189634096, 0.8814614517147679, 0.13864849279114846,
0.16460794500467918, 0.09505578352216006, 0.16998900823220034, 0.1811040024785720, 0.39399552277539884,
0.5596830877285706, 0.6029198463705917, 0.7396983870129317, 0.1722266031280128, 0.6052123310275787,
0.6948056360083167, 0.7935156146648596, 0.6444339442173008, 0.5399462180353113, 0.4881338236476833,
0.04632614305109729, 0.33452686009312477, 0.29471243627604615, 0.2638711628933961, 0.7261872079494762,
0.4031565114623203, 0.6045707655208331, 0.6487872210258777, 0.7428271711496368, 0.6794377616595576,
0.6347812202972074, 0.6224355089549981, 0.25783925168064314, 0.3221590338671206, 0.6237829733319297,
0.5230854267842011, 0.7789279962483583, 0.9935306090862049, 0.9999667481435323, 0.5027734401353102,
0.5553948470452578, 0.2590180181986547, 0.8980170371806945, 0.613936198649963, 0.8260733412345957,
0.6512252472178297, 0.55475323492729, 0.2670404683648905, 0.2565923334515422, 0.2150555560529501,
0.19102589297360578, 0.20126428358096154, 0.32500407191446506, 0.28178766524598614, 0.27173536922652514,
0.16679043912816566, 0.48708537894317316, 0.14038005055650946, 0.13142204603692384, 0.2375569316763641,
0.48508658752837575, 0.2728301085277853, 0.24975569007126808, 0.15040076512625308, 0.13484777228407946,
0.15320971336875008, 0.30237718553562676, 0.08583091536716886, 0.3744773902204991, 0.15090584363661946,
0.18682141583031095, 0.1548871369245178, 0.2074497963083886, 0.4383773338597651, 0.1995524510014092,
0.2627647467668692, 0.4706812658598592, 0.1501110709752036, 0.1814993389201577, 0.2815928808783798,
0.3056782314688974, 0.32357043748477926, 0.1664835257355323, 0.17890584618674055, 0.5205527176572284,
0.25024392351842684, 0.9586444975026261, 0.35936731926148824, 0.1914968356278352, 0.621097096078725,
0.38326933437334687, 0.9317908544035172, 0.4874352827361682, 0.7576209488838359, 0.8463692896883739,
0.21929460821730304, 0.9999579332676272, 0.6340262131236583, 0.3923907942960731, 0.0174622341375777,
0.17491991573133617, 0.3177712014234026, 0.8880063901820485, 0.668567381033678, 0.5264483525440502,
0.5597233576032387, 0.5615991181252101, 0.071224581586858, 0.4497584430168628, 0.19670339508089976,
0.3190409141808095, 0.22678318827419539, 0.3945563009721759, 0.39747165711303384, 0.28263369968109786,
0.14788628497115272, 0.49812877898000985, 0.569302128164249, 0.3467875852283161, 0.192680964572038,
0.41662402380855706, 0.7208521921630532, 0.31185493186232505, 0.7146689070963397, 0.2923440688014972,
0.748526954504169, 0.5688098793688691, 0.1280556500528004, 0.21653160507036423, 0.2977034651106149,
0.17583130272703057, 0.2975985018638128, 0.3798403850233947, 0.413813269422577, 0.8764815440769514,
0.5593892486312245, 0.510490128000524, 0.1941460597671671, 0.3740136969049969, 0.4599214942339393,
0.3270516168935371, 0.38999344857737506, 0.213113667872311, 0.3646370990427113, 0.4091578106530865,
0.25073097861467847, 0.4835245226360881, 0.8599447725501657, 0.8480947925370184, 0.41495849900223764,
0.5902887339363816, 0.9642372289532684, 0.8268543081060143, 0.7253387617610288, 0.6718209016841183,
0.18458001630399748, 0.021058889482088925, 0.019142608452660703, 0.0334893595631652, 0.010062231356872411,
0.019309695640563524, 0.028262952650232393, 0.5719825328394058, 0.546937002080746, 0.05402566180179926,
0.1591378696616156, 0.6201391748719844, 0.9052283870438902, 0.8902671272271765, 0.2657347497891681,
0.16757022229809843, 0.15220699358845516, 0.3365983637341763, 0.9991488674011639, 0.14518701205567835,
0.361117346003421, 0.4071487885639652, 0.10559001794010138, 0.10736365888230089, 0.17848915448391398,
0.535883958404039, 0.4438348318856942, 0.22351206115937655, 0.6744029213673631, 0.8014623841155291,
0.7450641654728374, 0.2829300991962007, 0.19700150438848732, 0.20650218892036692, 0.37480342656692867,
0.5433924493264245, 0.315052798696652, 0.7322147692343557, 0.9902711011815282, 0.20089450605645892,
0.12461113212038724, 0.6893143769999215, 0.2609781072669761, 0.4145214319049231, 0.0556349432062465,
0.5135607907371281, 0.17008968862211102, 0.22504084465819163, 0.2976755494039528, 0.5226097361748361,
0.437813694969689, 0.9224120239844045, 0.9997005541504642, 0.4880805308504326, 0.7245500207824053,
0.8470474911040172, 0.26482598482378655, 0.8568047203112097, 0.6025800867261056, 0.7701070467311208,
0.7873986102939853, 0.9588717621386473, 0.155120831219169, 0.07301238607717382, 0.27162160199775637,
0.27192996496652355, 0.28679985906200856, 0.8927983052680989, 0.45823359559020566, 0.5228573833074555,
0.6680288622730657, 0.7320933059938745, 0.8381911380271811, 0.18465292067316355, 0.15778122326486185,
0.797487106498929, 0.15273282517887746, 0.6264175228633866, 0.10334049340926586, 0.353804575500107,
0.2830605557440595, 0.6817222807047977, 0.28227278645696896, 0.5001380974749462, 0.3670663547577099,
0.21239976286796047, 0.25612941928045835, 0.5196734349122573, 0.9987598648163141, 0.3212630655048515,
0.6514404174603833, 0.9321623969892014, 0.7830646395832945, 0.9998450167692631, 0.9974228865381858,
0.9989412569573073, 0.9965183955987428, 0.15954344691204184, 0.11058793010525554, 0.13989066641418638,
0.19119393596698622, 0.2543838719426428, 0.20164689693350638, 0.2607167665289948, 0.3401567767450149,
0.23542187028719033, 0.32309438245909167, 0.3127524123248985, 0.7235519173647057, 0.15378564964110966,
0.18148588588986922, 0.2512218388525661, 0.37193458652239864, 0.356529336364946, 0.27513425997804647,
0.25891816982727334, 0.0967932435371529, 0.23647940603902542, 0.8178054563597886, 0.2522871213194207,
0.33600696039749317, 0.38968144253168413, 0.4193832242613742, 0.2818364344076388, 0.27950045297968434,
0.3637487065967279, 0.5773149873739841, 0.42102740781119724, 0.5060452498064415, 0.4965630516825888,
0.6787694533317701, 0.5339453188633848, 0.3576156595995224, 0.6585604019092633, 0.3231845685166016,
0.09190973834006094, 0.21021845240296094, 0.15084533165843253, 0.9607962737955756, 0.13170910303724453,
0.24649824015270425, 0.2988079389411818, 0.11086804719291589, 0.17293809086085535, 0.4187613586627494,
0.3506545502036438, 0.355694558034707, 0.24615409530415253, 0.32705211667415385, 0.33347741506867934,
0.3898705204100381, 0.18172875134861857, 0.31850703863374, 0.1517075014182979, 0.18616359176054834,
0.21844618514119232, 0.21484093558246806, 0.23117366315052323, 0.27326820792390644, 0.4252112061162674,
0.3023529021418751, 0.26077314004183894, 0.2237559262573588, 0.21387600151380182, 0.1828360422279722,
0.22070378993866127, 0.22192963222161918, 0.4946667673081407, 0.9973372294045417, 0.38140881896076534,
0.558902899203899, 0.5116290703057977, 0.10476281086881972, 0.2796779731657345, 0.30497048630406726,
0.9777265603251974, 0.7398964390895143, 0.7535208316944474, 0.5776032925427821, 0.5604297681101371,
0.40014245977655294, 0.13568990512112017, 0.8474356505987798, 0.5350259463245789, 0.14888948732288426,

0.1690139192211322, 0.8138086891771888, 0.3612206308614419, 0.32905533870077786, 0.9537567997026865,
0.5542563785301876, 0.2875854767626283, 0.3452181312572437, 0.24146924722883728, 0.2942573707260667,
0.6343866928762993, 0.5823833466860382, 0.4047741876458635, 0.13602460300238298, 0.33413875306799246,
0.17713452762805948, 0.46463672250757493, 0.26029090211465467, 0.9868308736140038, 0.5582528859195559,
0.1449025223082616, 0.3951573083403768, 0.9238235657076134, 0.9999816520415891, 0.5996284675409775,
0.9538394080828086, 0.5752365654927583, 0.5792786983055738, 0.23762083714600224, 0.9204866412975112,
0.5933843426220098, 0.6407894163609709, 0.20855108383756865, 0.4174508446209988, 0.8823030292107356,
0.6948805666603302, 0.19800883454914292, 0.21614176471571292, 0.2068018898072794, 0.132273001975317,
0.18038216871440563, 0.2770575708492421, 0.43994562171568696, 0.2152354972574608, 0.24503212219686493,
0.17467776521133577, 0.44869155602584965, 0.23806054825032077, 0.07638857538120 89, 0.6594023622618629,
0.9813073175741027, 0.0774959791678754, 0.5779745587100262, 0.6124359484425758, 0.4623869632089722,
0.620346984365552, 0.9345488543319549, 0.843618842773547, 0.6118651181099424, 0.3861698577644804,
0.7336915913692308, 0.9925659891660389, 0.004907848165068732, 0.2785937274424509, 0.19115787967975262,
0.8468782251189912, 0.21640532841839083, 0.5658352649007091, 0.6918453984334358, 0.4251238013742262,
0.5223321257635374, 0.521829047153153, 0.5610787183582291, 0.8029156730293889, 0.2879199987840612,
0.3774911930243602, 0.9872642189595996, 0.19137673024131652, 0.4711845928893816, 0.2952515773454093,
0.09745493062601396, 0.5037114472975504]}, "RF": {"folds": [{"f1": 0.7560189573459716, "acc":
0.8111888111888111, "bacc": 0.75, "auc": 0.8153353057199211}, {"f1": 0.7905211912943871, "acc":
0.8345864661654135, "bacc": 0.7959183673469388, "auc": 0.9169096209912536}, {"f1": 0.7308662741799832, "acc":
0.765625, "bacc": 0.7308662741799832, "auc": 0.8446874123913652}, {"f1": 0.6746273712737128, "acc":
0.6991150442477876, "bacc": 0.6730382293762576, "auc": 0.6961770623742454}, {"f1": 0.7450980392156863, "acc":
0.7692307692307693, "bacc": 0.7427536231884058, "auc": 0.8385093167701865}], "chosen": ["{'max_depth': 3}",
"{'max_depth': 5}", "{'max_depth': 8}", "{'max_depth': 3}", "{'max_depth': None}"], "pooled": {"f1":
0.7416285713487837, "acc": 0.7789799072642968, "bacc": 0.7400312874450805, "auc": 0.8229596591665557, "sens":
0.6354679802955665, "spec": 0.8445945945945946}, "oof": [0.82, 0.885, 0.5495661519043546, 0.40509319186615,
0.6411449189317584, 0.45214852334269584, 0.1539634095841404, 0.15609054230902492, 0.2640652367767487,
0.17733990261264648, 0.19299075115363237, 0.38972727304979665, 0.257399834550024, 0.3275496082852191, 0.15,
0.7380154130185989, 0.9843503172825682, 0.96, 0.7791805436555375, 0.7476891901884851, 0.6002260816044315,
0.20606309348610308, 0.7249668337046334, 0.904653215143249, 0.3198937165910227, 0.5558512510880931,
0.4162823287823288, 0.13692894956933466, 0.2786034819600317, 0.6304904033871235, 0.9415093699761727,
0.964613475177305, 0.4629828587155901, 0.9675833065881302, 0.5999397999307657, 0.9451792286945203,
0.8513846586624599, 0.26765280988489665, 0.23720174035556404, 0.89, 0.2, 0.06, 0.19844808728663085,
0.6826289128225678, 0.39503020457953464, 0.8913992769828218, 0.43243612670010423, 0.9254052985313926,
0.310585055983811, 0.45222854048159855, 0.37232433449201663, 0.18016127396269813, 0.3537826403895782,
0.38062772062627515, 0.32647319883915477, 0.40484449242904275, 0.2092075576657408, 0.14709918540616837,
0.20018455197976048, 0.3898861362196363, 0.16922156719848463, 0.25154349308422447, 0.25937041965572805,
0.2662065685636597, 0.235, 0.12, 0.6143341028377396, 0.6269756476024955, 0.6519806581922829,
0.3202084529673045, 0.5227376626778368, 0.375, 0.42, 0.95, 0.93, 0.875, 0.22460563112809964,
0.22214719121796833, 0.1242546388338041, 0.16671086098630827, 0.2642508217937602, 0.1417799645155147,
0.2990919216283881, 0.1964769218462025, 0.33020803057524284, 0.15348559312088608, 0.23572252617756284,
0.10436300275329222, 0.45396841845493646, 0.20928969757195218, 0.40325487842074287, 0.45304096128661775,
0.2887458667026177, 0.16502254650764442, 0.26711148412084584, 0.786044791076089, 0.24824621758803755,
0.28557492865912076, 0.2462580460908515, 0.19864928403594795, 0.2560659692322341, 0.26972417329440235,
0.5777379116810255, 0.1870346423487236, 0.30405442647202857, 0.9393839631128933, 0.475, 0.929205952442107,
0.195, 0.595, 0.02, 0.105, 0.03, 0.025, 0.4375456469476916, 0.16556639682402477, 0.15573054805377703,
0.2867247132259823, 0.2451151274407096, 0.3608814134478382, 0.7070349827375764, 0.345, 0.21983499523159306,
0.24126825910763577, 0.005, 0.05, 0.365, 0.14, 0.06, 0.03, 0.095, 0.04, 0.05, 0.0, 0.025, 0.305, 0.42, 0.04,
0.42, 0.01, 0.61, 0.26, 0.015, 0.47, 0.01, 0.8911309523809524, 0.949113475177305, 0.1, 0.4656442187846018,
0.3965107915762245, 0.3145818669334527, 0.290175949683245, 0.24212141263629394, 0.17980717730544488,
0.08152995246867434, 0.1887343180242415, 0.13006952872965188, 0.08068666232956939, 0.13859929036513288,
0.9054871850479408, 0.5401627350030654, 0.5405751823521693, 0.5458651963229179, 0.8893413729128014,
0.2563254414476682, 0.55, 0.945, 0.48, 0.7221841742125339, 0.3336272918750665, 0.038562127635426775,
0.01938079190454043, 0.008443431728727331, 0.0356444423457153, 0.05116052220042852, 0.5225413996713586,
0.07627554656680281, 0.018591614680235093, 0.02471752764095385, 0.17471083181228012, 0.9444316523664814,
0.7755663161062373, 0.6483303415676336, 0.31291340730615563, 0.593724805278389, 0.225, 0.07,
0.7380762941909761, 0.6533811066848156, 0.43357837915797653, 0.4367426442671814, 0.7868344424282612,
0.5500069405317359, 0.7788166151753062, 0.5965845697577781, 0.3409759563702998, 0.17008878575561892,
0.3549140244268494, 0.30578785298827127, 0.9141977146139082, 0.43575408925565173, 0.9034096256857836, 0.29,
0.3588996765082006, 0.6434331569225352, 0.49337324747022004, 0.20939187464206163, 0.24117401032705593,
0.8966508273995631, 0.6255151189742083, 0.7915862066248457, 0.93, 0.6, 0.315, 0.395, 0.935, 0.715,
0.11781619832559356, 0.2588047287987097, 0.1695523450543453, 0.3026799958899374, 0.10643205218126113,
0.13755700081267133, 0.15098189145682872, 0.32515113361114234, 0.3093266969466199, 0.3368662734167085,
0.45028874040499284, 0.9106878335503084, 0.03839021807363951, 0.22321667540229953, 0.0056169485521869456,
0.00852379049734887, 0.04806622049219242, 0.06810981951588344, 0.45759790222603686, 0.25639904336625785,
0.6548619413483843, 0.2408295408037225, 0.94, 0.8385579746090666, 0.8997028704577916, 0.9091475116038265,
0.16506914203482967, 0.935, 0.005, 0.3012410713409704, 0.24220459367535585, 0.2495853297046866,
0.7884962921653441, 0.4390664972679941, 0.5035546281440495, 0.4822228997155446, 0.585, 0.21, 0.225, 0.245,
0.4161654776231456, 0.16923936106244938, 0.878937771908554, 0.7440076061583051, 0.4644171950951008, 0.79,
0.595, 0.36648334386132553, 0.4270481113534026, 0.314611993910562, 0.9308560634784939, 0.6366715706398083,
0.9111082577692292, 0.27679343585877214, 0.571696576663083, 0.16913034803613705, 0.19725222948487267,
0.16611606461128386, 0.15675740471044533, 0.15045190182373003, 0.4085440571053141, 0.2825665486213773,
0.2835067609102645, 0.18133359089002832, 0.6325890818192641, 0.17014641389795035, 0.158809508024012,
0.26021379304017533, 0.6881129633724986, 0.27, 0.09, 0.45, 0.36, 0.355, 0.65, 0.015, 0.17534078924566424,
0.23491433798424333, 0.15441047532885666, 0.08378549381262346, 0.12157311307482853, 0.4373911108408695,
0.06585880927319057, 0.09194519895078805, 0.38800177332822644, 0.13534889338962716, 0.1736905153922246,
0.264565637927552, 0.2342677157375326, 0.23824232318857824, 0.24041210486512604, 0.10422722935634292,
0.49052167898175353, 0.18457322975052615, 0.8585983342759266, 0.388004054344146, 0.24383117255976244,
0.5756150846858418, 0.59, 0.9463819971009494, 0.5889070876256425, 0.7345982856429303, 0.852516033074538,
0.3040148767678177, 0.9534336506159015, 0.8078631211664157, 0.660397474770223, 0.3213959920241918,
0.30371067534674634, 0.2454114710760028, 0.9443932441211206, 0.6668468359419256, 0.41356680467831275,
0.4248560155709887, 0.3973572824276227, 0.29723637869036, 0.2767069308104708, 0.17706287421079356,
0.2771811155235187, 0.26738401826804614, 0.2415408738786124, 0.23625263877897432, 0.19369304336980409,
0.19363401055946594, 0.7418142496788108, 0.37940897894679565, 0.2592119029614673, 0.1938316646364672 8,

0.3856822756562754, 0.7201430708081008, 0.5096324748043459, 0.640189648929069, 0.2005446467528731,
0.8839650704669499, 0.7442224502612561, 0.21906526423190656, 0.11710294605134336, 0.169542624952138,
0.11932263275641977, 0.13811633359254014, 0.28590786539325663, 0.40631122587695445, 0.9614166399214636,
0.45639435163017983, 0.27228676618587594, 0.35, 0.15890686518865801, 0.2865688981423716, 0.18734051402975938,
0.26509222668200966, 0.2600245655656794, 0.3435650789620277, 0.27582756324016805, 0.25256020916090643,
0.33175822975963243, 0.49, 0.32, 0.215, 0.585, 0.825, 0.9767726781780091, 0.6952035942207813, 0.53,
0.31258660378851305, 0.02561228304064803, 0.0695899085669562, 0.0424045158722858, 0.011381670829057564,
0.005537148451310972, 0.06686615089115582, 0.7614204545454545, 0.40711307561829, 0.36679630669639307,
0.33924628607588486, 0.5333526640921414, 0.8346303057463562, 0.8415955888696166, 0.4476842415657362,
0.3656334574791139, 0.4490339539915228, 0.21152082294317112, 0.82, 0.23080061747954855, 0.24,
0.27034703034339974, 0.25076908124991126, 0.23029159338437707, 0.17099953314785787, 0.5036623002916027,
0.36442284236841155, 0.4745801062405068, 0.6042772075047657, 0.49824728171132215, 0.5111435946416403,
0.38180206833807306, 0.43897399799274794, 0.295703089164036, 0.21379667093040147, 0.295, 0.23541715951477138,
0.3430798344964988, 0.9670454545454545, 0.16543532516631954, 0.22128144105323774, 0.8378435586377573, 0.165,
0.385, 0.048918799718648584, 0.870772635003073, 0.49487162127214795, 0.3077115123457101, 0.3655711337416182,
0.6281604863489293, 0.41127348735959385, 0.7654450672952414, 0.7276871318474838, 0.42092573883574697,
0.9262375082306498, 0.9484105234825562, 0.115, 0.941521276242044, 0.7653930294624984, 0.813627916799228,
0.6557200628660109, 0.7343991203893587, 0.09676748792178376, 0.11867455613579521, 0.43563642764606286, 0.275,
0.41066910125583705, 0.6286210030807107, 0.34071813092738135, 0.2997729874088446, 0.8679867427508668, 0.965,
0.89, 0.12, 0.17, 0.7847120174847982, 0.19113924911237895, 0.6082635499186586, 0.2631812994351243,
0.5085305962098768, 0.42534168818296086, 0.8226966085844798, 0.1738636341558949, 0.32, 0.035,
0.22249187569058598, 0.21276309501220822, 0.5236745032976661, 0.365, 0.1932972214609801, 0.48232063228360234,
0.9531089808346163, 0.555, 0.765, 0.935, 0.85, 0.935, 0.24183589699204286, 0.2528983085560483,
0.26855628711675733, 0.2913555319361247, 0.5066363124129487, 0.4362887024379134, 0.3543770405370903,
0.24810717712375058, 0.28966100254451127, 0.2702498906970293, 0.29, 0.8473211942417039, 0.15819006830543744,
0.2648461153390201, 0.08, 0.255, 0.07, 0.07, 0.135, 0.445, 0.45, 0.755, 0.08242752993984238,
0.1546755521052769, 0.28074046394674446, 0.2525376039194889, 0.5952420862120291, 0.6694879886117936,
0.6792717357320746, 0.4807030532406785, 0.3500710061551691, 0.4543272514945274, 0.34990893482092494,
0.6947441768370807, 0.545, 0.16748926339985926, 0.7823159557577646, 0.10100903417837427, 0.058159500498584614,
0.07790229796192685, 0.02747334862050592, 0.9386207215541167, 0.095, 0.29, 0.2963230990369585,
0.15921223842275034, 0.1620026374450747, 0.3544345489510414, 0.46054929650998816, 0.45035221024659366,
0.42204298982908783, 0.42754528788708457, 0.43190534210867704, 0.4660051711251135, 0.3985884285199823,
0.43172293699503866, 0.4151779021197231, 0.3713094183559783, 0.22836790737000034, 0.21498307481500814,
0.40001707947223025, 0.4369906441150737, 0.48226351936140666, 0.17916179319777326, 0.19020640022819485,
0.1555072500128188, 0.14118418308555547, 0.1392343839148815, 0.15482687440012122, 0.36795052227775416,
0.38953796553105674, 0.72, 0.2212580322039672, 0.405, 0.65, 0.15711687038009767, 0.3444617486221335,
0.20785349241434037, 0.625, 0.7117469801784826, 0.5210998926875401, 0.47164622054173044, 0.5107588017146051,
0.4401222935966361, 0.19102777062218365, 0.7610234629556131, 0.4054715509515496, 0.13285053945197228,
0.1976837904692382, 0.8509270520831084, 0.175, 0.63, 0.62, 0.2997417640739407, 0.17764942290119975,
0.21724947922480453, 0.19298323483531488, 0.4644433649445608, 0.6025259453606531, 0.5904999225375288,
0.5695157955534017, 0.10847102827248119, 0.43160057859777945, 0.10055454089613057, 0.4421716018657535,
0.3022750437894473, 0.9323746849860629, 0.6033409162651373, 0.20473786969887037, 0.23435862963972828,
0.8989849584497898, 0.5719586005725162, 0.2552254868647194, 0.6912323968037493, 0.5643257955460883,
0.5276731706315103, 0.245, 0.835, 0.5607710062407275, 0.7742572540696275, 0.3322550872194444,
0.28449673258041175, 0.66, 0.47, 0.31200546405964724, 0.3008076554274541, 0.15333205601942768,
0.22008686970610994, 0.1781016830383833, 0.16202127732069932, 0.5517635726486888, 0.16198561006122325,
0.1698930786136132, 0.1384156887137303, 0.2545736861418702, 0.34833048824123625, 0.2033005161986032,
0.7117375041537183, 0.39, 0.10098927877919149, 0.3977618795237862, 0.548317153716984, 0.25452544831411883,
0.4831653285586392, 0.895176343199939, 0.7846509867586374, 0.7, 0.23019931395911108, 0.45415932515465485,
0.9705954512174173, 0.3603198591212616, 0.345, 0.3352695768644392, 0.5611535100467296, 0.475, 0.2, 0.405,
0.3638470878638646, 0.5946156501050764, 0.2784419564440045, 0.409458569165489, 0.6545690293683686, 0.445, 0.8,
0.5570171263804111, 0.23438546000755037, 0.8791748645879081, 0.33591492382585614, 0.3106263325887419,
0.6062447323009162]}, "PLS-DA": {"folds": [{"f1": 0.7117150750391996, "acc": 0.8111888111888111, "bacc":
0.6858974358974359, "auc": 0.8357988165680473}, {"f1": 0.7370596693026599, "acc": 0.8345864661654135, "bacc":
0.7040816326530612, "auc": 0.9341107871720116}, {"f1": 0.6666666666666667, "acc": 0.75, "bacc":
0.654892066162041, "auc": 0.7821698906644238}, {"f1": 0.6436693741041567, "acc": 0.7079646017699115, "bacc":
0.6411804158283032, "auc": 0.6673373574782026}, {"f1": 0.7502955082742316, "acc": 0.8, "bacc":
0.7321428571428572, "auc": 0.8312629399585921}], "chosen": ["{'n': 4}", "{'n': 4}", "{'n': 6}", "{'n': 4}",
"{'n': 4}"], "pooled": {"f1": 0.7043579299180114, "acc": 0.7836166924265843, "bacc": 0.6859217591976212,
"auc": 0.8071384192073846, "sens": 0.4236453201970443, "spec": 0.9481981981981982}, "oof":
[0.47358951073204125, 0.38338668513722385, 0.27932019472436065, 0.3369875110488994, 0.5086713630546167,
0.3785605738104177, 0.11796570550698532, 0.028145872651347215, 0.2031315544006934, 0.12637757317948853,
-0.06042858180766569, -0.01821047689143407, -0.023410426300631892, 0.03654855656992845, 0.5521969194190368,
0.6942739316795082, 1.0559550674413818, 0.4814165867080282, 0.7560929460494634, 0.6590149295605113,
0.21317683213793803, 0.15813925455680497, 0.5671137680929415, 0.6922387295858011, 0.24410272651990622,
0.3693226654008839, 0.5126687781488259, 0.10640639450923117, 0.08530923141051289, 0.49168028210702996,
0.7236736612775557, 0.42022873505768826, 0.3398140002247697, 0.4485587075460549, 0.4816561889601191,
0.888001100338309, 0.6236428837796824, 0.3656609697711292, 0.17532115946018706, 0.324558619244988,
0.18107413488727056, 0.010354963418279128, 0.2792823492414592, 0.48560077853019235, 0.27969111103661737,
0.5970471105789734, 0.4055336127494141, 0.6099224922777122, 0.2976190552020814, 0.25691949645865575,
0.24383445467254755, -0.01320699102796108, 0.3300787900840306, 0.38276099273482855, 0.08688465081302635,
0.29979679842287893, 0.21638053232983298, 0.10860794440126065, 0.22113415480172993, 0.1061461559161839,
0.15307867660881172, 0.07316701418407384, 0.04269388749433323, 0.10430438996376631, -0.017020192456475347,
0.04755764776984961, 0.3915298133795059, 0.2915718663439252, 0.6465963237922042, 0.03569313659806034,
0.504307381888725, 0.20332997775776085, 0.32949235098811247, 1.0266329323509722, 0.9767601984528074,
0.601779008918856, 0.04466325184662373, 0.2298542306211323, 0.2678280675808151, 0.2547441917575636,
0.3356513449469546, 0.25608986746521656, 0.36850365469317026, 0.21676532643089053, 0.2647073983423227,
0.16370811713027916, 0.34071374408316524, 0.2433629273832389, 0.2773873677762797, 0.2342842891854609,
0.3126981266995438, 0.2382686064752884, 0.2665616027984726, 0.12448775339720866, 0.24063678250847825,
0.410548130313504, 0.28381904075127456, 0.14929344132765446, 0.10686734122228683, 0.2214240821997176,
0.12588615974070605, 0.10259671019694017, 0.40825661612525355, 0.08840800915821076, 0.14550418457431763,
1.1998467266401065, 0.6333574474145645, 0.6755240352236663, 0.2185815877069227, 0.40071683175107775,
0.08270776271201533, 0.010324123765060422, 0.09136102173533026, 0.11677066848920484, 0.2365131993394663,

0.11474937748397868, 0.22427825110898, 0.4199901675280472, 0.415567990184477, 0.2130107372837171,
0.4335538435963671, 0.39865950313866017, 0.16957612849888634, 0.36918319404317435, 0.08175193881847975,
0.09422294114113267, 0.3706616319789513, 0.12927246031509157, 0.16948439101271662, 0.17581297368261314,
0.06040458932565884, 0.14427667468598152, 0.10057373528041302, -0.008498915875881086, 0.005757796668754089,
0.148015852214198, 0.340228970964549, 0.14310550857472387, 0.12351576424692462, 0.1351881479916204,
0.21787749047612875, -0.06646538824539894, 0.06099751919054078, 0.23234237821991294, 0.10460472556725026,
0.8997295450794507, 0.8328138527285613, 0.1789868353835791, 0.40434147439136586, -0.2996491027468872,
0.44194790172833665, 0.40582724260238284, 0.186797678997062, 0.13249120509626855, 0.11372991318335654,
0.09891395081079124, 0.0629181576250738, 0.04554355046638853, 0.05371466079196413, 0.6821035198983286,
0.4230184924555113, 0.38320537311830377, 0.04288684948474153, 0.9531562177820481, 0.21053445109106905,
0.29930862183046075, 0.48856906751042095, 0.28625291938475944, 0.6068998249204538, -0.10473093582367515,
0.13658052288175832, 0.15933874323591254, 0.11668801672770082, 0.24117999468731086, 0.1575237988699068,
0.42171445466229884, 0.07120090987259087, 0.10022335709395702, 0.16344215935161371, 0.2217044930676743,
1.0209439392569795, 1.0029224745297527, 0.43943030969996966, -0.05327487446194079, 0.4629435015832949,
0.2333254508479451, 0.293392490772621, 0.6967496939774136, 0.5312144823367658, 0.16361336217998232,
0.36735298543455147, 0.5711032206589972, 0.4575064637158769, 0.5388026707314015, 0.3158653601064723,
0.2309581163925906, 0.07790838334135627, 0.3880668418381419, 0.3398902651742091, 0.4807949972448728,
0.4670826107401224, 0.6805402992308016, -0.05612944485551297, 0.2494315854232516, 0.3429769620995187,
0.4866482729679345, 0.09049011828910938, 0.025521849522779805, 0.6170596854122816, 0.44651662663971803,
0.5301379390602513, 0.25077929445735675, 0.30691351770202696, 0.20343993103901586, 0.3956376261787801,
0.6923268996908594, 0.529671578322899, 0.30906242699825975, 0.36733566504162324, 0.25476542938456354,
0.25412197174520973, 0.2105170268675314, 0.16917488666470137, 0.21388663897591215, 0.4266063524078644,
0.29174000371972086, 0.32992459930689905, 0.37323791673439793, 0.7061990891378975, 0.15050670097964225,
0.22118674551200967, 0.07613931092505227, 0.1713347014188237, 0.18571280566506157, 0.26393414708828655,
0.4266444236020655, 0.3793213489983138, 0.5935890398276668, 0.10357765914551723, 0.47590394346128184,
0.5252666760752964, 0.612191820571102, 0.47237984835269164, 0.24861055052030573, 0.47319369949459167,
0.03566742772428433, 0.26344480636708045, 0.22045567149064865, 0.1869203171314271, 0.4571052091197584,
0.10878143043016167, 0.431673662130884, 0.43897267758735503, 0.4071911636527674, 0.3603523768214705,
0.2822617815919804, 0.2529505964859524, 0.1090857283594936, 0.21571088907834945, 0.4499913441565873,
0.4177473176567753, 0.44074800146519943, 0.906619048035094, 1.8557404986959383, 0.3520146974206687,
0.40322706401506125, 0.15333827286995783, 0.711763806008672, 0.45368538849225304, 0.645885910193838,
0.33172930074325596, 0.42666677529054087, 0.1759082994737246, 0.16942540004795562, 0.12372751026487858,
0.07749747085144726, 0.09712444009495116, 0.23327764875164977, 0.18259454002949363, 0.17456641802466247,
0.04916083826093126, 0.3592361826588396, -0.01994923109554364, -0.03472587582969888, 0.15112154992435636,
0.36481505639300205, 0.1373613509205773, 0.1376621066067472, 0.21355975107604744, 0.17966474747066657,
0.25870671760199637, 0.3624284861123389, 0.09088283555951893, 0.2968469863346885, 0.049874685551802356,
0.14082746478450536, 0.06466854272514211, 0.17796185941552345, 0.3693380920444219, 0.15358299501391792,
0.1871739375522016, 0.3300624559159452, 0.050432934381192895, 0.06849097285859065, 0.18423512685837173,
0.20310352081038016, 0.15608281910648555, 0.18272428198686927, 0.15661266132272836, 0.3757116574989495,
0.14974594695128043, 0.6017775575123208, 0.2838760760784846, 0.0869206710447478, 0.29616139275640424,
0.29556326956407275, 0.8035659274856485, 0.2829746728198432, 0.5899258471843629, 0.7302329929236295,
0.09181986468010758, 1.5784999378740774, 0.4157445155392027, 0.2754752021541799, -0.2919067638676643,
0.07949174977432444, 0.2509905320716539, 0.7516748121623663, 0.44094389683399676, 0.3766378042845614,
0.3902879212589569, 0.38942212421742145, -0.06744118183173387, 0.33550941991795546, 0.1371642893747104,
0.2379033315328382, 0.15223363371527748, 0.27626232358117664, 0.29710521021001013, 0.1914483651018397,
-0.005065626774457654, 0.326681124322963, 0.4065682907493199, 0.24661815953040875, 0.07814553284820083,
0.28770189102052146, 0.47564550674954265, 0.1503174522600151, 0.46905122264467114, 0.28251230937885574,
0.6258267138430235, 0.3487642096885652, -0.0094801866817672, 0.21799128305512894, 0.24786942041606308,
0.08780466752707347, 0.24345580024190994, 0.31051468244951874, 0.3276729095786175, 0.6029224806141235,
0.3636872654448104, 0.3627813932229454, 0.14635635639172662, 0.28984918114714725, 0.3423076318670152,
0.23104950165295876, 0.2712502002292556, 0.08215135767203288, 0.24463350682628832, 0.2847540474868448,
0.1118066351414313, 0.3432947736493211, 0.6546804852521805, 0.5144303796535827, 0.10693983349884212,
0.3897232903818383, 0.6352920389111548, 0.6375968246820871, 0.5220537978382414, 0.4795845655363821,
0.20700535748811377, -0.05362989672963492, -0.093349041370127, 0.007466533951568266, -0.10165345346278531,
-0.06089042473558187, -0.026290996928941435, 0.4945424052757159, 0.37804596360963016, 0.16563563218674304,
0.28129892462852707, 0.44649712560157817, 0.7753058585106656, 0.6815501522142915, 0.1342888586774821,
0.012457927786433398, -0.015507613774657647, 0.24549054257443473, 1.014836717747178, 0.07582473350677849,
0.33831355455059414, 0.2688837154561091, 0.023245128622548683, 0.045680932328492074, 0.0909572030418758,
0.4002772392010584, 0.3208356600568765, 0.0822503773265208, 0.4539744513980426, 0.6345926575454633,
0.5852449917843279, 0.17762665354359028, 0.08387324187313575, 0.13871049987152237, 0.2592495778954334,
0.20119942551248082, 0.23814871311255786, 0.3808820631810169, 0.8762543947129711, 0.09886749152177995,
-0.041884299301209904, 0.5341460929729978, 0.1841835370893039, 0.174146234403432, 0.08149210606468607,
0.4701485195164111, 0.3389076099076012, 0.14845363126719846, 0.22842696716035882, 0.2960898536123008,
0.3053260005201783, 0.8637434820741143, 1.862085134170129, 0.35619508785138687, 0.5566233967905307,
0.661302056211828, 0.2774598291132946, 0.6915827509146082, 0.4379510220674318, 0.5958849436307039,
0.5417969854525955, 0.8150858301299417, 0.09592007331661084, 0.08500573767846623, 0.1674211805304209,
0.23164681571833745, 0.15053699616676766, 0.6675207519836555, 0.2564791145901442, 0.25003837410246316,
0.49610436305944305, 0.5239416957581755, 0.5716153399932327, 0.19991009338755336, 0.18430657340321457,
0.6169735597981227, 0.1405099500790138, 0.48384792337511917, 0.01669490869340673, 0.27617392311259076,
0.26661519801015593, 0.5240451534138634, 0.21813252693137714, 0.3303214762140618, 0.2551860587599719,
0.07341173769856657, 0.10691324048526571, 0.4066747817247459, 1.2059943371127124, 0.1989160050120948,
0.483766991282527, 0.7837550058220499, 0.42983639373497684, 1.3833938990789632, 1.1650237983950644,
1.1834014133436885, 1.119810643810336, 0.1195495480837056, 0.05229249080094439, 0.16674112286713103,
0.06931932588520479, 0.206923969402453, 0.03527738995116658, 0.06824837543484435, 0.26660921620895134,
0.1444879067884851, 0.24863931331258152, 0.31415076855354135, 0.5615562744354554, 0.051749267318624004,
0.11074459949170501, 0.043976159988245334, 0.15776557409928002, 0.10268731531577482, 0.17075005909307023,
0.21185540199451897, 0.14721631065691135, 0.2226796997559325, 0.498685313137217, 0.15902823704468044,
0.21345844625167607, 0.2857357431692063, 0.28397493400220764, 0.15140201241171228, 0.05886038791632936,
0.051802186919128856, 0.4007844447120974, 0.33350832275460285, 0.3449776523619509, 0.358507622035869,
0.5161366734396204, 0.33777382897698904, 0.23584370966213827, 0.4684761590955481, 0.2540992102490038,
0.09786018990900694, 0.19835631495056055, 0.1320042532658114, 0.6526160005580757, 0.1337009325145631,
0.003992691944866844, 0.17356663963837235, -0.008289398843031426, 0.06665062930096649, 0.2855456138552375,
0.18133788246127702, 0.22456570241240587, 0.12638108682701776, 0.19775335387160248, 0.20473357463925265,

0.23801260931596788, 0.058420545527110845, 0.191880645980132, -0.006636826925207173, 0.057458040645130604,
0.11665113360667523, 0.1078002355300832, 0.08903220962619385, 0.12459542147746303, 0.20396629708525918,
0.22846603680997307, 0.178904348840621, 0.13958449311017165, 0.12423385795297631, 0.08395758518570415,
0.1352335682343032, 0.11239518902673026, 0.3623913067570818, 0.9838113430173555, 0.29427488628443554,
0.15449907983239897, 0.4147715215320882, -0.046233918011176156, 0.15654857917201875, 0.18419742600697497,
0.6024869485633472, 0.5406553142487815, 0.46181806378570645, 0.38742218812922946, 0.3792355735659955,
0.28445888068267267, 0.015914325185261258, 0.6417969031333296, 0.2826768881205328, 0.1214820126648479,
0.1831870828571717, 0.65806411849833, 0.33420240301352167, 0.45549572643048486, 0.5876382492877756,
0.4004989576502541, 0.2955513663973335, 0.2815726246871768, 0.28048700299034196, 0.3668392264442339,
0.5591223418396839, 0.5196969905403132, 0.46958017684747466, 0.13854470570388838, 0.3361036649189233,
0.19688599439590526, 0.1609041861634452, 0.18220635463274393, 0.9920423229649404, 0.40576266841601427,
0.04283801314088559, 0.2714272805206732, 0.771996704955338, 2.206945468134762, 0.20924859105161764,
0.873666006183724, 0.34506633285733973, 0.45558999809559264, 0.29246777876519847, 0.6885712223091867,
0.4236535086447984, 0.4553769731665061, 0.07383499610546246, 0.28167744397687877, 0.593524523083389,
0.45335605475144447, 0.04746301301836 3975, 0.033755304729482405, 0.11324520972677846, -0.0024306820816142327,
0.06259041177864422, 0.17786620927884375, 0.30838575705306576, 0.10570380673596769, 0.13379531536039935,
0.0677833147555788, 0.32130481113764214, 0.15057472493031634, 0.13460206140639958, 0.40766050560972633,
0.6745594579300312, 0.13342373420393494, 0.40427544677442784, 0.4538478806891816, 0.33317188740932974,
0.45812051704230294, 0.8379590895738889, 0.6344491943276528, 0.34263232864666254, 0.2702802194408434,
0.49714770175102074, 1.0652436476807188, -0.5538209540299706, 0.23229806738462241, 0.03604163977482938,
0.6119672512204863, -0.011328937637609171, 0.30478498976780294, 0.3555183445003976, 0.317055637993563,
0.38210737989074295, 0.3650723519282849, 0.40247129452981145, 0.616034440421808, 0.2783548312279585,
0.4065782344014212, 0.7202966018029235, 0.22883411676655152, 0.425133153221398, 0.2948117557110892,
0.1260578988324757, 0.3748848101869548]}}, "y_A": [1, 1, 1, 1, 1, 1, 0, 0, 0, 1, 1, 0, 0, 0, 1, 1, 1, 1, 1, 1,
1, 0, 1, 1, 1, 1, 0, 0, 0, 1, 1, 1, 1, 1, 1, 1, 0, 0, 0, 0, 0, 0, 0, 1, 1, 1, 1, 0, 1, 0, 0, 0, 0, 0, 0, 0,
0, 0, 0, 0, 0, 0, 0, 0, 0, 1, 1, 1, 0, 1, 0, 0, 1, 1, 1, 0, 0, 0, 0, 0, 0, 0, 0, 0, 0, 0, 0, 0, 0, 0, 0, 0,
0, 1, 1, 1, 0, 0, 0, 0, 1, 0, 0, 1, 1, 1, 0, 0, 0, 0, 0, 0, 0, 1, 0, 0, 0, 1, 0, 0, 0, 0, 0, 0, 0, 0, 0, 0,
0, 0, 0, 0, 0, 0, 0, 0, 0, 0, 0, 0, 0, 0, 0, 1, 1, 1, 0, 1, 0, 0, 0, 0, 0, 0, 0, 1, 1, 0, 0, 1, 1, 1, 1, 1,
1, 0, 0, 0, 0, 0, 0, 0, 0, 0, 0, 0, 1, 1, 1, 1, 1, 0, 0, 1, 0, 1, 0, 0, 0, 1, 0, 0, 0, 0, 0, 1, 0, 1, 1, 1, 0,
1, 0, 0, 1, 0, 1, 1, 1, 1, 1, 1, 1, 0, 0, 0, 0, 0, 0, 0, 1, 0, 1, 1, 0, 0, 0, 0, 0, 0, 1, 1, 1, 1, 1, 1, 1,
1, 0, 1, 0, 0, 0, 0, 1, 0, 0, 0, 0, 0, 0, 0, 0, 1, 1, 1, 0, 0, 0, 0, 0, 0, 0, 0, 0, 0, 0, 0, 0, 0, 0, 0, 0,
0, 0, 0, 0, 0, 0, 1, 0, 1, 0, 0, 0, 0, 0, 0, 0, 0, 0, 0, 0, 0, 0, 0, 0, 0, 0, 0, 0, 0, 1, 1, 1, 0, 0, 1, 0,
1, 0, 1, 1, 0, 1, 1, 0, 1, 0, 0, 1, 1, 0, 0, 0, 0, 0, 0, 0, 0, 0, 0, 0, 0, 0, 0, 0, 0, 0, 0, 0, 0, 1, 1, 0,
0, 0, 0, 0, 0, 0, 1, 0, 0, 0, 0, 0, 0, 0, 0, 0, 1, 0, 1, 1, 1, 1, 1, 1, 1, 1, 0, 0, 0, 0, 0, 0, 0, 0, 0, 0,
0, 1, 1, 1, 0, 0, 1, 1, 1, 0, 1, 1, 0, 0, 0, 0, 0, 0, 1, 0, 0, 0, 0, 0, 1, 0, 1, 0, 0, 0, 0, 0, 1, 0, 0, 0, 1, 1,
1, 0, 0, 0, 1, 1, 1, 1, 1, 0, 1, 1, 1, 1, 1, 0, 0, 0, 1, 1, 1, 1, 0, 1, 1, 1, 0, 0, 0, 0, 0, 0, 0, 0, 1, 0, 0,
0, 0, 0, 0, 0, 1, 0, 0, 1, 1, 1, 1, 1, 0, 0, 0, 0, 0, 0, 1, 1, 1, 1, 0, 1, 0, 0, 0, 0, 0, 0, 0, 0, 0, 0, 0, 0,
0, 1, 0, 0, 0, 0, 0, 0, 1, 0, 0, 1, 0, 0, 0, 0, 0, 1, 0, 0, 0, 0, 0, 0, 0, 0, 0, 0, 0, 0, 0, 0, 0, 0, 0, 0,
0, 0, 0, 0, 0, 0, 0, 0, 0, 0, 1, 1, 0, 0, 1, 0, 0, 1, 0, 1, 1, 1, 0, 0, 1, 1, 0, 0, 1, 0, 0, 1, 1, 0, 0, 0, 0,
0, 0, 0, 0, 0, 0, 0, 0, 1, 1, 0, 0, 0, 1, 1, 1, 0, 0, 0, 1, 1, 1, 0, 0, 1, 1, 0, 0, 0, 0, 0, 0, 0, 0, 0, 0,
1, 0, 1, 1, 0, 0, 0, 0, 0, 1, 1, 0, 1, 1, 1, 1, 0, 1, 0, 1, 0, 0, 0, 0, 0, 0, 0, 0, 0, 0, 0, 0, 0, 0],
"repeat": {"SVM": [{"f1": 0.7602920344339108, "acc": 0.7897990726429676, "bacc": 0.7652942351218213, "auc":
0.8434518262104469, "sens": 0.6995073891625616, "spec": 0.831081081081081}, {"f1": 0.7646226605764178, "acc":
0.7913446676970634, "bacc": 0.7731050015532774, "auc": 0.8334331868814626, "sens": 0.7241379310344828, "spec":
0.8220720720720721}, {"f1": 0.7582140754342619, "acc": 0.7851622874806801, "bacc": 0.767263568987707, "auc":
0.8519671148981494, "sens": 0.7192118226600985, "spec": 0.8153153153153153}, {"f1": 0.7649696378225981, "acc":
0.7928902627511591, "bacc": 0.7715572715572716, "auc": 0.8383093684817822, "sens": 0.7142857142857143, "spec":
0.8288288288288288}, {"f1": 0.7730731044009411, "acc": 0.7990726429675425, "bacc": 0.7814094883060401, "auc":
0.8436459858873652, "sens": 0.7339901477832512, "spec": 0.8288288288288288}], "LogReg": [{"f1":
0.753504109552545, "acc": 0.7789799072642968, "bacc": 0.7654329206053344, "auc": 0.8342098255891358, "sens":
0.729064039408867, "spec": 0.8018018018018018}, {"f1": 0.7388524207082616, "acc": 0.7650695517774343, "bacc":
0.7512870012870013, "auc": 0.8293391914081569, "sens": 0.7142857142857143, "spec": 0.7882882882882883}, {"f1":
0.7761358023713119, "acc": 0.7975270479134466, "bacc": 0.7923157147295079, "auc": 0.8554120623086141, "sens":
0.7783251231527094, "spec": 0.8063063063063063}, {"f1": 0.7618582767891435, "acc": 0.7867078825347759, "bacc":
0.7737374073580969, "auc": 0.8497093152265566, "sens": 0.7389162561576355, "spec": 0.8085585585585585}, {"f1":
0.7462745098039216, "acc": 0.7712519319938176, "bacc": 0.7598022899747038, "auc": 0.8302489681800027, "sens":
0.729064039408867, "spec": 0.7905405405405406}]}, "six": {"pooled": {"f1": 0.39453775302164895, "acc":
0.5564142194744977, "bacc": 0.43141031303727734, "auc": 0.8009147686941093}, "folds": [{"f1":
0.32188525395421946, "acc": 0.4899328859060403, "bacc": 0.40345011171624073, "auc": 0.8265377576701205},
{"f1": 0.373648889912748, "acc": 0.5815602836879432, "bacc": 0.37737079184447603, "auc": 0.7277815196148177},
{"f1": 0.37966195797315344, "acc": 0.592, "bacc": 0.408179012345679, "auc": 0.8376853838646059}, {"f1":
0.35119743605157955, "acc": 0.5853658536585366, "bacc": 0.3650252525252525, "auc": 0.8154555858553244}, {"f1":
0.4609459984459985, "acc": 0.5412844036697247, "bacc": 0.5952204228520017, "auc": 0.825169540518970 2}],
"chosen": ["{'C': 50, 'gamma': 0.001}", "{'C': 10, 'gamma': 'scale'}", "{'C': 50, 'gamma': 'scale'}", "{'C':
10, 'gamma': 0.01}", "{'C': 100, 'gamma': 0.001}"], "cm": [[206, 19, 13, 7, 20, 31], [15, 84, 33, 2, 12, 13],
[4, 19, 10, 0, 2, 3], [0, 1, 0, 2, 1, 2], [2, 8, 2, 5, 15, 9], [34, 11, 5, 2, 12, 43]], "recall":
[0.6959459459459459, 0.5283018867924528, 0.2631578947368421, 0.3333333333333333, 0.36585365853658536,
0.40186915887850466], "classes": ["ACK", "CBC", "CEC", "MEL", "NEV", "SEK"], "n": 647}, "label": {"A": {"n":
647, "patients": 319, "mean": {"f1": 0.764234302533626, "bacc": 0.7717259131052234, "auc":
0.8421614964718414}, "sd": {"f1": 0.00511078409843758, "bacc": 0.005603230323562229, "auc":
0.00618127965293236}}, "B": {"n": 351, "patients": 225, "mean": {"f1": 0.7425110499044874, "bacc":
0.747400479297031, "auc": 0.813400346158967}, "sd": {"f1": 0.013322632510401366, "bacc": 0.01370883071353827,
"auc": 0.00958943947891749}}, "C": {"n": 647, "patients": 319, "mean": {"f1": 0.6767281244852306, "bacc":
0.7096666305584141, "auc": 0.7661620538374045}, "sd": {"f1": 0.010874735318317947, "bacc":
0.011838498351777079, "auc": 0.012187347942112242}}, "D": {"n": 647, "patients": 319, "mean": {"f1":
0.656077419987241, "bacc": 0.6610238317134869, "auc": 0.8227227637133655}, "sd": {"f1": 0.008928594237647799,
"bacc": 0.009643172336541456, "auc": 0.003959866580338907}}, "BEN_vs_ACK": {"n": 444, "patients": 186, "mean":
{"f1": 0.7290643939171952, "bacc": 0.7385135135135135, "auc": 0.7939508765522278}, "sd": {"f1":
0.019679596988634057, "bacc": 0.0206440758343949, "auc": 0.019307427947047748}}, "ACK_vs_MAL": {"n": 499,
"patients": 250, "mean": {"f1": 0.8021852149643731, "bacc": 0.804871188922913, "auc": 0.8761632938357078},
"sd": {"f1": 0.009983308634823725, "bacc": 0.010687343236279104, "auc": 0.006856390962880098}}}, "stability":
{"jaccard5": 0.24087301587301585, "jaccard10": 0.3350215750095059, "spearman": 0.512392782941498, "top1":
{"1032.0": 24, "1050.6": 12, "988.6": 6, "908.1": 4, "926.7": 4, "994.8": 2, "1038.2": 2, "1044.4": 2,

"914.3": 1, "939.1": 1, "951.5": 1, "1570.9": 1}, "top1_908_1051": 59, "imps": [[0.26258329397588387, 0.21367603645900363, 0.14676330367992255, 0.3321782041276737, 0.25896559789527823, 0.10759927653075724, 0.0533224488043347, 0.10065610731146149, 0.11894623374079112, 0.044675283249395036, 0.10187137306428931, 0.16192885228413514, 0.16870134175590645, 0.28258166093215026, 0.16950601108202523, 0.008166606308355091, 0.036850736990361996, 0.029524761471843148, 0.1991786948400825, 0.29814761242206367, 0.3985182952813, 0.338962474616511, 0.4305505258377559, 0.40235675317416136, 0.08183269998215928, 0.04857647017043358, 0.08888510632301107, 0.01977529372160439, 0.09755698215841156, 0.04769308849293026, 0.0481371839638692, 0.14238939041843282, 0.2014739489484527, 0.09257338148525654, 0.14581337984759665, 0.14250256966020788, 0.16836115021760814, 0.018775603186493798, 0.12530801045815454, 0.06352801275102345, 0.16292404769357455, 0.17817683247580957, 0.09499493603403664, 0.1412818404003677, 0.052970709756954205, 0.013646818979719403, 0.09834404772888043, 0.05173569781410722, 0.02653952897312838, 0.07738417432017906, 0.04827435387062933, 0.031604137977166095, 0.04147076295845116, 0.11235655916775796, 0.09879081639082642, 0.06166310160671117, 0.012566022765312794, 0.0059412330399967435, 0.058870859166699846, 0.05171678098627587, 0.058916279897114565, 0.14364000797540838, 0.16839636084774406, 0.14549589048649336, 0.12439988744967172, 0.10441179938207111, 0.026635186976808934, 0.005850102454550963, 0.004821886313908808, 0.049883045213964825, 0.057365134135961116, 0.07613729446626517, 0.15463469461022064, 0.2417778576660058, 0.1747272663697492, 0.0770587380335137, 0.03665848493195283, 0.10020005377149767, 0.11701899787237283, 0.04589362332676069, 0.047204535962756144, 0.07289644488387144, 0.1286306187200106, 0.17319924249725996, 0.16029779762998594, 0.15698911166133775, 0.14806074022485852, 0.13059719318845484, 0.11006537000770174, 0.0507984922114037, 0.05186503550414723, 0.09508695498627967, 0.007894994262900362, 0.04806557029771485, 0.058401739882203166, 0.058856980489575814, 0.0693060491168588, 0.05391490379762038, 0.030526517746518007, 0.07382124250242476, 0.10724892936420463, 0.09200228809700804, 0.10258735117119486, 0.03167828384303323, 0.027633966774291842, 0.18317003475777416, 0.1729437736560755, 0.2788957003945686, 0.21998865488952254, 0.2302856346559439, 0.26120571283676003, 0.21457727897315365, 0.048590862277676776, 0.021988258579062166, 0.06782312512478422, 0.02466325359724348, 0.06793424360463583, 0.029365142747716684, 0.03053189469481465, 0.05181847633304738, 0.07647013058851032, 0.03750643706868261, 0.06266934694890788, 0.1118190878445368, 0.018241823679882287], [0.31240157615254277, 0.46002125099945523, 0.10210108143124295, 0.2550204659459295, 0.07968891182627791, 0.0969685970509421, 0.04037489491388828, 0.2784485797417979, 0.13309361819145007, 0.02020211130146994, 0.06349724985447285, 0.1936264845461775, 0.26703902837862, 0.1550873834155559, 0.11369858071233874, 0.2819646929244477, 0.1416125169051843, 0.15625992826925522, 0.1367750702138592, 0.054116350184467976, 0.5143578732755049, 0.23361980785075173, 0.4562627978494122, 0.6213313803895615, 0.2590002528355239, 0.17464641372084722, 0.028199501684541783, 0.09638991015039014, 0.19732210898728006, 0.05573079737502073, 0.23731339759892012, 0.29670177979991713, 0.36525431525498464, 0.17552800480732458, 0.060158883146336245, 0.058462093258614074, 0.05117916154154506, 0.2596448677706354, 0.309055967989949, 0.35163926848884014, 0.2935103473203635, 0.21890279068799842, 0.002302610697457098, 0.015054832701856516, 0.0008072044506045434, 0.024481252695660746, 0.09012213927654347, 0.13303102642041142, 0.01227760477818279, 0.05330338145328017, 0.029483737183707514, 0.050462215423654, 0.012697202227872477, 0.011417526324887808, 0.005684882984147783, 0.002940854842444413, 0.05465837638770094, 0.04880220471166898, 0.022963263617352774, 0.07310170515803811, 0.040425465454717174, 0.06075869180884835, 0.1182481637839135, 0.13724833451206656, 0.1401003213719978, 0.1686049164405579, 0.10290569325383299, 0.050145960074730746, 0.04870415643789595, 0.017077282477958738, 0.0209878230484905, 0.006093075099083023, 0.07504659656221768, 0.2143683565510302, 0.13979988085278702, 0.0246535114047673, 0.054484581788719566, 0.1126479818480781, 0.11664803363354478, 0.06710784129881059, 0.08238589264309318, 0.06610491911530098, 0.08552378628493885, 0.10738007792824576, 0.12671278174456277, 0.11003445377055632, 0.1114767930792212, 0.0690786302669759, 0.024515242333267483, 0.15875654501715705, 0.21725141345288262, 0.2849737136749026, 0.1142930633769414, 0.006364783758141761, 0.07188608214526854, 0.09191735915707161, 0.1300100605257431, 0.09845953715944866, 0.024253472601429356, 0.02250177115165145, 0.03260959701034907, 0.024292561630177453, 0.01685050075973259, 0.04727619807857622, 0.07725016875532244, 0.1485295613087427, 0.16355224972004295, 0.37486098378862476, 0.26260750521710363, 0.20462582989059383, 0.12815145113256393, 0.07857929067222999, 0.05970734047524296, 0.14635760257207828, 0.1131132161404143, 0.10317760697821503, 0.09492937172081917, 0.014394024798403729, 0.047123382889214985, 0.0797313750325573, 0.026619759967801224, 0.04540540931725509, 0.003490516585040549, 0.16210532031421154, 0.14618585461109318], [0.3395280959071207, 0.08692871279440426, 0.06825560844087529, 0.3507911937971343, 0.09333887883559133, 0.20920750578151823, 0.10424801404170964, 0.1457877266197197, 0.12231570909408016, 0.09216275294809423, 0.014834572909299345, 0.0638360197468573, 0.3337803440434552, 0.3544821256881715, 0.356059770958263, 0.07982287281855578, 0.06901890634119544, 0.09022643654153101, 0.048874538237204085, 0.1582155395597591, 0.4389190410347601, 0.28092607676721765, 0.18925987629959448, 0.27570970251390853, 0.0631996834964817, 0.00839343503677235, 0.024497603074848533, 0.05531408736098216, 0.12774670791983658, 0.05331297089979192, 0.009838719966252409, 0.14209937935952108, 0.2500018619753872, 0.004298274501732354, 0.013510805213239326, 0.11458220276906378, 0.04027923187089091, 0.09653874602074781, 0.2684342807260537, 0.14538913824849484, 0.2473832234337453, 0.15894083417141286, 0.08752036541682427, 0.01660427955406794, 0.03288805852908602, 0.09183889260445177, 0.06545540851156731, 0.1687325221208098, 0.07170345706153725, 0.05618250729625482, 0.033119392639051524, 0.061297127270714506, 0.030379588138761085, 0.0023950236759159027, 0.009563943775023162, 0.01391666914428912, 0.0004777750421787827, 0.0227213985342861, 0.0315037987680139, 0.01507848901032175, 0.028689943533789588, 0.04937541588870947, 0.09747637307210819, 0.1070635198113213, 0.06544786436656319, 0.09288699335911531, 0.019221476931423002, 0.02390597530912369, 0.021251872588474205, 0.08188318686264014, 0.0849485485531732, 0.05415431464509428, 0.009790268571303903, 0.013226378908117443, 0.02738931595583046, 0.06851377587141308, 0.044582654327561706, 0.07401516526902692, 0.18497596737693198, 0.160007741542536, 0.1245482107289503, 0.11153014665369458, 0.12860513904192045, 0.1321963036574433, 0.109796828755583, 0.12807359470247798, 0.09991395825647173, 0.0862551582574121, 0.058088925041124895, 0.014511825989203376, 0.034296580199537845, 0.1393281747882382, 0.019555017596752582, 0.020105709459227124, 0.00967025546127791, 0.00688212516951978, 0.03241092395732296, 0.051508206492008116, 0.09543001129238823, 0.009148516852445327, 0.04478134150920281, 0.10462901662496134, 0.06764398545986175, 0.036113079397257976, 0.029304616400459386, 0.13915851249998704, 0.08973955593304773, 0.3023070536392057, 0.22284100297116888, 0.20052301099755707, 0.2820058442339237, 0.1418016311326085, 0.046615267606486585, 0.04809270570058802, 0.055133629587928334, 0.014827828504223067, 0.01148321197622422, 0.050839909193238074, 0.07956860812658273, 0.1582051447578948, 0.13962063658601606, 0.07974220819015485, 0.2324765844551356, 0.2932188987126706, 0.2769750843949778], [0.4908066613086241, 0.15626611487032088, 0.04728532160059661, 0.22803998096881392, 0.3511035204332406, 0.0152566556239797, 0.02598859119250867, 0.14269705881946443, 0.06619393208355338, 0.04003641002790377, 0.2215299525935308, 0.06171536685404267, 0.19264419671285474, 0.5314217229822359, 0.16166529627898824, 0.2384110712548468, 0.020980017046376767, 0.25413565045192316, 0.07097581481292416, 0.002298746905528509, 0.5559068680066339, 0.37670517575255075, 0.29840456390867015, 0.3820982862149257, 0.12800092976046173, 0.046100060433571906, 0.08211551225218834, 0.0022854412519901514, 0.2935376093648827, 0.023066104908208658, 0.009566632672510409, 0.0770998705938731, 0.3201060328653799, 0.2290824552858528, 0.13147166933946938, 0.37900188689310116,

0.1579331306940592, 0.0981967132822017, 0.295761750585511, 0.07783910093394246, 0.3180261745955821,
0.24067068566069227, 0.09192655141327333, 0.07854380029778159, 0.08800590855353173, 0.04891649709286864,
0.05810784976879038, 0.1877576117120626, 0.02709508139100791, 0.01969892580240391, 0.048721809872919464,
0.06434829509182488, 0.013041565231141573, 0.061137474557938484, 0.015479053708915506, 0.020952652103950243,
0.04762014996080196, 0.012167920370006283, 0.049143886964167334, 0.04075373581552017, 0.10161976132879408,
0.17110196647896897, 0.23431246439934195, 0.24140337845295043, 0.19368790618934165, 0.16075126555540561,
0.08756079886638844, 0.003307558001798667 5, 0.06452082588366048, 0.134947844275924, 0.1395279174745807,
0.1792706362611991, 0.18032153593469227, 0.1690254771955076, 0.10456993660999106, 0.06718083840128464,
0.05309405566788274, 0.059708839639395335, 0.11197156628976182, 0.016137819798886052, 0.009918569886401388,
0.020802045269744288, 0.059325957650273, 0.08967143270290114, 0.10516559122950728, 0.11283300303544089,
0.1342187010825434, 0.10328277464484667, 0.07920870517620615, 0.06231556488982213, 0.1095292036223098,
0.1980987926266863, 0.06413993115170022, 0.06893535944153818, 0.030954418246499964, 0.01130705750124949,
0.01234447902606198, 0.06127494901389793, 0.02218410923410708, 0.11312065191942153, 0.11054394744195331,
0.10597141136116917, 0.04521317228998129, 0.11425333151518201, 0.04830084594558529, 0.02874108490712995,
0.05770923006263346, 0.23147433740178533, 0.16153582614864184, 0.24201218732451582, 0.3048991697708168,
0.2004495331377033, 0.13196082725518515, 0.0100314112135633 4, 0.041111987981741656, 0.01956848699652123,
0.003356175004555904, 0.010619307304971681, 0.009500758043424413, 0.0783115127610041, 0.07415435653667057,
0.09119981238404842, 0.24771541277172773, 0.3044435438049409, 0.10227087296340569], [0.35223036613413694,
0.03728168417980568, 0.03512298912562204, 0.5851790374636613, 0.22100607275820752, 0.16488731707534263,
0.09554153454728073, 0.2153087232967419, 0.17856032152359375, 0.0014698344645267768, 0.13807466017862396,
0.13928916875082997, 0.2473662561461192, 0.3696086087464661, 0.3416915521696095, 0.16791384848656535,
0.20232258185526977, 0.0836862220015872, 0.06309763551987775, 0.1459813207599899, 0.5470165091573469,
0.4269763286145888, 0.2256384451186241, 0.26790880944186085, 0.07654761146891562, 0.08969156695872944,
0.0603596927029286, 0.0530307101119353, 0.15425308239367783, 0.09020699799140026, 0.10634208577315457,
0.19551052476207018, 0.3260714161560918, 0.18858441262471068, 0.035886910195319505, 0.2831055137348305,
0.23846569438702786, 0.15729807521557926, 0.33067198151384014, 0.22241280307304864, 0.3111579610613722,
0.21240627510259574, 0.04737131913933826, 0.03021219112849942, 0.09597571538743288, 0.12334940252063761,
0.008063983866060479, 0.20212490140044645, 0.005148762135015515 6, 0.029327675171706605, 0.09005271440868859,
0.12074734940395732, 0.06320450703686449, 0.025012235929007573, 0.1000417994413746, 0.10680758584050233,
0.02665739968936947, 0.03236762030148091, 0.04643949814739623, 0.004435475136585639, 0.0923717548010075,
0.16749746734526103, 0.19431314187212553, 0.18815784980790584, 0.1684708247621616, 0.12239191718416198,
0.05719179614955786, 0.01798943489114438, 0.019295012093038598, 0.08586287602504326, 0.1280100420499474,
0.1770228140511296, 0.22154411956035877, 0.2889401064555992, 0.2563110260431736, 0.16848721784512496,
0.08975765178164809, 0.05628286743963652, 0.03412032298594554, 0.02893779886312424, 0.006391182123221503,
0.024654491695946003, 0.12512214945323172, 0.16638138545072925, 0.14110345085271775, 0.1464627333961448,
0.16490171608552465, 0.1250061979143643, 0.10626568272937521, 0.005519192595889805, 0.08991949146873325,
0.17520623712819755, 0.006872834313374296, 0.028563146507471295, 0.015107393763696554, 0.007996881962270498,
0.01649819523933714, 0.015684364824744747, 0.08479997529976228, 0.02526340461267954, 0.07569566821057189,
0.08804845929316818, 0.07426209051041313, 0.06546351024601578, 0.02837051076442367, 0.03945304101851094,
0.09125155067555088, 0.33339297119555095, 0.28756410575650726, 0.29900384189441304, 0.37445645251992327,
0.23640932348257532, 0.11176188837184974, 0.04870955099563197, 0.019172453099563197, 0.021356443616915223,
0.0023301558202222725, 0.012258600570331598, 0.020841389890378155, 0.055919866289417926, 0.07612207943063211,
0.15272212555943376, 0.24876439504393058, 0.32153935669875666, 0.15219792155687767], [0.3741231029201813,
0.2349189089645383, 0.11017786933223402, 0.3544790456206823, 0.14420704683237284, 0.2354147874329467,
0.08799970961838166, 0.15604979985819226, 0.019088076170079696, 0.049079669122549865, 0.04032980318161481,
0.04474111120408203, 0.16710958102060658, 0.28109336077622676, 0.19335995772006107, 0.010985519631782754,
0.07029690209545235, 0.011700069773074421, 0.0983983807555742, 0.018541543732279748, 0.5458852867558657,
0.22158559818698778, 0.05087523591675577, 0.24026065660122956, 0.07871858525136669, 0.04607258062460986,
0.028868999375037698, 0.04170759272759917, 0.19383313435418276, 0.061923951405705245, 0.0027417172382460772,
0.1973805011689617, 0.2940704292945655, 0.26004538981262054, 0.18734411002822818, 0.2193702523715168,
0.17501003354753455, 0.016427750551999163, 0.13358635196439392, 0.18493288585629158, 0.29097598645446066,
0.19777997683848922, 0.07993435646740868, 0.04131383161278293, 0.047319023737114885, 0.1360690436044275,
0.004049074451196464 5, 0.09539662788267987, 0.005273296970159257, 0.0034513271624006976, 0.07310112065883992,
0.07397836910752421, 0.051170168850425274, 0.0096956715186819, 0.03310867834336046, 0.043898653442291116,
0.021116478926257856, 0.007901074412919306, 0.03165935455513412, 0.06121427073017956, 0.005341728429183127,
0.0646437307638985, 0.1409460765851932, 0.1800547987523711, 0.13433699305163033, 0.11334379847580213,
0.06460622298140355, 0.02683497985654347, 0.010209963963912024, 0.012568585524415766, 0.0028058473486203455,
0.004443219003539654, 0.038944392995519266, 0.08961858012661501, 0.07193993383824498, 0.040848044131388456,
0.04495124504379184 6, 0.11389094731598567, 0.15983565876455896, 0.058903286855249135, 0.028993850312524595,
0.03329599957783351, 0.060270051178133825, 0.08898282637254593, 0.10588507791123954, 0.10785207063998513,
0.08695810589665624, 0.03974361048243437, 0.05346193655033265, 0.21982654407863295, 0.2699457733484589,
0.3641923718766742, 0.22923926389827864, 0.2045842937085207, 0.09753487642721644, 0.004212682085376359,
0.09218422853085359, 0.1021273165787152, 0.0651128417159478, 0.11335669031030542, 0.08655709388947627,
0.08220378956946461, 0.060758148970268176, 0.04956809941210146, 0.03194557369207757, 0.09066843705286082,
0.08973910040697934, 0.2803376982344397, 0.21656487761480459, 0.2659984879547624, 0.2379078996976547,
0.1847211395137819, 0.059005730753119295, 0.07100072016998718, 0.12130357684785953, 0.1209200357603978,
0.1768465936983144, 0.1358195455129511, 0.1684296451090855, 0.11599086890018497, 0.10626811586059028,
0.10256121743154466, 0.12627865618047568, 0.16470814125146085, 0.06946792492887374], [0.37500643698851277,
0.1317223451868066, 0.05531741979570734, 0.2184183935235645, 0.28681017294622346, 0.029277759578778297,
0.1346036452694482, 0.07662622252817107, 0.10862417177576977, 0.005413771144933183, 0.11665106542712612,
0.1297917734426112, 0.3606017334495105, 0.665812375399672, 0.3203845974900139, 0.0930557830640907,
0.07746027059737007, 0.08995872861850925, 0.04210867668743838, 0.31486178383474805, 0.6252518420532136,
0.2601177212907961, 0.3405744783596256, 0.28147768928593353, 0.11243539545810456, 0.08054752512969987,
0.034312931531797146, 0.05663408946434085, 0.12051496000937295, 0.05022852338019873, 0.12026287658272354,
0.19684458285398443, 0.23894072220227977, 0.11148267395791375, 0.03870387595758004, 0.20670473211903959,
0.112477728648437, 0.054089138120094304, 0.282108859120855, 0.17607655602376357, 0.3460671943796708,
0.2593458817787428, 0.0049879664852207255, 0.1357395269367159, 0.18923935500271205, 0.09910412245961411,
0.0412419041198361, 0.15937332668657542, 0.007510144643857955, 0.055508792045220086, 0.1450062542035312,
0.1356604362922213, 0.06640065561221846, 0.016572425642343323, 0.052474402629339986, 0.06402622298503953,
0.011985361830664404, 0.07445239005800625, 0.022711148126536813, 0.007645548570034708, 0.023333228625515734,
0.1062434494633578, 0.14991657590666557, 0.18035809218918583, 0.19083336139665566, 0.15661242456926686,
0.12072023727579853, 0.07292443672426899, 0.013656761564450235, 0.00758407087611213, 0.05729088197277561,
0.15382752900055757, 0.26623855260602475, 0.3510300778446495, 0.2783304901373962, 0.14389411746764416,

0.1246147601652274, 0.08828597437292383, 0.04952774581251196, 0.039585664616291254, 0.018954835924047553,
0.06755595303233392, 0.11006111159727244, 0.15152261905504025, 0.13743425062091533, 0.11650613324724572,
0.1329035572992676, 0.09881362576636896, 0.10299919208457911, 0.047683307166002026, 0.12316662535027018,
0.18764114570896065, 0.0897971173777775, 0.023236762369336098, 0.009907109895279952, 0.0396051493528441,
0.12695830252535945, 0.10060815870130291, 0.03847673550037726, 0.16498704510210963, 0.17147416762044143,
0.16260436886017457, 0.11877436660968856, 0.052828360115164084, 0.002920295518681764, 0.04506168161308035,
0.0005003497158463769, 0.20157786503028932, 0.17873887427476073, 0.28294551586934963, 0.3528773494874605,
0.28088933996715815, 0.14276534676597508, 0.01729936558094565, 0.04116318564842628, 0.025075114268567036,
0.09310398713970051, 0.05044077352712226, 0.03685586871535618, 0.010207955688819085, 0.03987236362691628,
0.047466611622831736, 0.11884221240509966, 0.12686349147385115, 0.024349717679634206], [0.0477347912378097,
0.11352054917205458, 0.011534945642006227, 0.43935824425745706, 0.21235574017137812, 0.24517181631591228,
0.05714335874295087, 0.42141778628366405, 0.0306562985059077, 0.09313728405486525, 0.07167574969180085,
0.10650461051062426, 0.23576488312867985, 0.22919288925716233, 0.2620116355703733, 0.008167522450224743,
0.022220447184590816, 0.1459747059868178, 0.007547441805497386, 0.11481918039339893, 0.4573281980739761,
0.07700104641207298, 0.282353690689026, 0.27934058911175474, 0.13225886539259094, 0.010002949575623287,
0.1774892196236757, 0.11580373714672272, 0.2740340325648634, 0.12828740463043528, 0.14187316819907106,
0.3487338926419559, 0.4199205472306414, 0.012934966694433283, 0.0772423511962475, 0.09490171889449518,
0.022799680280473456, 0.008419904146084546, 0.20181998178589838, 0.15218502529495143, 0.19960616158622074,
0.058450464983247, 0.14516320752516634, 0.004710821514431632, 0.0025454078424617917, 0.023116587016621688,
0.07217898227058724, 0.1863411175336398, 0.01816589574849773, 0.0011741152672031722, 0.07331780764917852,
0.026759279626206327, 0.028548475687972523, 0.03084542802028016, 0.04456900250459182, 0.03446179922680031,
0.017077650508203368, 0.048858318647707594, 0.057689681849186276, 0.011478289907685906, 0.04951122307300483,
0.10320405550147888, 0.16030694047666813, 0.16557538199017255, 0.14405300099537052, 0.13343165470442941,
0.08260157135162258, 0.07250878452009309, 0.05248491905550885, 0.019469319524354323, 0.029680776370299013,
0.03183494984881539, 0.12781473523463094, 0.21035576154667684, 0.16068775944943717, 0.10873546038958808,
0.1370256785976113, 0.18417276756010398, 0.21225998444264618, 0.17539725418280874, 0.1593186147362644,
0.09367940702450156, 0.06376310686378228, 0.09930684513010944, 0.07335981015191662, 0.00817735739134102,
0.011761986547482676, 0.03589742353993387, 0.09405521445691889, 0.19399198751473046, 0.19618459883527153,
0.29331340797490774, 0.12501985807232546, 0.04954619637109848, 0.014938309816864153, 0.047525144354101365,
0.10229142088352303, 0.09236945517694488, 0.006607484476459411, 0.0904265382560659, 0.10789646881227458,
0.15588900165139372, 0.12420412688818529, 0.063796548537771, 0.012317010461021422, 0.08367925516227927,
0.0334637288164921, 0.18090945313751328, 0.16600774059386542, 0.16404558791835475, 0.21667841818376535,
0.1052502781019753, 0.01602954019852732, 0.055364656987563814, 0.05876541828387229, 0.025787822777940762,
0.09212686083665694, 0.045869138731769, 0.020451237394498435, 0.03216118350944434, 0.0348342148483079,
0.015278221883953728, 0.08761403322909701, 0.15863281654374464, 0.1768940266258781], [0.43045451847668864,
0.06374726754902742, 0.12657197215734972, 0.2601336646403163, 0.13154351512098045, 0.17855331227061924,
0.03382959142388131, 0.17347560868710957, 0.04387939161895417, 0.09307086657788878, 0.008100339377093273,
0.18534370358060148, 0.3316730891723473, 0.27750583929343614, 0.224037221544909, 0.032681228158116825,
0.0025027230448814225, 0.011166242086636616, 0.04714690875214694, 0.02388281316250379, 0.5309790336008174,
0.29481365754215266, 0.27363567029216923, 0.34031122627129895, 0.09499907132803806, 0.022679645225538607,
0.1369824284941458, 0.0765028761585743, 0.23759287997337397, 0.032263208233560374, 0.014302345758516181,
0.16653308275537476, 0.2729307351042817, 0.08299689078084149, 0.13427491554945564, 0.3039557977203668,
0.19430952646832791, 0.0024233499545952258, 0.13247447242009158, 0.018077227036340106, 0.23070742445351955,
0.28065317603469014, 0.18989534379277542, 0.18701914688737206, 0.06881963816303939, 0.04262119897694077,
0.01891168274239511, 0.10458552317400614, 0.09037005038622745, 0.09650073027450075, 0.012417233812451522,
0.034126453643653736, 0.08443568023317734, 0.053057412083983325, 0.0513688149128326, 0.07111946535981101,
0.03463457316882906, 0.025269396303077337, 0.04405119372930256, 0.013845638237519522, 0.014756199666018556,
0.09108076037343266, 0.14270577084268382, 0.15856638287278546, 0.14567834445865727, 0.16849150965303003,
0.08709670583646426, 0.010066393872308748, 0.0022541347772717848, 0.022101198587478863, 0.01305405400799241,
0.053969558016782104, 0.11509773063637797, 0.1636201495912384, 0.11092171589424844, 0.07677324062023833,
0.06683564373859788, 0.06876417761264847, 0.04921583328125984, 0.013663520056440639, 0.000318027277648399,
0.07295700437482437, 0.16392845223800817, 0.19792934843184656, 0.18855158343294587, 0.19423362680310655,
0.2046989153628648, 0.17445520412376095, 0.08238787510860777, 0.07740817166635337, 0.15192441501656245,
0.2730557999784704, 0.18559455381147577, 0.15785195524285653, 0.09868833900463236, 0.025149129067652994,
0.008300777309049384, 0.0576833111294114, 0.00012192509656921672, 0.09432742503023894, 0.11132276599747155,
0.1463315724749628, 0.12776599931699098, 0.019101864914357277, 0.06932156206172686, 0.050720863875866794,
0.026726899554700798, 0.24307415662112689, 0.16281518431528227, 0.19248842750820389, 0.2613422787598579,
0.21941539449209635, 0.07875358401753887, 0.024693389251185232, 0.07724390597317302, 0.07895641751842586,
0.08560971554780605, 0.05936644661310316, 0.0627983188292647, 0.013675283765975762, 0.0017474251414659075,
0.026185638039483844, 0.06560505783046663, 0.16359387500433553, 0.10603249193911496], [0.3619315430322874,
0.2251837384122284, 0.12060944999441525, 0.21528744642673156, 0.05764309877602475, 0.017046488679691336,
0.2778145825738708, 0.24399241616301207, 0.059248798878553485, 0.024194593875485445, 0.2541729898271449,
0.028167977029737883, 0.3766122853194513, 0.16208315213536165, 0.12619829449561593, 0.03904452602889347,
0.11546925354493967, 0.07788151453217113, 0.044953637573871705, 0.10676334915097206, 0.3315420924243375,
0.32528340965587066, 0.41591183919801544, 0.5191841365010865, 0.12062544015450487, 0.008919538494141298,
0.22385683796631956, 0.013862052616884375, 0.17311855278542898, 0.023814336436188327, 0.06480519613471097,
0.3107361197433446, 0.30542062244878004, 0.19119046834044615, 0.03346290274182185, 0.10926052820829224,
0.05602498096730074, 0.15864658394327671, 0.19078356523700057, 0.1792822924183658, 0.22205863230256295,
0.2550340224370246, 0.03377375867278, 0.009482260135558794, 0.07408082637161906, 0.07297810902631838,
0.003845161335095143, 0.16340333226515322, 0.10924457377087193, 0.08602346328728731, 0.0960866338605832,
0.02400822586888522, 0.09156164912314912, 0.1465792149947431, 0.13358988476643882, 0.09532422584549329,
0.1289168232907247, 0.05893711413065795, 0.022366998804280138, 0.003540422839600252, 0.014556397368233455,
0.1532158435841324, 0.21536347496800426, 0.22633768500407167, 0.20708995176041536, 0.21586720595280118,
0.1262556120883085, 0.048569467846072766, 0.0039031563046555795, 0.05111092891587654, 0.03391492607398275,
0.07615138266633217, 0.12186914725019239, 0.1381131005511886, 0.11676088601415525, 0.09606155591899172,
0.09419328844885748, 0.13226356121231633, 0.1783950601385494, 0.14772047607062255, 0.16384945063752257,
0.13070269993921257, 0.1303957039086659, 0.1710894539533842, 0.19257270183031924, 0.20239852765884084,
0.18536093354747574, 0.13781167061732874, 0.025050379390251974, 0.11556324441261615, 0.16979789198061876,
0.19704651656452912, 0.03637545563334936, 0.027461809899293848, 0.05671730179417627, 0.07892918382577155,
0.14615006354635054, 0.05067149326313377, 0.026258166626624363, 0.00810983914532752, 0.020032079468147935,
0.021265325446709366, 0.0025676614021095906, 0.013187302294937923, 0.0023670130062050936, 0.12521611942802,
0.1860539895231356, 0.4060206893362705, 0.3515473416344056, 0.3868473218856769, 0.3107497298296058,
0.24898482741931727, 0.016224948310304292, 0.10033967366410386, 0.11212575157320487, 0.11711882251428697,

0.12418086702942195, 0.027674406260756974, 0.003232266487842953, 0.06016028231760936, 0.07488023067368475,
0.03962807498305497, 0.05451379964029848, 0.062473786707074624, 0.05222217483664791], [0.2936538150741012,
0.09103417303537596, 0.02105535808780042, 0.6380987964577911, 0.1854598235018939, 0.1895712730899358,
0.020170215495122195, 0.3471843725826979, 0.06730132532715248, 0.04025214822859688, 0.00752948067247783,
0.2629204960012059, 0.12964133907497855, 0.36039522510435046, 0.22351730770689346, 0.10718443533393239,
0.03283341415413405, 0.0408189559646248, 0.040497732943979715, 0.20143277939767718, 0.4520740844601083,
0.22577840418264125, 0.26865742051840075, 0.33324445637043404, 0.10821320891232908, 0.03876776005293117,
0.07628784861341027, 0.026952234122721443, 0.26618106676373593, 0.10439159020428228, 0.02140302437943327,
0.25092604524756723, 0.22907642900517386, 0.05104847313978066, 0.14676058784906684, 0.19183219080272762,
0.051162053199062564, 0.27189184401166955, 0.3074142343294538, 0.08483263774256049, 0.17313276043338424,
0.09561479132164434, 0.07160731681973975, 0.029538221031712913, 0.13808577537332406, 0.029236862855140468,
0.053586430751476365, 0.18669030595359695, 0.035163216990721564, 0.0892424592308007, 0.16279833951461237,
0.21219455353394076, 0.11805452763034148, 0.030665213090539666, 0.07656117167853921, 0.007716923369518391,
0.029013355126414325, 0.014640594063055996, 0.04034962545750783, 0.038610007061453316, 0.014181910329424598,
0.09630418253690641, 0.15017692833280122, 0.1625432899331968, 0.13452260878844485, 0.10894087058621219,
0.035904592084217035, 0.012910892080736813, 0.021639042675986042, 0.07138353800607518, 0.0734326108027422,
0.08972955441629761, 0.11356399086848724, 0.12288764830230249, 0.08124005427368494, 0.04274570134417236,
0.014011421488926633, 0.025658908691877307, 0.08690994170111649, 0.13541360410053574, 0.14629580159947295,
0.13404218132758247, 0.1642076047111204, 0.2000896640152938, 0.17394614692956428, 0.191213250716 6418,
0.24637318964923072, 0.21123241910700183, 0.21137575287883278, 0.06366029379905262, 0.05824410839740799,
0.1458291083248236, 0.05825787293523193, 0.05782898812601107, 0.05000183508249956, 0.02439990394129941,
0.0015818662384058145, 0.06871337207387614, 0.025363189995653138, 0.09897025052845107, 0.09277308870033975,
0.03793185321955034 5, 0.0122736310902027 9, 0.1332168383796691, 0.10172452572143274, 0.09858239027610025,
0.16879554650521916, 0.32413664214734755, 0.19835481837110627, 0.22985629478362007, 0.3654924879250437,
0.22738894588115452, 0.08860107589734496, 0.06074829335747373, 0.04009939360846199, 0.017658532174974515,
0.002704987894767697, 0.02054404246620875, 0.04120901053877362, 0.10078373013004209, 0.06899167843607315,
0.033742095858891145, 0.05933627195254159, 0.20766154053271432, 0.25037583865161833], [0.24317016202200184,
0.2228240102013657, 0.12742540835780808, 0.12206182117134118, 0.17490455700581978, 0.30239740561686484,
0.14715790517504485, 0.04036013021505818, 0.053254617913989034, 0.1331181422221835, 0.20976524025382742,
0.15235119437926425, 0.1568755839427257, 0.4101127618881174, 0.23334588832976608, 0.06553198523406131,
0.077838163529199 4, 0.014098387748143697, 0.1305394080225763 8, 0.11960767304330834, 0.34253032466238853,
0.26496781679589904, 0.340838976596786, 0.4259258268436172, 0.03258461138388811, 0.07394505166655264,
0.10742199944987287, 0.024458180264186596, 0.0777937416060782, 0.05508879322393178, 0.026327253316494253,
0.14150885947364872, 0.20995591379294856, 0.13417342235444035, 0.08725088976340688, 0.2562344110045414,
0.2050070060164656, 0.16734654870555352, 0.20397673261541588, 0.03409511190165228, 0.0862377699047937,
0.09718923994695958, 0.04070332970389166, 0.11582120529464039, 0.12079609006942364, 0.05152203936617933,
0.15726059114987304, 0.165353091444087, 0.02017678701430852, 0.09576839954680251, 0.040760379297787315,
0.04584924155865498, 0.07767081610267712, 0.04016921786807666, 0.04384376492771096, 0.0416101489033133,
0.061295237894684655, 0.07892614155309927, 0.09996630665093857, 0.10100050247017736, 0.12032264812768745,
0.18072489659162894, 0.21409031422904004, 0.2134340454073444, 0.17783706532535123, 0.15293842433750232,
0.08659229429025327, 0.021116253142889382, 0.056302589144458876, 0.12467391691918643, 0.14317829809335933,
0.16962138873100394, 0.1681743155392969, 0.20131702623242856, 0.2143634613528364, 0.21253691937839878,
0.15997728889488555, 0.015181671428372207, 0.21067542230678882, 0.24819440304681867, 0.1045538487289416,
0.07439153122695354, 0.19456295939230292, 0.23246506961793975, 0.2466480166119183, 0.20381835113611196,
0.16577156332963622, 0.11952056067849773, 0.024109373882871737, 0.11190672769623082, 0.19259501389650607,
0.1806220715988236, 0.046862688488090304, 0.025625022802180 59, 0.06345530936321116, 0.09936450128126872,
0.10732397434419347, 0.06571205891758927, 0.003272676677449102, 0.054086689784841505, 0.08352123337213536,
0.0821390668410404, 0.07281552668480841, 0.06131015078896421, 0.07973827221961508, 0.13771829508667743,
0.13960536661529357, 0.2729881565221132, 0.20178870163591328, 0.19184127383639485, 0.14607143394529862,
0.10527542617044534, 0.020690149500899823, 0.07575964105307999, 0.04339416202358496, 0.008063036522940248,
0.05236906327770384, 0.014876094546776476, 0.02684238605352313, 0.06377189746439263, 0.07645075836369797,
0.015112078047750725, 0.08452270160746272, 0.16315404655414886, 0.021210628100650258], [0.2694688623048673,
0.053678648338893194, 0.1727190187365103, 0.23241742011899993, 0.01786568734430971, 0.029701615812849944,
0.05848715172079257, 0.07427917445295164, 0.042488386003861976, 0.1557931180378551, 0.06524549656023981,
0.016192412796373106, 0.26876766417209047, 0.2912153606175075, 0.21199154968682493, 0.16321856651525063,
0.17749846170596772, 0.08574995590104831, 0.03237589481711453, 0.17485619523171003, 0.4731055369071691,
0.2685317732038894, 0.28992967045017654, 0.2489778929863478, 0.11703597734764057, 0.11650508130071122,
0.017051201419252784, 0.119733657699501, 0.10138428659013296, 0.023224937982086427, 0.05788855770200022,
0.12475365893521514, 0.35425168427828707, 0.0027870558786334252, 0.15904318585719618, 0.2695991765375309,
0.18312527045765786, 0.04185148189211922, 0.21758424820557756, 0.08410830022043564, 0.11191465290135838,
0.13504586093436521, 0.014014764146148342, 0.11755651740099045, 0.03799200787542198, 0.02818838286255443,
0.089059105798053, 0.2473344310669972, 0.062002808099462875, 0.025621399015925677, 0.016896505703798307,
0.03298430852074404, 0.03668679575884524, 0.09447993635527906, 0.034719396085617776, 0.01658424308703465,
0.0062996383673634936, 0.0020737206901924035, 0.05187431031383689, 0.046160995525838626, 0.010106832891749072,
0.10551154311214184, 0.17496659742603654, 0.1845043826149839, 0.1823614737499312, 0.16110826235347495,
0.10785107934196297, 0.05962492295011468, 0.060131524115936585, 0.026294235606475307, 0.0037554930107295895,
0.030202464180620302, 0.09858444245338488, 0.12681056098779575, 0.07717818141270327, 0.06604125317732804,
0.08074204213585089, 0.10308737610719092, 0.13631999775541617, 0.14645586335444324, 0.13974497722912158,
0.1548717079971906, 0.15263328036014806, 0.1445795111996741 6, 0.16975016459511816, 0.16482116593279272,
0.1585133488862827, 0.12259217045497646, 0.058930101658935884, 0.0631508774652827, 0.14172523393891023,
0.14619027494702083, 0.05873027749010688, 0.01891979403341223, 0.01681249324289963, 0.019323244507843933,
0.036008227835547434, 0.04117822858265976, 0.030609568819899367, 0.019457922640183666, 0.015950838765484587,
0.006642794856889144, 0.03208842472160479, 0.06538364250282956, 0.023260011554656342, 0.2128576736675237,
0.26288701646578205, 0.3871122385166662, 0.29028727195229587, 0.23220779238102993, 0.22987179496905158,
0.05924055912373402, 0.036564119162574106, 0.07526113563608555, 0.06894284992434593, 0.06863860584626949,
0.05904517600814378, 0.016110254145193438, 0.023973319856928705, 0.04120373145938468, 0.02411201062097578,
0.0960442180761491, 0.01388221030993816, 0.15583512036286398, 0.3055738996839937], [0.3288589567032075,
0.056848235573031286, 0.05432732826658873, 0.3067939922439272, 0.34114563007945814, 0.32562073761608296,
0.07748324531023851, 0.2964941402531103, 0.20648710458227126, 0.08935509881451506, 0.02612176328727769,
0.2777690586856601, 0.18243469268045232, 0.3684047390583268, 0.2583636504544888, 0.04076043780358931,
0.1961994296162394, 0.14333478810168968, 0.007510537579508091, 0.2978217251880141, 0.4571122653345687,
0.3293589455803628, 0.30095109474948195, 0.3588606422515518, 0.18510851730968236, 0.06711024307005783,
0.10901207307469014, 0.039770630482534035, 0.09152429925545856, 0.09625293233656312, 0.02905283814344616,

0.1773420543032499, 0.2649064688986789, 0.03246362930372221, 0.004310711751768818, 0.22787642545413672,
0.0996173285744052, 0.03206272183281594, 0.22360668779274073, 0.12481726768187583, 0.21674082457401136,
0.2015268274081343, 0.01001418424425275, 0.030701337867288646, 0.042143675417056346, 0.07140610776950555,
0.09587377400464334, 0.2423502317266096, 0.04007101992204616, 0.030187850143905647, 0.10166449504221445,
0.16368424655958952, 0.1906791496159564, 0.12079792350863412, 0.11250343375545568, 0.07043308628842211,
0.0023700517242915453, 0.000799245500404599, 0.028234044112422144, 0.0001671312569603039,
0.028114876349818404, 0.12150276413764095, 0.1779161738437902, 0.16760439305189867, 0.17047675006233984,
0.17282894566869125, 0.11676773831920809, 0.08147460898698942, 0.06235172799881161, 0.01461713546234147,
0.021181756195307426, 0.040835582672306345, 0.1829072414901069, 0.28792423590911903, 0.22750615426663684,
0.13884235524351426, 0.12362974021257399, 0.2011817282095237, 0.2782537535013595, 0.2438902779106029,
0.17317671250121083, 0.14004214556203445, 0.16555286546643325, 0.1966395018133243, 0.16503126735719548,
0.16505003244555036, 0.21476691540191273, 0.15891242758639545, 0.12478226817979417, 0.05644989225874627,
0.10809165663234316, 0.2031210280069204, 0.10323864236976953, 0.08183776066121612, 0.08190896648490184,
0.05966024201812368, 0.06837504807064852, 0.09414433116262448, 0.02184072351792902, 0.16436419490530138,
0.22331208093791394, 0.23637669000293712, 0.2243423398258372, 0.10776953226304037, 0.022555004802297253,
0.10564739757136211, 0.04538164164641583, 0.24756107471983047, 0.26977197043202217, 0.2884054641819721,
0.3723002504359616, 0.277220186845315, 0.18098269293540722, 0.11734570236024029, 0.10214868173312738,
0.07874038156158554, 0.07981042053728754, 0.07840788921533565, 0.06405691049375944, 0.07870648582084414,
0.00915117475771139, 0.06247688574502676, 0.045058814070932963, 0.19304323026943146, 0.2118152374546927],
[0.2379218714044967, 0.027967500886012946, 0.23643399931741552, 0.5576480560376257, 0.04866459279150316,
0.26183923613511706, 0.11280662828043628, 0.2705609091585868, 0.13719120940553675, 0.12373073988158279,
0.026456639347654624, 0.12644705202189221, 0.3189052309266564, 0.2750786230266866, 0.3591058409894853,
0.054036115613203504, 0.08987189458227964, 0.24077735548454052, 0.09834164667581505, 0.16590988564218775,
0.6091256281949456, 0.2504836373496142, 0.3403464891080985, 0.35997441957340093, 0.16127955474670316,
0.20911262526487379, 0.0733684150430295, 0.030186725274837966, 0.09122593681400748, 0.07883871535599343,
0.18265248934323375, 0.09363058861971247, 0.35963725117161754, 0.12255788088549979, 0.01938620096710577,
0.2118417630891234, 0.25618885833393024, 0.09123422142763889, 0.35100630410867434, 0.19813297184649253,
0.2393574904811284, 0.18176796226635827, 0.12469672174126388, 0.08206230821135581, 0.019376465685826013,
0.011517807771081811, 0.0893106488953274, 0.19029943683683484, 0.017649386288185798, 0.03072805743349228,
0.031741954577029316, 0.024488070396450647, 0.06150525127327385, 0.1632194477604212, 0.08159299499762714,
0.03999184102863555, 0.12832419517904875, 0.18655687785555622, 0.004901427796036055, 0.012248170218206535,
0.0490324082932145, 0.18806235900169596, 0.2012221429150503, 0.23651705150316413, 0.2505198974679941,
0.22847653050779218, 0.15361638120611631, 0.10752918725376845, 0.1270302248840572, 0.08752543798909701,
0.06628812721012001, 0.008687548842335684, 0.09811871958524895, 0.20118864541334994, 0.1605689108307172,
0.11385741164778897, 0.12263339253074133, 0.1280271842926824, 0.1785196963664682, 0.1673476057670857,
0.20366795694941717, 0.16220405088725706, 0.14083990748457406, 0.16179081156165626, 0.12463286750940622,
0.11937788673134841, 0.128180233080141, 0.08866603400925945, 0.017073403772886644, 0.11981782278999738,
0.1885356796259628, 0.24610603058989386, 0.11357218203237927, 0.04968574102586143, 0.017102971583980508,
0.07671841242548907, 0.03366769807521057, 0.08630611927205445, 0.03227803348168845, 0.08505143616671625,
0.09781976011653343, 0.18092344413476025, 0.09351365137387496, 0.013163486670508902, 0.022537177127122606,
0.21551851331691552, 0.22772899061308083, 0.26658167135317534, 0.16348584163329488, 0.16060924012423158,
0.18561265368646068, 0.02441091822930409, 0.04466179926653959, 0.07314116899493478, 0.0797841847007282,
0.06648106214255682, 0.024160044124815303, 0.054923273461343534, 0.1217223403837837, 0.15722097749213687,
0.22125782714689013, 0.12713772815394875, 0.03859464175658356, 0.15091696717378994, 0.2930210376368566],
[0.4276547729085612, 0.2750015738121262, 0.01639916704822854, 0.2921152655381065, 0.24991270968353158,
0.2418643200357823, 0.08224898219271393, 0.2042892433265553, 0.07815650651728383, 0.1109518727239686,
0.07311439851572511, 0.2514260917257202, 0.24934571841447395, 0.6001768358881799, 0.30915933323324574,
0.07312609766099457, 0.014776479146008598, 0.5488369261895937, 0.05510422117219346, 0.09632886595023471,
0.49532385447855914, 0.3133261530871496, 0.21854253234930962, 0.25138409284863683, 0.0522003640371718,
0.08564058807138673, 0.09806163008938454, 0.1430536446098944, 0.261654963364467, 0.06302887064846492,
0.026644133112103622, 0.13135948116833865, 0.27365771808132644, 0.09676444348555976, 0.15498284434562806,
0.16878345696916056, 0.06086168071043686, 0.02023528530244152, 0.1788957827961144, 0.09996937973767425,
0.40988083026148575, 0.31987108086760774, 0.02844646422312737, 0.02895274347474961, 0.06433781040182658,
0.07834500184018671, 0.08435083389055135, 0.13486122742008283, 0.021430218667070112, 0.008075631966067632,
0.06788646238590648, 0.08815751473747163, 0.010458395168603914, 0.06733147669706083, 0.01679827984967293,
0.020564664674412904, 0.019459292137529876, 0.08287747051256389, 0.06318644890697504, 0.14282978076806235,
0.06580542260044825, 0.041195465641136415, 0.16068995119924653, 0.2260814397546035, 0.22278997250826416,
0.17317782300647658, 0.09991249768487923, 0.08510221108042881, 0.04745439751416296, 0.008951934242269493,
0.009465415895234489, 0.06875213854404663, 0.20261452680026223, 0.3352838160254375, 0.25540700218249124,
0.10211141951178791, 0.07134236390166696, 0.04916664117185905, 0.04341522180255769, 0.08222244297932971,
0.1132657303711027, 0.045665490960977725, 0.04859728126639557, 0.12993951741591628, 0.18547429938491256,
0.18460842189767399, 0.1493338314627845, 0.129053049040447, 0.0746112706043936, 0.07064068781313078,
0.21067224588836433, 0.2956697734951343, 0.19910021476765058, 0.1830576374616292, 0.12861499343720534,
0.06642677232993949, 0.04375500710523351, 0.07377721647625593, 0.029920266701850072, 0.17078400269840877,
0.23974992402480033, 0.21157207472749706, 0.14599584612412744, 0.0019032983026254708, 0.0937680254980937,
0.08233009521748368, 0.014489474866191352, 0.16944029004106853, 0.14516076593004074, 0.18745273275271815,
0.2568795854007281, 0.13836779147965023, 0.08879792892156681, 0.0023082661573973805, 0.029542689720191546,
0.02413051495406501, 0.0571993102102987, 0.019472606539248143, 0.043363271514088975, 0.04236247664524596,
0.09316163524217468, 0.12851352103656982, 0.12929904784198204, 0.2366761002542261, 0.2593370668741461],
[0.3841125487524191, 0.05004185572403028, 0.00728428058763674, 0.351846268734795, 0.2502455042414075,
0.19495180299222856, 0.21999613781513636, 0.11619238573873067, 0.03534888702927021, 0.024416439423567637,
0.009196184821807703, 0.45822213197029027, 0.1958559140661337, 0.30738334264707795, 0.07986162188778727,
0.04749577407081994, 0.23995027625055232, 0.1341420770360638, 0.27489322105835445, 0.17150759595065626,
0.26948809607237373, 0.26018193649016763, 0.44096245268688394, 0.5421503044414167, 0.09453111500884154,
0.08092450446851021, 0.0419751780143278, 0.02979586772955192, 0.2604335931552535, 0.09109459454465875,
0.1439469144943822, 0.3048621082104228, 0.23865132513900478, 0.038141654193928, 0.04379413331624862,
0.12413356858493632, 0.10209941653157757, 0.01814931795375863, 0.17166512852365307, 0.1980240449278707,
0.2052620106483934, 0.09572328711514934, 0.05111763950624801, 0.007535111400995512, 0.05284536160006966,
0.038313983618922846, 0.15406576904548372, 0.1529875622338684, 0.01384989595796743, 0.0090052454212332,
0.046417016856362854, 0.12938087132622525, 0.05771322098564985, 0.0001456162464377961, 0.05016491674910376,
0.006663545343847814, 0.014702827696215772, 0.012000113007574921, 0.0568391330498998, 0.02511787399650883,
0.029528209717803754, 0.13069279821730478, 0.19932429772518734, 0.18775862590772274, 0.2006873699667572,
0.20544527968222243, 0.12379011506200362, 0.05291886587389536, 0.02124568469273436, 0.034902857868828165,

0.021258116131361977, 0.06152819884642307, 0.19046922142797235, 0.2679569044060404, 0.21982417336820823,
0.10776373053904244, 0.0774244716282013, 0.07344545767630178, 0.055000071201501646, 0.006002324270031431,
0.034747411355558804, 0.035095442099629774, 0.08166132649705245, 0.11367280305827675, 0.09914656145489768,
0.1257615858118808, 0.15067922478025655, 0.11556935276500536, 0.106722539958473, 0.007912230033954576,
0.07283727932917339, 0.10975857194955731, 0.08720397668836782, 0.04983760682122111, 0.01394593406652059,
0.054628898390164854, 0.08451296431786956, 0.04524422139906226, 0.00642695474564263, 0.09867520566872742,
0.11763842533209615, 0.05670710853438331, 0.02792338870000044, 0.029777263474247594, 0.0842097431890592,
0.04943879195093634, 0.06421225992755264, 0.3035467205084836, 0.26044157733004236, 0.2651310835684205,
0.2381773481535958, 0.21913731885144652, 0.12348360761040515, 0.01192789183785128, 0.014672040170013688,
0.04642849381665084, 0.012154623327383934, 0.08535597436144732, 0.06444205857473409, 0.020669114605661832,
0.05244647436532795, 0.08628495424803072, 0.06611337016675983, 0.3022499971880725, 0.3283551886376492],
[0.242757546120467, 0.031473322649079705, 0.08984578232986469, 0.14852740606388226, 0.06573864427513568,
0.10287846078687035, 0.05792062581334367, 0.0019153863597465904, 0.19183803676995542, 0.012850914305425397,
0.12716989844473833, 0.11247186765720073, 0.31645264724847155, 0.4395775024800375, 0.23270529694252343,
0.16828124218139048, 0.04993149604230264, 0.10961494599302864, 0.025725562974103063, 0.01124964550858031,
0.3605931596024833, 0.37989913771888395, 0.2353608374043027, 0.3066127956802605, 0.05034146768475259,
0.157202080647522, 0.16185616768021965, 0.023684515071880945, 0.17212156432023226, 0.0460317082621938,
0.016379034843675266, 0.0905580826006243, 0.3670932361179772, 0.18560603003448622, 0.04162105658104623,
0.06100116338013014, 0.1403296753880943, 0.13110019100203077, 0.28341533729520174, 0.1626571250753679,
0.2428453076708242, 0.18157724871051695, 0.022736619694796872, 0.03896220148077966, 0.08034899163794577,
0.044883592127970336, 0.010377967298306343, 0.25732860986689876, 0.0318569382573082, 0.00020468551964257487,
0.07035298103049624, 0.041415521569369104, 0.060064505313084084, 0.11599978995790561, 0.0797297927988936,
0.022387736243822225, 0.035470809219046356, 0.018862687065651702, 0.0010285910380086526, 0.03222058280705979,
0.001375421922445587, 0.10394608635837506, 0.17245104336161812, 0.20075674698009854, 0.19790254793186382,
0.17716841570390543, 0.07507589376287582, 0.021664483514836255, 0.04720434293068833, 0.0926559480212921,
0.08239339109379669, 0.060165249018998425, 0.04796556781670376, 0.10104358790584114, 0.09453457469618023,
0.06508371967775475, 0.07361422608018635, 0.02637788189348389, 0.03958188382434861, 0.05849021148296449,
0.0007406816180771829, 0.1149795856065953, 0.18669864511662423, 0.2544569173871998, 0.28006966471519373,
0.2838627909163919, 0.2721995294510645, 0.23571123786137876, 0.13492856488941474, 0.010271908740748308,
0.12090632049512705, 0.27236203841667594, 0.12871797791524012, 0.05974645404249065, 0.0008543025723278235,
0.04091729798677436, 0.03003061179229346, 0.02918204721913742, 0.05477416405909022, 0.01034691479984874,
0.07370832937333516, 0.014472428873288411, 0.0987782702319341, 0.2437632541389357, 0.15955773310958002,
0.09419078227321136, 0.16675314767646865, 0.5240631542664264, 0.39047638887318253, 0.3469825355613872,
0.3316522247795604, 0.12317502789273745, 0.03277812186259333, 0.15102973407197545, 0.13007655130426637,
0.06851806365208599, 0.10541307349225731, 0.02156588642984869, 0.06319081363412409, 0.08777192663133397,
0.03922635678197315, 0.0013828533842086981, 0.07927526097446123, 0.14916603529995542, 0.16505318122778115],
[0.27322322079198824, 0.14101704655409145, 0.16596353484340354, 0.4159719478253137, 0.21252122072841118,
0.4388331930533757, 0.07317977406426038, 0.2369677939766343, 0.16624590150842328, 0.3054216647703055,
0.21330753339741154, 0.16749611299706796, 0.22045135177574038, 0.36571152318757083, 0.41197838842036977,
0.05842548747466569, 0.14010958228969383, 0.34203622887512036, 0.07713549194026466, 0.018912890017058634,
0.4720295045968844, 0.3208340633333835, 0.07527010384815623, 0.48383385293773873, 0.10437541410039841,
0.06550728334297208, 0.027915495067453857, 0.10396235801414, 0.21996410712811645, 0.046700927138821895,
0.15393117655988234, 0.1604571847359204, 0.28246115000718525, 0.06956577220185026, 0.01919256621898269,
0.07056569855827918, 0.026288259379857404, 0.03329231861874848, 0.2876539148664131, 0.0755307321595193,
0.2896532004939397, 0.16058591318490292, 0.1356327562504987, 0.09184481543234815, 0.15223177219573178,
0.17586736381405735, 0.15698602887220942, 0.20005469373978654, 0.03441320416546786, 0.10983263578446051,
0.03213457596690158, 0.07816392035316759, 0.08288151968331157, 0.05376647565126566, 0.00012995089754148294,
0.010077991616726687, 0.008807729264462773, 0.08019116648374383, 0.022831179253016816, 0.01056187962328515,
0.03833363274070965, 0.13051679247598985, 0.1518734808852542, 0.14312232641418524, 0.08750175067027825,
0.11727816615320782, 0.05257502412658733, 0.015250108282151316, 0.053683859724328285, 0.09331086015465515,
0.07717356715941995, 0.10163218150446222, 0.13503262705952856, 0.19182771930252018, 0.20379312038562405,
0.18009208041783295, 0.10287327790540604, 0.054114657224740764, 0.008004622437958008, 0.014336656095985195,
0.06059884317476513, 0.026571823906491855, 0.10874356560916286, 0.17852813992758018, 0.1980564854049244,
0.15911869382460764, 0.11613609680454572, 0.03566342054070555, 0.029378165090958565, 0.193089704089889,
0.2725555170272381, 0.33749809551795834, 0.15876787456782696, 0.06302775926909111, 0.03564887383196888,
0.058823126358510815, 0.09812622386095021, 0.025575133099139405, 0.0957958433629672, 0.0243609298126717,
0.11705642494118818, 0.23885638948196156, 0.2470774566006092, 0.1295832683902146, 0.1500661722645649,
0.22691172810677893, 0.12799026312636078, 0.33689936172402124, 0.139293703622025, 0.0997771108484443,
0.04316619625471342, 0.05036610248234529, 0.17429199821884075, 0.216496783291571, 0.12713942342072337,
0.058577941863939643, 0.026521759646374078, 0.05377325970716031, 0.08017424898126435, 0.13228668194272664,
0.07277455769904367, 0.07048481551137362, 0.18859901932582693, 0.2099663127586081, 0.10647940397729526],
[0.266553327253711, 0.2326446384532793, 0.03894582363252406, 0.3377984009402854, 0.2661279954137487,
0.241054211552816, 0.03425107604070565, 0.2061381674049463, 0.24715525980358682, 0.0415029480933261,
0.13067927646257702, 0.08307648172934251, 0.18525838091718969, 0.3452767515486027, 0.25396850590255604,
0.046410283718094074, 0.11941164782937955, 0.05821464624412017, 0.03844327543529616, 0.08572529699839489,
0.39751211349744703, 0.3425065317649887, 0.31802952996401335, 0.32471201786794157, 0.012619047352882518,
0.112736616811413, 0.10032607166354178, 0.04352861807981163, 0.2491269641052933, 0.003534245493538613,
0.10361130099797286, 0.272304514262276, 0.217914436226608, 0.030297016538144907, 0.011369711599572905,
0.09050625075703336, 0.10100726713225665, 0.13886396090860237, 0.25354760830963596, 0.15205908125144926,
0.24362689178057909, 0.11937857664279719, 0.07228634353735014, 0.014605989316568369, 0.08454082853840339,
0.09163663690203326, 0.09936766873998563, 0.23106817522163037, 0.004036420240447715, 0.007367295582443254,
0.03863051525623762, 0.03660696317406037, 0.01326603360000476, 0.0037519385685667508, 0.0162027688960178,
0.026161826937071784, 0.03245074341935911, 0.030731157422934, 0.07367577893854756, 0.06375721448550876,
0.09399110443854385, 0.1318802591266016, 0.14522478775514575, 0.15679826757218138, 0.1508865252806636,
0.15554888086656954, 0.08552412054861365, 0.04291166766354811, 0.0025772263937009115, 0.03899040513515457,
0.029583429987749002, 0.08115829234426221, 0.1324885762507601, 0.18224822559590642, 0.1295517367886762,
0.021952969856694256, 0.0025990993037474275, 0.04360324666920266, 0.09889482650568387, 0.07855907343474791,
0.12572359812835038, 0.10163566782138103, 0.13166445404322993, 0.15722662666531512, 0.15595234307372938,
0.10563911815655211, 0.11979756361117978, 0.11955160106725031, 0.0025240686400739955, 0.10524488806605571,
0.13960725306724325, 0.24626193548880881, 0.184222372881149, 0.08579315853530273, 0.04492925959586419,
0.025266654375050858, 0.09198824728988891, 0.0838206193758987, 0.03342626385727021, 0.1152670819155127,
0.12172305092958534, 0.06201732119840687, 0.011361876848293335, 0.014242483190758755, 0.17222020155547726,
0.0028876174558845973, 0.05220956626729318, 0.2290755934552727, 0.20100210587868153, 0.2699896488632189,

0.30819576711245833, 0.25356754595577785, 0.13042842744660357, 0.0021000719731160047, 0.06978123161420005,
0.03196814314219318, 0.11532938621952399, 0.04379222359260829, 0.046257335959675815, 0.006151052165403347,
0.021153367212197374, 0.03354690020214233, 0.019473257423525087, 0.1144619766114224, 0.07784757451619466],
[0.4775693718802198, 0.34181629387970464, 0.01746091195568012, 0.34158519475161303, 0.18999266441097531,
0.18186333147031525, 0.04714049390792346, 0.1361999151442889, 0.02161579701908048, 0.07754561822168328,
0.08756233128695733, 0.18634791822205898, 0.1075617374655442, 0.4001817751663062, 0.3495411987421661,
0.03889341626086744, 0.025383336606558064, 0.24075971952895384, 0.17834859102473868, 0.20706811894093713,
0.43906092923502715, 0.26080270497323993, 0.26367675333376744, 0.3267152153220658, 0.024132845206834928,
0.02752766230085654, 0.0382352612286873, 0.14748090891909893, 0.3590218723163883, 0.12092980271184676,
0.1350049522193008, 0.16833524693604968, 0.3087111016399012, 0.12411400111976352, 0.11083967725113443,
0.3020590689050705, 0.01838056866946641, 0.08172688655042558, 0.25721299921228125, 0.19296057468593417,
0.2194316876408329, 0.17597829462628437, 0.05249032964578481, 0.008831042260649246, 0.011765764111267984,
0.006268356299545066, 0.0501942029412688, 0.17548750850443867, 0.005574833977311188, 0.018984722564220284,
0.06302791197878978, 0.06356849535682053, 0.044205566837045136, 0.07258306064617363, 0.03122795179711994,
0.10907983649958267, 0.07194012549913542, 0.09749501699366411, 0.022074181075189103, 0.06626362568125566,
0.05397914314803664, 0.05631312647868661, 0.12390949769250396, 0.1808387099676735, 0.151614388878484,
0.17319576437902356, 0.1164838471016472, 0.03326732049069184, 0.010200575438165934, 0.017275651792235627,
0.0046305636742749075, 0.03308750796942284, 0.06483699886286486, 0.12523329771442643, 0.06973305819444506,
0.021381911304873564, 0.0004897465262496258, 0.019759883294707906, 0.05090992829690047, 0.028682238381706887,
0.01559120329698154, 0.029875640226076937, 0.09288859736604246, 0.17294634332941303, 0.13021472094679817,
0.14775229918304966, 0.13807485323488594, 0.08279658999836316, 0.0009749866980208086, 0.19048872967605313,
0.24248882896308946, 0.40291338607846106, 0.24198299409105278, 0.19384605716776, 0.05329117740723371,
0.041917808797808015, 0.06822346507881213, 0.08127349695894102, 0.005071254846990211, 0.05988271635513273,
0.04442999226230708, 0.07588254708093725, 0.07088698381772071, 0.19076158388074896, 0.11902418477629088,
0.028802137981628815, 0.1387163980462214, 0.3601035917311159, 0.20910541236219654, 0.23377097210682266,
0.28089939042502965, 0.11371917812032012, 0.057239437607642066, 0.17847182988050525, 0.2251989598278731,
0.18515241485386058, 0.17505993726574978, 0.07718330900746855, 0.0679272207420133, 0.059312797086974525,
0.02560422124630845, 0.053650544489411485, 0.11664201541475631, 0.1623516679624917, 0.10041060397249138],
[0.2249029443440438, 0.11878577711960774, 0.13796367511264362, 0.39594715025187543, 0.24239324788235583,
0.25344031122481125, 0.025007429141968483, 0.007491839273401632, 0.18196853975708066, 0.16067160372531106,
0.04371214929162355, 0.17370800849350232, 0.21561767309988703, 0.4411957882130381, 0.3081068707231516,
0.0829270500066182, 0.0009676326334438562, 0.12330289008817577, 0.06977320812761119, 0.23257666589584308,
0.3499480483666956, 0.42522866692777195, 0.25718640410522925, 0.509087454574985, 0.1969041374877131,
0.06718773043499805, 0.028737319986885817, 0.012114848728585485, 0.18145555253687196, 0.04490207873357689,
0.1688213543597446, 0.4053602693299153, 0.36720999166214824, 0.06269074962705104, 0.03809865244574139,
0.1110162636682385, 0.05719682096132536, 0.06694937494044013, 0.18532163491337383, 0.024075620402103676,
0.15337826332589377, 0.13359101841788504, 0.04154300331008199, 0.053034500457489066, 0.19013513533429183,
0.13596337200570685, 0.010459436342233307, 0.18875023236918595, 0.004191501223535416, 0.02095891570374218,
0.06171150714359199, 0.09667793816077476, 0.023921187549357535, 0.037245175091185447, 0.09692199994535394,
0.09364221920250633, 0.05940584461894113, 0.07826650792522766, 0.0878935469232624, 0.04476074808760376,
0.07055320487380833, 0.12533420527350395, 0.19487605706219846, 0.15394055408493748, 0.12963887073292416,
0.11549533305269685, 0.02533777235910324, 0.022449179682996687, 0.016569373673989436, 0.05096353734420193,
0.050317743376266405, 0.08747163838006627, 0.14416565618153687, 0.1889505980229174, 0.1631693124129907,
0.11459161792637079, 0.09354510780259463, 0.06374907328736946, 0.019340838334177688, 0.03397650233292435,
0.02918596002256929, 0.019126958765458963, 0.08572265737646134, 0.13826857417554497, 0.17549020166547033,
0.16557977648606156, 0.19301606093273038, 0.16971484341803478, 0.09455786055278159, 0.05244032414764725,
0.14053642291207083, 0.22121439839716175, 0.09588630162347626, 0.03396707804106503, 0.02950364243362357,
0.07529822944817141, 0.142230684585982, 0.09730902168479637, 0.004319741270425447, 0.050934468091123515,
0.0438847063081467, 0.018720222406151076, 0.011339001003191584, 0.028314302504465382, 0.07611989705802477,
0.08647181421930125, 0.04730072217596254, 0.21224733116488884, 0.1458727629061881, 0.1989773073152761,
0.22291522614362147, 0.1783970038381602, 0.07973115018062289, 0.020881304695908015, 0.010062157274731245,
0.019617953702632918, 0.02581351196161045, 0.03462994571872826, 0.07346515346194973, 0.11528274689722064,
0.06708722700497646, 0.011792045693251535, 0.08277658683091714, 0.17906881137551825, 0.17078596522941533],
[0.29421321049832966, 0.06781737780284339, 0.014056138355191901, 0.43750019541536345, 0.205667121522681,
0.34449118295476133, 0.024751827190513317, 0.03413736890200051, 0.0018865008747388486, 0.04398999098440951,
0.039404017398900176, 0.2997177817430856, 0.09071870090361393, 0.4268698099968741, 0.39039223152802766,
0.28914246330577453, 0.07821967949849921, 0.12498345405313394, 0.03181444092581267, 0.1584676970241594,
0.4805176725949141, 0.3233190514650939, 0.32909165016137926, 0.32598576892097125, 0.2023544221862961,
0.09074019500748338, 0.030930009449461122, 0.06237404382055046, 0.2985939156769487, 0.061122376027159454,
0.007022230726563608, 0.13834738454065112, 0.2507219430602192, 0.1913161943570077, 0.08897391554722309,
0.24193409861631318, 0.060089593142602905, 0.02318167736962681, 0.06985756366269233, 0.05489671111961756,
0.15524332070800873, 0.0884802344380554, 0.049273849877754176, 0.0008440730580024752, 0.0653085526892295,
0.06930751040139399, 0.01588705038765324, 0.1625661807339925, 0.010824656164254577, 0.04998048700863049,
0.11675461096246076, 0.11123755378268373, 0.07023990538412246, 0.08086937548637664, 0.17448636501004228,
0.12074157610143528, 0.055616384687480806, 0.022041485947596716, 0.05545727982100097, 0.06183816646805073,
0.03805321054724481, 0.17169739199766387, 0.19338836802488915, 0.21689276374779898, 0.18187221679002336,
0.13171448942349676, 0.08279536719227758, 0.03508952850320038, 0.014531080252077014, 0.04588738486060421,
0.02984646660693286, 0.0729400898153013, 0.16223080347768223, 0.23909527560462307, 0.20456424453019414,
0.13935416947557666, 0.10321576092669163, 0.053358228870236196, 0.03709078842751966, 0.01061316391331042,
0.007803816830917868, 0.014770983734002251, 0.07496871264121614, 0.1416092890143262, 0.14567925667563109,
0.16753240789009677, 0.1904172931835623, 0.15403796120529806, 0.14748468570077425, 0.005029731986873947,
0.1152381430542966, 0.18227314249267781, 0.07978432624757997, 0.02773691680863575, 0.03244782738594694,
0.01747019362247509, 0.049689381990603056, 0.06568059648919457, 0.001891415130338635, 0.09738809843612836,
0.0672915683539083, 0.0018789894148498881, 0.08389042104333289, 0.16090790129349813, 0.045473234371533666,
0.05755436029260312, 0.1576164839427065, 0.35192481761902616, 0.2795054188918123, 0.2750508711477891,
0.2740863310996644, 0.1208794105538931, 0.0470734962823202, 0.023496596813397413, 0.02602783101871831,
0.01592327854138526, 0.0018223016144006327, 0.021848917117025548, 0.017992116542824996, 0.060343691105026145,
0.0668525175294129, 0.14176677488922765, 0.2030406314694541, 0.1963336741996491, 0.0005327333764098094],
[0.23025507254130687, 0.13335043045470416, 0.013706280644014369, 0.3159889392109442, 0.17310522433457556,
0.1715733365839124, 0.05201790925736735, 0.16173448890841077, 0.10276434533419715, 0.19788941949908587,
0.052849009316381625, 0.1894586263034264, 0.2685401037482496, 0.30365726598571297, 0.45731354056041346,
0.12765945233755183, 0.026326722210141238, 0.08705875385988046, 0.12442603193441923, 0.18873461530280417,
0.5859147245014711, 0.24500627448150836, 0.355025737222107, 0.35762184689767756, 0.16916470452156923,

0.0758484360293336, 0.038847607491994327, 0.14899023972991, 0.3564501628619837, 0.1415340244539032,
0.186256642446943, 0.2882168056550175, 0.42350266078521126, 0.05777174409919633, 0.0485749323178936,
0.10037941667455218, 0.046068329134208705, 0.13096743871394154, 0.32112718801649887, 0.11990167113313399,
0.10152661429640364, 0.037858890180859105, 0.20215961063238855, 0.07083047669834244, 0.1611684788909847,
0.1080865348492955, 0.011768766153371968, 0.1501721584740572, 0.007331608647336732, 0.09896881814801488,
0.20453701462031126, 0.20385739028084876, 0.014566509477484604, 0.03953394123638109, 0.08836022846048332,
0.01972285042557464, 0.079709357677297, 0.0314140014460852, 0.03573417181564836, 0.02590164726865308,
0.04365038033346325, 0.1259912057189548, 0.16443496938534163, 0.1726564047148284, 0.18179972944905815,
0.1486530573076155, 0.09440776426210495, 0.017679526157420723, 0.0015142165285113784, 0.023081325174539515,
0.06532857978794812, 0.10519350544149038, 0.1494564456600699, 0.22122124896412232, 0.14069129603752836,
0.09557221962879235, 0.06453343293154819, 0.053486103079176193, 0.05062768510131636, 0.0030895725372661323,
0.1177379518197032, 0.12068219165349728, 0.1124154875087424, 0.11541442139938483, 0.0834900018366154,
0.04481631760849432, 0.04492280893043202, 0.07787616051706166, 0.11388927677539969, 0.025898898404123516,
0.05193051382137524, 0.13357137254221185, 0.014389125417132288, 0.028959041830660903, 0.046165583031421535,
0.026866061673949376, 0.016027654169799385, 0.03204631919988243, 0.0210053822913911, 0.01847316859165957,
0.04067422308631739, 0.006595932247652611, 0.09532121079219037, 0.22546598997931927, 0.10047552648707517,
0.1251079481859518, 0.21637272950635097, 0.37653103068012544, 0.23463735281753692, 0.1977328523540234,
0.30217931767799805, 0.11665895134216733, 0.04466830001327411, 0.030252697606517998, 0.042593446879771665,
0.016722832770282806, 0.024016695191582085, 0.011630221178063147, 0.02039505665677011, 0.07117485699730083,
0.05305802142397357, 0.09155026325527642, 0.20685663409186747, 0.2696639764219935, 0.18055215128537083],
[0.26732447839702134, 0.07418039067075773, 0.08189120446937932, 0.5007083976179925, 0.3398659175721947,
0.20463466634438462, 0.01798104946613938, 0.2361423790410389, 0.04564407611828521, 0.19985498182828726,
0.07308811029604674, 0.19557969059113425, 0.26884912914741943, 0.3024030587190636, 0.1490393638054552,
0.047168886998398864, 0.1218522192048475 8, 0.07089031309321489, 0.14177322202668782, 0.24745740644634937,
0.4171003261911706, 0.2765617174819637, 0.2588808949396411, 0.3677106321452989, 0.008740696555430532,
0.06122633878605175, 0.015264148923718064, 0.09563788671677989, 0.17388843018735997, 0.17334085618730405,
0.0696477581280421, 0.2877200468200454, 0.3184931668626382, 0.19869015105595184, 0.09720793900095231,
0.20527603935241323, 0.12891822230027122, 0.08895513333169851, 0.3746687582418843, 0.3029936536581298,
0.1879693070048348, 0.13287246634654656, 0.01869814469872016, 0.057853792103288414, 0.06434610361404776,
0.004113289225999307, 0.18310410871003185, 0.14278013851934637, 0.014048487497211772, 0.057598272272428 45,
0.08959359086003317, 0.03505591080778332, 0.024085403846957603, 0.0998907370659901, 0.06078430929538215,
0.0542555187861955, 0.11358516761005508, 0.09933915210497245, 0.018601240019472984, 0.02025914544403373,
0.04893931167258662, 0.17901158664612085, 0.2240611028441437, 0.20725033384244373, 0.18812162981253588,
0.17820935317736067, 0.09595924984312704, 0.04689989882016122, 0.028306499600114304, 0.044817342243233915,
0.052281710232338977, 0.09945665856203574, 0.1802206833123501, 0.2141930335100821, 0.12012358944847962,
0.008007172996454722, 0.025233614344268008, 0.0381216757438565, 0.11111403994819274, 0.10281413184387257,
0.10033709306282074, 0.06344123006665932, 0.07917391913974993, 0.08762859712598796, 0.0633274021046313,
0.08595144392195149, 0.12035354234232583, 0.09280450301510113, 0.030166252634336855, 0.02599979364152045,
0.12335732428396574, 0.18163887454742939, 0.1083610189055646, 0.1167796568810131, 0.10097211498488294,
0.04845666534608892, 0.041652221285638356, 0.057423944627418705, 0.006767063397705756, 0.0548325804176159,
0.09274988363891218, 0.024466774 02992586, 0.005473274545119168, 0.06935482895814458, 0.11725496331158734,
0.06054054000708737, 0.10024499301368789, 0.3465933394210866, 0.27779015529676376, 0.2945888031315384,
0.3013922665457063, 0.1421941340804895, 0.0023716114462325913, 0.0769794840515632, 0.1532740455180974 2,
0.13172551249727302, 0.17006666334201262, 0.05606161625028762, 0.0362875271394476, 0.04445540696911681,
0.1007974423859599, 0.04576855181251106, 0.18287456635335048, 0.07322268691099691, 0.2438160832622534],
[0.45452077066384755, 0.14719013960566069, 0.0015760959947526167, 0.3671167553772659, 0.2866485752328458,
0.32068161336087436, 0.0815400823757121, 0.009813189278442838, 0.038456812397241275, 0.22089643540780898,
0.02238974424670949, 0.13133139603840152, 0.33199877525321797, 0.31041620070463355, 0.4213062836527293,
0.1854003506791108, 0.06394336679980506, 0.2515812212532837, 0.14683062423366683, 0.2817725056738134,
0.6893594000227713, 0.1457738880425807, 0.31128258389181196, 0.3632622991300572, 0.02493487627935845,
0.05986644915859907, 0.057093358179239044, 0.0737027885745956, 0.13086186954999798, 0.04400237441800009,
0.09900479553212255, 0.09781514316560223, 0.3543516798338008, 0.08437135828054379, 0.0771050244638152,
0.28348715053729295, 0.22639955185613364, 0.09220982521682156, 0.2697276545890229, 0.11947514218917492,
0.41735957507801147, 0.31223233617005114, 0.03271305118091005, 0.015724387534234736, 0.11095095001305824,
0.08925104129330622, 0.05150865599063211, 0.21302362410178513, 0.036554714523429635, 0.06089816062838515,
0.03437884414290283, 0.003200959442463786, 0.012269241621715653, 0.042287936803016395, 0.024951367814525987,
0.0493850982010135, 0.0063097032739153356, 0.113412782459822, 0.0955578577084215, 0.1068016111981131 1,
0.07376405257829685, 0.0328607974587472, 0.1059128939068946 2, 0.15260601555456088, 0.16392240143904685,
0.14918741883332462, 0.12448227219885874, 0.14404537863644365, 0.1707546443367515, 0.18816638290356186,
0.1887007225968233, 0.08434112919130145, 0.0777610323710123, 0.19464952682047384, 0.19621750057019008,
0.11911887279816415, 0.04271469571856012, 0.10379821297921335, 0.2034120326604417, 0.061000759069283275,
0.08495142091358868, 0.03484233445241046, 0.05520052440606389, 0.12800858391006625, 0.12234050573572408,
0.10001333927180832, 0.13582623558044138, 0.04012443281368256, 0.0671273198941298, 0.24785258762154522,
0.25872714984235523, 0.3422225123258042, 0.2070665999391074, 0.1356394999247167, 0.05011262735572305,
0.13649434218436718, 0.18090805181805106, 0.15092066883281038, 0.07780781364024035, 0.16164700623501568,
0.14089087133300338, 0.23399246400558393, 0.1526651185359717, 0.012332788743547145, 0.025514145059298846,
0.060581369515523675, 0.0568999117365936, 0.15899841570820786, 0.12781378170347482, 0.16958378433492902,
0.2004425726573516, 0.11943163698654953, 0.013116206591420614, 0.074493385824896, 0.11747206529089543,
0.09225439888093499, 0.08313918674955197, 0.04009876725631639, 0.009176702459605151, 0.03589781868988575,
0.03828143296235521, 0.14016929932148325, 0.06937068135149405, 0.09947529430830745, 0.19881893802748316],
[0.382917140739578, 0.380706964400082, 0.0565701280494962, 0.10264268499949707, 0.14063611313585314,
0.19438078500154457, 0.1982191903264528, 0.27737335728822593, 0.08312930459099124, 0.1115766865655921,
0.3656433674131905, 0.08272988972826353, 0.14029212752484077, 0.10612607540478095, 0.2095896580474221,
0.2831077607896345, 0.09182744320603317, 0.11586282802084852, 0.10821617764969974, 0.2693043289744271,
0.3151718887021705, 0.22874312237328193, 0.21410195946394756, 0.6017182311344896, 0.18855354270066763,
0.020825372053534963, 0.06425471567010753, 0.02911607901072149, 0.03004731579351255, 0.0819189656921218,
0.06341170575583542, 0.32258772860571816, 0.23645236018281127, 0.12338868498603632, 0.016279706746888622,
0.007249543212336582, 0.1297106708436983, 0.045375906110232546, 0.20997364604519353, 0.2580418951928025,
0.2919500229443455, 0.2232156100037349, 0.012414757112811101, 0.019547456007224278, 0.11573871444418883,
0.07197503723427177, 0.13325745053978205, 0.12503050169271468, 0.00796874646449312, 0.015421895605831081,
0.02400681225731813, 0.03960618822915912, 0.03994222677851054, 0.06458502952860297, 0.10842091661211231,
0.08543044229198951, 0.022280537230610395, 0.045071145539779894, 0.021677093632779417, 0.02271245584002799,
0.01476813401976740 3, 0.06190922868836803, 0.11043051555174407, 0.10338092865439709, 0.12339997988414442,

0.13288340726711, 0.05251755549975859, 0.012881269807607303, 0.04253100644942976, 0.08583907892416544,
0.051441255371963396, 0.06227725878484277, 0.16132873834672892, 0.23914993020860512, 0.23139374445466132,
0.18327747068747874, 0.1747314843546431, 0.09039841979176429, 0.052217693026810626, 0.09786550278598354,
0.03665963214706868, 0.03555892157930899, 0.12862291827756767, 0.20249708350203444, 0.18781412313825452,
0.17271639788777438, 0.16792116666406806, 0.12646131471847674, 0.01903192458362241, 0.1389626796271652,
0.23014862215511242, 0.25781373545074404, 0.14660427589868583, 0.07269844508846719, 0.0548624598079999,
0.11729412017056677, 0.2378110483017106, 0.17285474676317386, 0.06566678530484263, 0.12499793861688108,
0.11534718106190038, 0.09240771158100466, 0.012370515476867057, 0.03188559099595591, 0.040561478200323305,
0.1419500737487691, 0.12644696621356588, 0.35753620715939377, 0.3027297157979105, 0.27699009517678974,
0.2267659020292363, 0.14805915849765058, 0.019585916570552753, 0.10317781520003654, 0.11995432869456035,
0.11347448093221184, 0.16442780921858677, 0.03833096183451551, 0.00034753052964926236, 0.013255127058375032,
0.008118305787102084, 0.018440980807675294, 0.04465684291448973, 0.1016232886286502, 0.17813058670230214],
[0.30129988513587924, 0.3233356268776296, 0.0944752119858855, 0.1155248953620151, 0.09854541876816117,
0.19705428592964247, 0.15123943916700092, 0.07040857092660502, 0.036423951046024, 0.2560240683028603,
0.1039300225699997, 0.19101046945201003, 0.2093501867087223, 0.26705352334047033, 0.34783891572607856,
3.8454501951575216e-05, 0.18731629434522548, 0.05663677766839029, 0.10698990967836684, 0.005743330721199486,
0.44174403310010507, 0.19494809961861162, 0.3685132262604138, 0.287243188514657, 0.0980580278722987,
0.09132380709082488, 0.06909293219516967, 0.024966605083026772, 0.2230644704990507, 0.05511617228097389,
0.058398094294338114, 0.2288996355782585, 0.20193274628490163, 0.07116203850497042, 0.03940600715625561,
0.0481358563906435, 0.01255936912640934, 0.012365384033098245, 0.2850300058743004, 0.17162906122452412,
0.19265450302131704, 0.1820620881679206, 0.04146602530079773, 0.001382169387501532, 0.011843077726332717,
0.04876435844257234, 0.06874778288653045, 0.1350991866129552, 0.05091954007686001, 0.08614316482937354,
0.05562415976690002, 0.04808842068450191, 0.05422484235269586, 0.05733169676794809, 0.1079684667759427,
0.07891687721011081, 0.033305761431597664, 0.04127822697643844, 0.05092153232371666, 0.006958689041212372,
0.02269565312167836, 0.09824035704914738, 0.1097245678180748, 0.10438113742371574, 0.0912176348536772,
0.09616936791702949, 0.03429929343480624, 0.01421411552961296, 0.010890821776280302, 0.04400455578431531,
0.04332095097878192, 0.08483168651897945, 0.18332047758160086, 0.2705712248379661, 0.20061143206132037,
0.09663878520282984, 0.07157462436450153, 0.07322251567438567, 0.08674856654796759, 0.057277113201808616,
0.03150769246512703, 0.012154595621195913, 0.054691082726099814, 0.0657870093157731, 0.09573819967279774,
0.14708641338755635, 0.11947335880071658, 0.06871532618180269, 0.010398618710470003, 0.13232022697889745,
0.17103696680415892, 0.24323781575328687, 0.07369110590609441, 0.04183613736125214, 0.059119919092510784,
0.05628988118523047, 0.10011016431787526, 0.052021953002691905, 0.00752526166244409, 0.0654604880334273,
0.06552111177435281, 0.03413267701216536, 0.05237881697161766, 0.009861966666809512, 0.0013653283575878752,
0.1663699987322263, 0.15271545904341136, 0.34020165431978583, 0.24224282483003184, 0.20727642755178916,
0.2133965001569891, 0.11694596683692202, 0.004255333020110565, 0.04186368791700202, 0.048771475410079576,
0.025159505378315787, 0.020222386484259107, 0.08428149904182138, 0.05550882308373057, 0.07751986613726387,
0.0010070489444754492, 0.09535459337789029, 0.16536871240050424, 0.24763449607568333, 0.17906279531589772],
[0.28387474747758973, 0.40261255467666096, 0.03818431320287694, 0.32168947239166035, 0.006729462479554468,
0.3723921801225567, 0.02284671645856422, 0.07294215691449062, 0.19390416789059664, 0.035939658094197136,
0.13378828300605577, 0.17431301272158223, 0.30367091997740897, 0.24810114076907544, 0.21474071273522075,
0.12475547527122438, 0.06367938450757595, 0.051105108276652225, 0.10184610972265484, 0.14590687317907072,
0.3702360267204561, 0.2536739083166755, 0.3228800722186817, 0.31604643668118093, 0.04571047935146659,
0.048454850181155376, 0.028894363803568107, 0.023763991693913255, 0.25201203735551203, 0.0045267132636773785,
0.03348540398102352, 0.13422353538278037, 0.36671336320392944, 0.04670362064630371, 0.09430975681995947,
0.20866620561998087, 0.1716181566872061, 0.031117158852758012, 0.347851199978029, 0.12856855161756917,
0.13528534010030002, 0.1367417129749188, 0.02144780953032528, 0.06842807259709786, 0.027656470561592185,
0.047419473143797436, 0.050845941894739934, 0.13672811451457703, 0.06155991940199035, 0.06713131273144393,
0.004477270134255678, 0.09759071378648732, 0.056621086884811475, 0.0017517187868724485, 0.023882796811691257,
0.013983835376054535, 0.012174503436772432, 0.061312723405043935, 0.006851992540189486, 0.021536549617608036,
0.009824142998667742, 0.08363752127594729, 0.16755861347888218, 0.16321329283686797, 0.1575815781421649,
0.13799799626101103, 0.060434449508932696, 0.004903373995600057, 0.014889845988130628, 0.03887548092568709,
0.04500786136876486, 0.05434593622057475, 0.06391964984555291, 0.08507062016167262, 0.07790903165836832,
0.06666673911128601, 0.025198716725011283, 0.03959886092478474, 0.07108351200336326, 0.023311801697444037,
0.025969470541986228, 0.047606048768214795, 0.12475920808535053, 0.1755228541611667, 0.18920387180718767,
0.16417382768264324, 0.19845204809246372, 0.1627296543382429, 0.08776347805350762, 0.11089739032967243,
0.18224582070622533, 0.24336856901576207, 0.11768428718168551, 0.07828600618116066, 0.07845250232784533,
0.08095892996857632, 0.001685937255928812, 0.032607016797889056, 0.14837040274316995, 0.03140559306678374,
0.11167929823331163, 0.09001907907333717, 0.035956045553851596, 0.04138054498247461, 0.0861450212089892,
0.03213541501665058, 0.056374305193472515, 0.27665641871977126, 0.24788271939711853, 0.2961160643175985,
0.32599652884916996, 0.2000579756183693, 0.009833066627085527, 0.015418868900936125, 0.09950770154044887,
0.09492807758418134, 0.10164889619804886, 0.025624349984193204, 0.03759861649265547, 0.011217113940612042,
0.11826212366368129, 0.11034960105690186, 0.10822606354032142, 0.1775045309085333, 0.13518441049124552],
[0.5543345107216953, 0.012651604851071335, 0.042653917557432884, 0.1421546646972727, 0.24177086064261713,
0.162042086120326, 0.2749447977533534, 0.29082454524866697, 0.2010318681313839, 0.05246834008877389,
0.004698775483188052, 0.05401541843084603, 0.25002576666547105, 0.2101359014258696, 0.260089265096817,
0.07971742741162328, 0.18216154476940166, 0.16599056784872904, 0.041958655265115476, 0.02920244426700659,
0.3887063143746796, 0.31410356930839334, 0.3379994911923742, 0.2759508072584957, 0.04214235723357028,
0.12751941825085192, 0.0869249992793832, 0.07821457084812533, 0.2755992225552068, 0.07467703887129928,
0.011071107114839196, 0.2000394274387526, 0.2667666979978693, 0.13787764610825956, 0.06124477084048938,
0.24443722721897407, 0.1259395316207223, 0.029095397538868795, 0.19152250172738194, 0.06952353198982657,
0.24392017681119593, 0.25897503876520495, 0.008238220822652334, 0.007544973771251776, 0.11862493175867349,
0.1120343023489858, 0.029628230363721985, 0.14637829977554745, 0.005927151204023945, 0.007417661412193907,
0.04241550702270087, 0.12376165193533935, 0.08561644514593254, 0.03892423009842472, 0.07647577331369522,
0.08137135341408706, 0.050670688413004115, 0.0004988491253925509, 0.007864588222769992, 0.04455665433492921,
0.01494209175776919, 0.10911901798285048, 0.19380932081812333, 0.22231213854859144, 0.1869032314762941,
0.1338504164361805, 0.06602076542805581, 0.017476983166898073, 0.009009004045025455, 0.04587139064208562,
0.019659677002900326, 0.05763396486124132, 0.16385969574363785, 0.22585186472077956, 0.16579952441747062,
0.069672781491986, 0.028948496092092966, 0.1146404166176540 8, 0.1595127597819975, 0.07278998453161072,
0.011657194675492881, 0.009445691671650672, 0.07841910267105046, 0.1396198585216961 7, 0.1700405274754768,
0.22576681180147828, 0.22451602798312328, 0.14298461664798853, 0.06083707053064989, 0.14770689538569717,
0.23869173575698327, 0.31833753351437355, 0.1505723013847271, 0.12921159196753926, 0.09293125557993241,
0.06773993112611178, 0.016416727775699334, 7.017379365842545e-05, 0.08022726563683587, 0.06451499830429411,
0.10310710807631163, 0.13310837600696226, 0.12727240667547982, 0.0460908728330566, 0.03754695826356422,

0.1544472192492834, 0.14445669736257977, 0.37623152765440926, 0.29532204963865005, 0.22028806054071867,
0.2429817719297232, 0.111520467214197, 0.012291272282269708, 0.05873235481506512, 0.05014808622716472,
0.044884845781276564, 0.0010422295687153957, 0.0397627629746658, 0.013623972215866496, 0.03738566669074429,
0.016003029927357207, 0.10558372651888953, 0.16811701795894085, 0.2351781522705019, 0.19200780941842382],
[0.4208822690302852, 0.09503554699295995, 0.020412660420498396, 0.21409476019243057, 0.32289316890330283,
0.24769450205009072, 0.26550216751357714, 0.35300987226602076, 0.1035370631152868, 0.06594548909058212,
0.29762200324907334, 0.19035709917977153, 0.2237854282153682, 0.2772272087204675, 0.3451981685815388,
0.04244345463057185, 0.03426864261842122, 0.28953679406892335, 0.19096281132383924, 0.11823329503541014,
0.2510516287370885, 0.4685552278184187, 0.237608654522739, 0.30486198861896746, 0.19941143194125596,
0.06519242623761257, 0.05951687258683863, 0.025020941228088234, 0.13263967205139607, 0.029755649959486978,
0.06081783930364262, 0.054738150113594446, 0.15133629060518652, 0.11808365413058958, 0.12282377301425043,
0.22636837423465853, 0.23524193512994254, 0.026408739895891226, 0.08553805156202285, 0.011606590821943184,
0.08375562939770517, 0.20565725931844295, 0.011365306144926406, 0.016945850629437414, 0.032245797358060435,
0.13830051307807437, 0.0760274963240198, 0.09002339281436887, 0.003211696069886234, 0.00902086432381525,
0.06430467928578334, 0.1648153022917085, 0.12842309698275012, 0.10962909805235642, 0.10126810552678141,
0.07787805602561045, 0.03709275086166093, 0.01633712225569517, 0.04972528576156941, 0.014298507037426302,
0.010318823317450614, 0.12242121475192119, 0.2141021646900665, 0.2101207938185885, 0.13924710570151422,
0.14340429524893533, 0.08443470315347046, 0.03726788189823389, 0.01007737676127291, 0.1061326770728772,
0.09883344735950003, 0.1384142454080234, 0.1941051004193859, 0.22652872715237685, 0.15029328331800562,
0.03997143186850265, 0.013685815965643122, 0.06428070202467571, 0.08255053826055013, 0.03601936727136795,
0.0927140981501728, 0.12286406779048148, 0.2161585436971634, 0.28944422279226856, 0.2563061304074931,
0.22243664208902258, 0.22969319515360617, 0.19524107751138642, 0.10350772684142086, 0.012229011475145279,
0.1287771020444432, 0.23653980425571483, 0.10415129653438224, 0.09673739122412198, 0.11838176014578664,
0.09469956653915715, 0.024748691821988363, 0.009788056875489877, 0.05098357464619203, 0.04814602192905597,
0.10871464335870315, 0.08450314608080103, 0.059484668760710006, 0.012960347422673275, 0.09376149978931819,
0.15353575951050857, 0.16218227629081908, 0.36632643137656673, 0.3081869930407309, 0.30779166448872913,
0.42935890634124685, 0.26455880670939264, 0.17641544706410592, 0.05747300558391989, 0.020723713771581467,
0.07412214755333409, 0.11307822463110287, 0.06524672108293897, 0.03634599509991988, 0.03246851741346066,
0.03011949479474137, 0.047575977143539, 0.11080241539168915, 0.17463279911811166, 0.22792174096232157],
[0.385349307913296, 0.13139741530018706, 0.0711340322495046, 0.25247380841672984, 0.16126312946901175,
0.3744173829110931, 0.00410823361602298, 0.21792995252630382, 0.07907271842027047, 0.1989963831294718,
0.2986232891682488, 0.3038961948775439, 0.18443656429470948, 0.19409248812874255, 0.5094392113314004,
0.0013841378253468726, 0.0746357752746466, 0.03377059140628568, 0.1752011061831405, 0.05912862432100692,
0.46799928378899386, 0.3209099026555611, 0.27993766113270835, 0.23151914249853822, 0.037210916790872205,
0.00490755636092995, 0.006913374038067523, 0.09734068146194082, 0.20769913199579745, 0.0030449178596142846,
0.016352870434642192, 0.17184587336054827, 0.1684895559305649, 0.07377885278697166, 0.007722298733667239,
0.21364363144482124, 0.11696189613725197, 0.03952543830805079, 0.1606237761939664, 0.15428753124370348,
0.2585118930876613, 0.155905899837972, 0.07327476078172772, 0.12191096845887882, 0.10305648067335715,
0.0121006468658045, 0.007672296466948733, 0.21583812038953143, 0.02887726641718835, 0.014964645030577397,
0.09497304385010942, 0.126603692840368, 0.07273201005208355, 0.06987229563512352, 0.09827743504739408,
0.022769738506727116, 0.003519792068261698, 0.025563584036703467, 0.03907068228074973, 0.026028263101996022,
0.07077337227504855, 0.10004344483347752, 0.15825656446152855, 0.1494006748702729, 0.10325868925101334,
0.10276012386657438, 0.04837578744798382, 0.013145982806774354, 0.01579780159054122, 0.030147047459473886,
0.0426365510054172, 0.09571733582085765, 0.17493388367040066, 0.22731831206515432, 0.1974826745494683,
0.1322807984615176, 0.04198740380786278, 0.0376972593649182, 0.07787771960264475, 0.11688973098003437,
0.0796609387879808, 0.11503711590691515, 0.13074000076744344, 0.14342562761381136, 0.13272052089607997,
0.10620276197346887, 0.12258967450196022, 0.09251862482080504, 0.013278482520438024, 0.09664611167693019,
0.08283356526860673, 0.16947387878181924, 0.007405135607528897, 0.04214857259114861, 0.06152959752953765,
0.05615204518753711, 0.05561210749197996, 0.027331893475538053, 0.09489729889469649, 0.01691393719378633,
0.019502267725082508, 0.08862085219708034, 0.10468933377377199, 0.04500065001354674, 0.0871899339840034,
0.20089860090073608, 0.1731324745364067, 0.333229534161037543, 0.2030923355864987, 0.1698750291205772,
0.22672550265645308, 0.11135708789232596, 0.04156163966915359, 0.05764420449440237, 0.0524142617915555,
0.044080224459290475, 0.01454320197185664, 0.04593985263657578, 0.06146792392089602, 0.11736137421915835,
0.031104090371463423, 0.05549259840666258, 0.1246892473811292, 0.20595188030149508, 0.1219522540370498],
[0.2681970160620495, 0.13542755733265827, 0.019277521284486055, 0.2921126875771155, 0.16985050880397018,
0.03289664860173277, 0.0977042091560446, 0.12586162699630815, 0.14328016314440173, 0.11233468857653793,
0.0013260372429472296, 0.007705396624953142, 0.21375516654657092, 0.26413078493105435, 0.2965749904664439,
0.11803463243222513, 0.19770190394879925, 0.06438937872385611, 0.134338139659734, 0.19441990003170276,
0.40970771418863505, 0.4170056753802919, 0.22805770880685758, 0.4985367839720781, 0.15509433881472615,
0.017488605658638925, 0.020374371267026754, 0.12228377371201213, 0.16142625795989246, 0.08064074835256202,
0.0740180178831735, 0.1485537807581242, 0.259256527426122, 0.08085000396608262, 0.14891832701427543,
0.062443896364256964, 0.030053280843685175, 0.022473294810343557, 0.20924307883524218, 0.07063138784945201,
0.08917686227381508, 0.12388757826278779, 0.07323403269716383, 0.13070710619041515, 0.05132492983079737,
0.0797974824429061, 0.13943129334325383, 0.14189093373440104, 0.035666682139592905, 0.06781972820352603,
0.05744836316172551, 0.06478065852050516, 0.04465595808338489, 0.05689090324796147, 0.09297407665439993,
0.07491199391692284, 0.03827583433678286, 0.03335906895924104, 0.05725651658924608, 0.01555287446708361,
0.005291543771265456, 0.1020825713709587, 0.13233606629430053, 0.14536202569527099, 0.1187940911023736,
0.10830754425027, 0.04390280287687843, 0.009988250897166519, 0.0365998720029844, 0.09189655688490755,
0.04486551711926739, 0.008976959723864156, 0.026472042589228032, 0.02362793807899423, 0.009209148080851232,
0.04659636168867074, 0.1334536486752775, 0.2106936933166977, 0.22204239484627086, 0.12420248341940665,
0.0865009552927129, 0.1300889359099413, 0.1835267399548474, 0.2179027670251155, 0.184491723756284,
0.17406865801582075, 0.1432919695210926, 0.07211548309959126, 0.06280469544759872, 0.1929022557046901,
0.25233582273366584, 0.3502914365598921, 0.1986917278034894, 0.12490933270504796, 0.014498212083424859,
0.009606655044272853, 0.09047063559516512, 0.08536996150438704, 0.02283355498511082, 0.040546995388788226,
0.045436755717020674, 0.06276011498559536, 0.06764096132693838, 0.04114976667562174, 0.011319942240178306,
0.12609530106479577, 0.134030923336935, 0.2787603375241129, 0.20371325770567547, 0.13896686779029255,
0.167582956559449, 0.051429458140811055, 0.036798319481563375, 0.06446047383028397, 0.03851554207318386,
0.05821119331091461, 0.0766118747365497, 0.014575007509943153, 0.013860782151699553, 0.056124457274446676,
0.04917553108864721, 0.05110736953407471, 0.04082722225220689, 0.15690906882783456, 0.09373811351224046],
[0.2649493365867719, 0.28885000915057135, 0.02281436572196436, 0.36887616272447304, 0.00369193665840229,
0.18574088013423393, 0.2669668954807199, 0.2511750625137204, 0.0040963406512801795, 0.08534387256973602,
0.35093980389432317, 0.2523949966909811, 0.37997371695683, 0.21598675923644245, 0.42972884080774704,
0.012290141198879192, 0.007167159685642707, 0.09897506822826539, 0.18109577032062496, 0.1340526293660412,

0.5167781599956885, 0.2634727594284373, 0.19544702086487076, 0.4256581217371381, 0.29714625223380725,
0.06659790876491575, 0.0067980836351544, 0.12208746699381398, 0.1545222929652763, 0.014409861941361164,
0.11977167343661344, 0.3208710043509156, 0.2476831303256597, 0.10870571740345181, 0.017209691845772805,
0.14682127757624233, 0.021418212960643074, 0.16243613682088445, 0.40378326983246443, 0.38429622800921115,
0.3012074288283891, 0.1669557015707661, 0.033616450981940535, 0.11432190727303439, 0.1738173228043319,
0.1206093093346555, 0.0004975366622698912, 0.1122772918982967, 0.041702304708963064, 0.07903849128842944,
0.17564810909696416, 0.1872883354220166, 0.09043089002435993, 0.07442483317219606, 0.11302780760973893,
0.023415435172237917, 0.04849845924106108, 0.026550316072825424, 0.03251657748964634, 0.053520100180024184,
0.026415008957219308, 0.029769669251283153, 0.09664825788410485, 0.11433885283993336, 0.1420209812020047,
0.1284211642598684, 0.06181142204742312, 0.03919294969173938, 0.02730573431864082, 0.008530604135538678,
0.04304843637602438, 0.0034566932497548087, 0.1142958546374156, 0.27189594330702016, 0.29913347462152073,
0.2131382879466346, 0.1590168508403094, 0.13325691237659584, 0.10283137728437769, 0.09128295487031092,
0.11766165522491982, 0.12410250134774109, 0.15464287834233778, 0.19564569970328344, 0.1456022843397463,
0.15223830421302453, 0.14254991399508202, 0.10204780035813292, 0.016611149362414073, 0.17688683849402131,
0.24737755865327538, 0.3295102633483657, 0.1421155490387896, 0.10263285480317057, 0.014343637095605648,
0.08822356416842154, 0.21494627514187695, 0.16397712066217449, 0.11801342641149601, 0.16790669456854643,
0.16732858729406205, 0.16070730389706936, 0.10494143615708851, 0.03334215681809515, 0.01295913593475384,
0.050216026838020546, 0.07666235497381638, 0.2888162670726405, 0.2516666339634429, 0.24670324547929914,
0.2822440981528421, 0.15804924556695107, 0.030684908457062572, 0.04405659659166824, 0.09780685106193314,
0.06160181773409289, 0.0723344595269604, 0.016970523627655387, 0.04278574439908689, 0.13438289479089408,
0.1852507038265923, 0.012635078447193073, 0.054181707877177014, 0.23130816325284526, 0.29972950845999531],
[0.10605094904737561, 0.09135113106977859, 0.2079925489380134, 0.24609492888734563, 0.06966070684817463,
0.18586908718312428, 0.025116098536411606, 0.09229249049950823, 0.24889915813211966, 0.2198910899445158,
0.1235251557508169, 0.10770402469074512, 0.2721202182221437, 0.4381148747816473, 0.26451463243390644,
0.007479809521427135, 0.0404544622414285, 0.32809733508147926, 0.07764632582555761, 0.21590777942119088,
0.4674875845942948, 0.3512994171160011, 0.27170235685685346, 0.2888967752209756, 0.04999455351652352,
0.10502062123963178, 0.06258634400781768, 0.02084693798334076, 0.11339502181810139, 0.003565905158918541,
0.0026150014471080325, 0.12000620822708183, 0.1817182606566962, 0.1661292945724735, 0.059135718027600384,
0.20642489242335646, 0.23702629509238374, 0.0372307392811315, 0.23127542638287601, 0.05817891823079408,
0.17833167440238246, 0.117232059810942, 0.03252888570121621, 0.01154437012134293, 0.054801574771225825,
0.0716251711142857, 0.06292912291147945, 0.08475992249321816, 0.012585370116992779, 0.0005191052640340308,
0.09066714503448603, 0.12277157942832041, 0.055476111612775485, 0.045896478637041944, 0.0529669169026428,
0.057910760173637596, 0.0119202608729723, 0.03001426133880461, 0.06434946940110484, 0.008833195107104315,
0.06989901675922731, 0.13022277237733185, 0.15095357305262339, 0.1580699204093008, 0.14350725863185085,
0.13019403470963226, 0.0756374634755116, 0.0485050640120074, 0.028423149413910463, 0.012322740843514472,
0.0027301679010801336, 0.04635912771853695, 0.06725086114982169, 0.07132812828675425, 0.05685882638376847,
0.028338623286056266, 0.03466926971793245, 0.10029167780477032, 0.16827337896129266, 0.15141385557935771,
0.18857961972081755, 0.18847020072941578, 0.22507811348523546, 0.22625382142147624, 0.19443129209122678,
0.1309212848069399, 0.1323162515131503, 0.11861815892209436, 0.0649768676061289, 0.05003374158494159,
0.08737054297049146, 0.1189771782373045, 0.016338918834451353, 0.04327527170216189, 0.08180804349818441,
0.03496540568219219, 0.03335351585785283, 0.05702023291717294, 0.018622363150102718, 0.0380346792477203,
0.04772927380201511, 0.038611157834739555, 0.0031206595585903116, 0.04320639292609077, 0.02508565027580622,
0.00682469681363603, 0.053841961390671954, 0.25209982559599475, 0.19917485995652096, 0.26788904003697017,
0.3459831767164292, 0.268042648099096, 0.12216866368434019, 0.024975050956193765, 0.033705672919568795,
0.03711383070891763, 0.03317402832222692, 0.1005724585935942, 0.08320632205430535, 0.1320048910095467,
0.0974791147577609, 0.003101465864837199, 0.06913798510842119, 0.23104246529061087, 0.2376044188460186],
[0.3802095228520747, 0.18217052015399188, 0.06428408802933165, 0.22261387010517844, 0.25452335525071124,
0.14147371195687075, 0.06707286087376334, 0.0510722641810304, 0.09486468373573198, 0.38305628454547785,
0.07287906752617573, 0.14835143592365807, 0.3312256209654469, 0.32385548135661624, 0.32859989528858097,
0.1864098306811858, 0.11858056285901604, 0.10088987357988495, 0.11519072713261574, 0.16495135901411634,
0.6937777248980921, 0.2341661131568464, 0.43870933301140486, 0.3240645557501761, 0.06164863896919192,
0.02566376864429886, 0.035770181808413716, 0.14407257719899583, 0.17390615331765355, 0.011278501729450548,
0.15322277045168978, 0.1500069352497442, 0.29574055601662347, 0.12231123777197003, 0.05673643915681951,
0.14605351267148264, 0.05161959758139906, 0.1331331256272483, 0.13761081882392365, 0.020251290268261942,
0.08177789162702478, 0.13312416467795268, 0.02068688760361933, 0.09676571027473618, 0.08801040280208469,
0.18519015189213595, 0.16665187869064813, 0.2258035994909301, 0.04165681019142014, 0.004844231731554343,
0.020593961638460317, 0.037550287158755795, 0.02026578078380285, 0.03264743662769527, 0.01923717403580044,
0.005359894187917567, 0.024975626572125563, 0.055939107731032764, 0.12205572405710544, 0.09056241348653803,
0.09257062232586016, 0.1341047318090781, 0.18178991739282496, 0.20028651229764236, 0.17352254335853284,
0.16150758749874747, 0.09246053201199855, 0.005504972725423663, 0.04960942209128695, 0.10898816600047681,
0.11713367403318585, 0.1169982124279984, 0.13271660316090705, 0.14347411371925545, 0.10401520083606607,
0.05929983302303092, 0.1034289494044146, 0.11943172910392795, 0.09848312854260413, 0.057612337906240346,
0.06848649824396146, 0.0835946534301279, 0.09137454289796842, 0.1291863573470829, 0.09211250363271835,
0.07447444669587046, 0.08054032191913921, 0.07062180533119011, 0.053247847787469035, 0.01306610525007465,
0.06949122957189185, 0.12089596169466789, 0.055964316381285746, 0.019579741788952575, 0.04549810876343275,
0.03060840090574901, 0.04953996542328853, 0.055538419465615316, 0.00904792287863696, 0.0829144473418257,
0.08931225999443299, 0.060814036989582446, 0.03985667051042217, 0.024331089570908963, 0.06576182558785551,
0.11299618290120723, 0.07978638455219113, 0.291228194140447, 0.2200771205452945, 0.29322562687143383,
0.2737276363983933, 0.1682165963644857, 0.07186307663821212, 0.04202590446117728, 0.12122599247837092,
0.0338156020545874, 0.10308601445065808, 0.008844233688078205, 0.016082298817580165, 0.05042439961201059,
0.07443355737698422, 0.01694335408476065, 0.06338199959876138, 0.18884090524317917, 0.16800895714533493],
[0.4180237693121848, 0.004680351211626526, 0.12063132421027882, 0.12263418723777841, 0.15958650878578962,
0.1403777961263605, 0.15861129966425314, 0.011897628512945915, 0.007896655664265109, 0.06429748132653171,
0.26469174978868526, 0.2829965131285655, 0.3583539260122825, 0.3317243113836612, 0.3479507191056702,
0.021702399101798973, 0.012316001476032751, 0.24927733432661525, 0.12536582761038687, 0.22143483698277935,
0.5241446652287127, 0.2370864647518788, 0.09615557970513021, 0.4838880975212757, 0.09306084138123816,
0.043449292604873076, 0.04275238759293028, 0.09023989147022339, 0.2769690683251394, 0.08384305085300417,
0.12318829181694624, 0.15018746783877673, 0.21410761321160707, 0.18234000090059566, 0.06032966591259094,
0.29233673546534783, 0.08946526492492543, 0.060989778405105916, 0.31139222895387997, 0.14581227761992027,
0.20012846884253352, 0.05716541943427437, 0.13086891552911165, 0.0891678729550795, 0.1372580901891207,
0.02800451545542391, 0.11158645591543447, 0.25093673103299313, 0.06844343057130264, 0.1044144378098933,
0.15461442667970213, 0.129090911274685, 0.058308910102873944, 0.001953256353784311, 0.06587394579775299,
0.03594820677108922, 0.017178302540667417, 0.0012841770204153591, 0.042685780049116016, 0.015206244923222508,

0.03525628592044633, 0.11955935983389378, 0.20658218822106214, 0.2086928753070356, 0.17655489899097365,
0.16899489348649196, 0.08488615087445617, 0.00978529777612072, 0.05867859243637078, 0.15446272676743322,
0.16220577383713627, 0.15133006214694858, 0.11490923597866522, 0.12666553224679206, 0.14651297177199596,
0.13750712765539863, 0.15501483700581437, 0.14071913780731482, 0.1006850528426117, 0.04386434645818882,
0.046946697936584895, 0.14192191170616558, 0.18856732737864854, 0.20633628974535856, 0.15862627093677337,
0.1774799033903648, 0.18571431217377773, 0.1436254369215383, 0.10783667713076633, 0.05634513495022897,
0.129303153673526, 0.23983089915981304, 0.13417117911645682, 0.10645982327657301, 0.025110445569358826,
0.045624655733981, 0.11133133764572159, 0.10301728534131828, 0.05592030994457463, 0.12362987594289607,
0.0819915319559661, 0.07022463363167657, 0.008370308242872538, 0.1115078871687731, 0.09688560297605353,
0.046780468949441094, 0.10382271029681395, 0.29976181930326734, 0.25915426385685925, 0.35100248831152464,
0.38805212135640244, 0.27029098703058146, 0.10231799762929229, 0.019395248976347864, 0.08747690857854913,
0.02074125778678722, 0.07310330050588673, 0.019505529839398985, 0.03469466290818553, 0.04871454200635474,
0.04244639168941052, 0.14354088032914292, 0.137165801635411, 0.3002761330359137, 0.171527837241769],
[0.10939705721974953, 0.2605483242876462, 0.17559519780029878, 0.3523925381194384, 0.15509433931998814,
0.07027279377317534, 0.17248228668662632, 0.007303541541625069, 0.008486271055216812, 0.08344269396432165,
0.06364532627320851, 0.28109823255551236, 0.4603175409415288, 0.3269532985575703, 0.42910169095889394,
0.20175359709518592, 0.0219485130008318, 0.14592912739136185, 0.16269655907984612, 0.03805146189527301,
0.7174230464563638, 0.2904800616602023, 0.42027563725700734, 0.5021735556397524, 0.008131249034956916,
0.002006013988557407, 0.06011217939278258, 0.1335862049441239, 0.22602474192034663, 0.008169234208811006,
0.032970318104830594, 0.19733931743512775, 0.2935478609478905, 0.09458366363705022, 0.031308546244961856,
0.1897830377729514, 0.09457349296220625, 0.20708094597277574, 0.23631186193792533, 0.05583272666384194,
0.07173414745431242, 0.04053299406539703, 0.06504647034381142, 0.0327110295261261, 0.04598918929801261,
0.08805829251294457, 0.15530188747966936, 0.21149194378211825, 0.03670881897081746, 0.02619769338354059,
0.08666779566942723, 0.10075699319942145, 0.01656677348756847, 0.030220209539918916, 0.1279727613655774,
0.0727914733217425, 0.03374932109962438, 0.03791046827184641, 0.11501694990223754, 0.06569896382267117,
0.141561746638187, 0.18123142081816362, 0.22191188938705506, 0.20519265242878418, 0.15806344602880237,
0.11984907679476813, 0.0653227727087835, 0.003955440292261378, 0.06385621779919998, 0.1308501502198042,
0.13357594250111857, 0.14103265889990438, 0.11228865927788936, 0.09318986154410003, 0.07123805445530974,
0.07306838342580856, 0.059648386495947725, 0.010880905295908055, 0.046478544362691665, 0.061957437054889875,
0.023201087959750396, 0.1293080422730989, 0.23005011903507377, 0.2752002757186224, 0.2233326752067068,
0.2062351767999879, 0.20739432118238435, 0.18493136466492865, 0.09538060778207696, 0.019792482994214777,
0.04108013496779745, 0.1355865027978927, 0.034756197214290824, 0.04104429796074161, 0.019749239374006793,
0.04794829177501262, 0.057034364325580136, 0.019446426000640782, 0.0022333775688861213, 0.0068021806566089076,
0.002761633030960374, 0.013951205803678599, 0.04550606123523677, 0.08753348196168867, 0.029291490478063276,
0.05040559171265712, 0.10716998032603531, 0.29546044477555217, 0.2567668423214544, 0.29830733191948683,
0.32291417244089665, 0.19814911130489896, 0.11086504213103844, 0.04028726930447341, 0.017823389150685504,
0.06320609191673542, 0.0185042951443731, 0.07191609720741415, 0.06798875279541244, 0.0978387129668797,
0.10181239003862498, 0.006792858082142025, 0.09869498462525983, 0.22812057969773913, 0.13266671190716556],
[0.15566307960320144, 0.2367540788805075, 0.018306707786896675, 0.3130512176732618, 0.05007300518882156,
0.18726889137787367, 0.06459430116208427, 0.19245354147035426, 0.0828352419083046, 0.005849669493655892,
0.22417987718151552, 0.15490154864664615, 0.13790316178482198, 0.19857115212024787, 0.18270448210031706,
0.015278233582509404, 0.12603493146036301, 0.1523017511198131, 0.0724785854448344, 0.15281266011350977,
0.3254692420656815, 0.2764910027343829, 0.32753561815166743, 0.407236909705729, 0.1341433556562336,
0.004951782724650905, 0.032994425714093514, 0.11413731339475343, 0.1572324415204205, 0.07264688791048374,
0.06489746713429592, 0.12696270831225712, 0.26149206253687585, 0.16177901704011594, 0.13142853045039266,
0.18600995165902492, 0.25234099254241593, 0.10569629557686502, 0.05983244063433111, 0.10747519720518584,
0.13535798401529522, 0.21799137377503994, 0.07193242547311055, 0.09693328400055184, 0.1090913216974972,
0.10667103447415271, 0.1762838089713566, 0.16085357533868322, 0.018020536487613346, 0.044012862680988536,
0.056023702793264685, 0.016234925371218394, 0.06688343952588464, 0.04649536596650611, 0.008047855208758165,
0.0002452264841847713, 0.03859565108377249, 0.02874153639479733, 0.03131676992091738, 0.07332743932598212,
0.05891101692045718, 0.09326745948621967, 0.14157410403787188, 0.137314710806358, 0.15415907040527074,
0.14330125792358955, 0.04957002572329271, 0.0025765601149772503, 0.003105181808305113, 0.03725605109546309,
0.004312075601201822, 0.001524102208465763, 0.08186325656186264, 0.180148652704489, 0.1403919750548641,
0.06117469216877677, 0.061693167984261055, 0.1323970803054631, 0.17922022369404939, 0.10692077601286107,
0.056304845689918274, 0.0456818553675322, 0.10425382379886979, 0.21615022608687695, 0.25028575645659173,
0.221701503655031, 0.16833602288944244, 0.10891108458725107, 0.05725129127917507, 0.20122257961983622,
0.30879403726346466, 0.37352593956386565, 0.2522125715945046, 0.18872836998468168, 0.029308105355798263,
0.020441167812371163, 0.09990015993856449, 0.07854466877630142, 0.04088846944277234, 0.034624887294464064,
0.09764832337400663, 0.12042972669761916, 0.13572746319450577, 0.10229221094726679, 0.10262248103699988,
0.29624299928549896, 0.2278970597189931, 0.3229423736839025, 0.19895652838350739, 0.13379721593886806,
0.08641971628215053, 0.03597587991262483, 0.1075074883302619, 0.15455403406617113, 0.11467975656332476,
0.12302543356616386, 0.07314384811427078, 0.043991363359451995, 0.023658631868669035, 0.029233035380981928,
0.08488162985619294, 0.016252803547231964, 0.0014512089002864748, 0.0757904770578991, 0.23330548439930243],
[0.2884405578050437, 0.19042408513711503, 0.15588931650132118, 0.16550918704146292, 0.2021670904934177,
0.3631454270214088, 0.03596060412120022, 0.11721260859797515, 0.05198842522088502, 0.20205056327511828,
0.14115115694316968, 0.2559716249677917, 0.13539451419091156, 0.33784120735568796, 0.36113101030025796,
0.0022610387291701842, 0.1932606461761518, 0.037520110630068396, 0.042050461699243626, 0.30472256655356944,
0.44592266383290813, 0.41141074335676453, 0.17551867180699865, 0.4075303496373789, 0.22979983895683948,
0.03466310346181458, 0.16765307047530972, 0.013302003547178603, 0.183001075880736, 0.1286550539042068,
0.09420518709108619, 0.19188581316777822, 0.1359488316160809, 0.17055732547306302, 0.12743845621395894,
0.2099342933875455, 0.017614636935134244, 0.06269161871864727, 0.26496305166108775, 0.06125332621952041,
0.21088176224511965, 0.21224695411334446, 0.047639640708140675, 0.10843176817300927, 0.013470992383918856,
0.05619750528824975, 0.09430952655580138, 0.1342808995102542, 0.003107528465413504, 0.020006064927843514,
0.06154554465641958, 0.06330244165111883, 0.011468796994530957, 0.00480860522004705, 0.015626371558526923,
0.006044512608956869, 0.034603303832313505, 0.031051151450511554, 0.035655340788677006, 0.035576393383198165,
0.09962491441235365, 0.1499231173038855, 0.21200164050718345, 0.19268673237431183, 0.1556976695618108,
0.14931867788523048, 0.05431957186887262, 0.016009341205442007, 0.08619182352872955, 0.16307233752561892,
0.15116233366971177, 0.13958240362969693, 0.09758721748977435, 0.09556242482475455, 0.11536852961722899,
0.1363460979233745, 0.10528536815761286, 0.04646754325805386, 0.02843788536029319, 0.017781584861933375,
0.05132370756475835, 0.1173864348391585, 0.17863497741642304, 0.22867963230829114, 0.2297889574577262,
0.24735011981419014, 0.20031733816200653, 0.1761901430887793, 0.16727524990205933, 0.003924716894929545,
0.09413490520116406, 0.14979297919687343, 0.003867400129886208, 0.03681905814225923, 0.03465233051422911,
0.026512358834521504, 0.10203562646803399, 0.021432335565188997, 0.018461443631390115, 0.09589098168548028,

0.1476530904722748, 0.1581119905240602, 0.1129649002010046, 0.015495169496113048, 0.01866296752138078,
0.15389786799165941, 0.13394678480098177, 0.3029702551517467, 0.15729682807596987, 0.23103701448051114,
0.25126337434989626, 0.18925827027721054, 0.11971196920602827, 0.03124446432602624, 0.024492003590747245,
0.08094985322521593, 0.04763484444510981, 0.08081928974310844, 0.0500762381218679, 0.08729871936818986,
0.06748214215287891, 0.11619233580110572, 0.16138188772554807, 0.19029235552049867, 0.028214001783448996],
[0.2566835977892607, 0.23301770615544748, 0.12099025119930591, 0.17257060623439335, 0.19201144948807125,
0.03118028012313888, 0.11619222872387458, 0.06586584923746996, 0.024780272854442866, 0.10571809187178871,
0.26573691875900646, 0.032328829987431494, 0.22728128765456798, 0.4015178320484435, 0.27000239361043904,
0.030076874905804993, 0.04308822243819954, 0.11391511046295355, 0.035989910488769046, 0.218592486249351,
0.3232542887539697, 0.28072963365524967, 0.2886606328811143, 0.45253274951462236, 0.17293972856958736,
0.0364267069007221, 0.01466481848907104, 0.07092488973203875, 0.2550349243090662, 0.08701428565534913,
0.03862542338956262, 0.20851486077027617, 0.2096378739869585, 0.18647544380309974, 0.07055762138949835,
0.17320588299071724, 0.05640213311701639, 0.103810450653432, 0.13096380263897645, 0.010276459382415464,
0.10265283903683478, 0.10839482564195127, 0.0681139264825127, 0.034538005997619955, 0.042398247879855085,
0.06726931292805889, 0.16260770966624427, 0.23387568052991872, 0.024040306381725076, 0.031380530912897546,
0.0015824968285781669, 0.01817150012307877, 0.05018107606393468, 0.023764653719582186, 0.07628276357498061,
0.053126711230973796, 0.05132030388960888, 0.09316601198252651, 0.09336099744639695, 0.07520683707876744,
0.12282334684717011, 0.19922926433059335, 0.22416478678261695, 0.20345758961657, 0.1449805396588711,
0.12927253498438365, 0.04110609446258507, 0.03150533768740997, 0.10787174864084668, 0.179790930907369,
0.15592601555682684, 0.1576497255113281, 0.10783678486798308, 0.05706048809044538, 0.05189956329061008,
0.03754206385122574, 0.03185536699925385, 0.017705869719954893, 0.013578089541956505, 0.028480392254822812,
0.00042199603682600964, 0.09697566325507913, 0.1981329056728151, 0.2387521389830438, 0.22826172667746264,
0.22746907566316818, 0.16695340427153743, 0.12944988423995063, 0.07047996150996218, 0.0831054992291476,
0.20944292524790023, 0.27209055689215794, 0.148744387504457, 0.07710551691698617, 0.06435123678161456,
0.030058638422351867, 0.015591558946835468, 0.007884354511117315, 0.047672019261049164, 0.027580821501459407,
0.04134920982451 6654, 0.01013069420892582, 0.011476229629079595, 0.015976223767452873, 0.028353470201040547,
0.1786426854701243, 0.17666841754054058, 0.3626475764913496, 0.24408750606815577, 0.23913389772780116,
0.2395418131954463, 0.150669070398757, 0.025796679154735148, 0.03465207760725338, 0.05323595514280011,
0.014044358258567424, 0.0465206837929799, 0.028439794642148755, 0.03136838578553041, 0.015097519318544514,
0.007705034564329715, 0.1566369713762484, 0.22082648965403662, 0.18293052865975512, 0.036469044411583815],
[0.3228646650778665, 0.17389218543437895, 0.004328901023044 2545, 0.2641283076368257, 0.2851582325429946,
0.24767663356674344, 0.02278479716971371, 0.31152222175311045, 0.010975715950578955, 0.04187861464470783,
0.2659136094608402, 0.27650273897593397, 0.13524753787624494, 0.2923866511068254, 0.20124802308578343,
0.005569543839775228, 0.11823965284109322, 0.05853727836732191, 0.07578086424604265, 0.22882149095547188,
0.39732394476240745, 0.2668422385665749, 0.24980308913479712, 0.31243507186383696, 0.09948370384568596,
0.025918103816509493, 0.08195427654162381, 0.038504009762357065, 0.1508321542810258, 0.027036803394233526,
0.05602199101794849, 0.20383274004315785, 0.24209717908732403, 0.07916391488716777, 0.01625818642095 6458,
0.18392250881956196, 0.17947232312114256, 0.049431172758865174, 0.19453217894106903, 0.06604711275153756,
0.25370489384021294, 0.19393428047420203, 0.11902166736163201, 0.08208416063603695, 0.15233226345619175,
0.050711363068872015, 0.035778224026768675, 0.1773139969538798, 0.012529159612114172, 0.023970716015760708,
0.11774186794481284, 0.10541899672531435, 0.1318777013061464, 0.09313571894413841, 0.08955603261189986,
0.05695543348472689, 0.0021622188319431915, 0.01946569824808491, 0.009732013888895573, 0.05839730322191656,
0.02845684410924137, 0.06412317690813853, 0.11040718875138632, 0.11953001522846633, 0.0953377144441306,
0.06790619056645252, 0.05574736472191582, 0.058773657353601995, 0.04860529197973442, 0.019987186121694152,
0.0029231364325420185, 0.06486555755064916, 0.15909104854407655, 0.24126651433583912, 0.1983479769253091,
0.0605050324239 2833, 0.01543674108355 3893, 0.02647998767595768, 0.011853527516285356, 0.03755598767134357,
0.006387079771091 5556, 0.05041051373186401, 0.1511137251194851, 0.20130520925584466, 0.19987964426149016,
0.16864890439565658, 0.14419439968080042, 0.11717564598760244, 0.08999501381102226, 0.04940763961810978,
0.0952151153595132, 0.15326338697802092, 0.016496272191184286, 0.02033262028467389, 0.04761056035950828,
0.0652787493469651, 0.07295341576397579, 0.05563829403726641, 0.04466139638593131, 0.028048367076112687,
0.05748271901208 4675, 0.08821976045543324, 0.1456287208743064, 0.0331161818559978, 0.04548495575867613,
0.009569412348366921, 0.02628854004868653, 0.1479031360148848, 0.0040824646394368, 0.0502879654116168,
0.14005223972244013, 0.1341482604852726, 0.04152937741713862, 0.027407341516287702, 0.020680461952245793,
0.10533234977121249, 0.09093437849302403, 0.13612102350347663, 0.14924991261574663, 0.1877492987275344,
0.19157939756128872, 0.05951254620428011, 0.1670182774128062, 0.24048836444457405, 0.2085475700058823],
[0.22287726218798853, 0.14330060954751014, 0.0779007346400296, 0.17582609123638152, 0.29898252342758874,
0.2414332181779553, 0.05294536777959435, 0.15659750009267617, 0.037869153223236446, 0.15432178021157567,
0.05075245037441287, 0.11193657545326484, 0.19541101584246082, 0.2873331332629844, 0.2491810041363136,
0.14997386858508105, 0.04156429666793549, 0.172457519375673, 0.04561740431091506, 0.10162281734365562,
0.6008449387182995, 0.18934862235820898, 0.17845190113732753, 0.49224946191247226, 0.08413828424461742,
0.08653240569007642, 0.028045019063251183, 0.054000703595876026, 0.10158652000090318, 0.08941674764476093,
0.1360551227529369, 0.09592892656705057, 0.3171863883014486, 0.21313438374705665, 0.0008883259280842,
0.04637239767997561, 0.09965348654554508, 0.05932063675992786, 0.30285974040516045, 0.26443817037014816,
0.3682474079761512, 0.11936671402902999, 0.1557646025612315, 0.07973728057054309, 0.1489946254647259,
0.037784644922906056, 0.10078685423082868, 0.2660001731989862, 0.11171330301807664, 0.07511092223236945,
0.08259212481049015, 0.05149116853876666, 0.037096363543628, 0.06635291774604263, 0.008471369580037604,
0.021840718466342283, 0.046237770038940175, 0.10904173981020805, 0.06120856127863736, 0.11918962931453513,
0.007042240170421668, 0.10329526280084819, 0.15110489636153993, 0.18267025348838722, 0.17622528038637914,
0.13532072014272936, 0.05601916870442332, 0.011543401414678434, 0.0018236734092661356, 0.06722725478887862,
0.07159257012106847, 0.07062646771741067, 0.08444660574420895, 0.09903481040215432, 0.08782834847371424,
0.10787151063663862, 0.08441277586985793, 0.06795229662353376, 0.05418678065634 1735, 0.05159453729313441,
0.07558332351821855, 0.145734071094665, 0.23995060220651224, 0.26255880104358464, 0.25820601558879946,
0.19754215421785545, 0.1407987673468643, 0.0620136007516434, 0.0305616151952459, 0.17034515815522935,
0.2339123772269625, 0.31953843321817377, 0.16028305038490323, 0.11043274637621889, 0.01319193850076815,
0.007614210143825208, 0.02466880639136807, 0.03130344880568607, 0.034516429371415, 0.013605917746948928,
0.0811110929592 7223, 0.15976366048643853, 0.14331160023681339, 0.08633767975514806, 0.12168950448367546,
0.24674612781992958, 0.2292643198736336, 0.36343254694869576, 0.23305857068281285, 0.1628184218612327,
0.13093189906883412, 0.016524257730881457, 0.06083203839592326, 0.10429989285169669, 0.060862678102707374,
0.0352008398839109, 0.02002465832312331, 0.0349718206169629 5, 0.040621755444831466, 0.03598631941595328,
0.06282757749611183, 0.07641901317578187, 0.1291364336971981, 0.18626161881382317, 0.14153421630324292],
[0.05245313616669292, 0.05654274459075142, 0.02354763560840691, 0.10251922872736322, 0.095170430042941,
0.17745423919535935, 0.173550870089402, 0.026190376700713964, 0.06077496427148004, 0.02849044475322885,
0.11757655494047596, 0.011150523325237643, 0.1283281615616 3504, 0.12705768021104344, 0.22789463999333925,

0.17568072747331645, 0.11669602402246904, 0.0608979770347096, 0.3833570958809014, 0.21721063289411957,
0.3383770480906858, 0.25243984657213986, 0.29969135600301344, 0.4871479306272196, 0.10776984221376561,
0.0731443454337275, 0.025123202481046984, 0.1272172983688527, 0.11458258926938576, 0.08241397192054366,
0.11706013577301193, 0.32772262344334707, 0.284506254388309, 0.02400522242125759, 0.010495494173212177,
0.09903965814733325, 0.07741672943670226, 0.03694362749689619, 0.270889672157661, 0.23795924742932206,
0.10769751841338987, 0.03830855838953435, 0.18323736405769897, 0.0804620286225751, 0.0658859389170365,
0.020263527251442683, 0.17626872824636658, 0.22083047622504637, 0.08995257544562144, 0.07640385939213737,
0.08393710913868986, 0.05426279878638413, 0.011860220349151991, 0.053117342363805144, 0.00994388879984844,
0.0228708195334391, 0.05336850750795045, 0.03598957505466925, 0.029443860246269188, 0.012657233220942594,
0.014953802700895044, 0.08338867028359025, 0.1252152731425549, 0.10929579571443437, 0.12900924097950303,
0.1388426172065677, 0.08102152802634485, 0.051357653958371025, 0.030647765281770108, 0.010106322202943205,
0.006930894415305402, 0.026970614414741037, 0.0021076832657255778, 0.006174904750567878, 0.05350997843725433,
0.06286280940999978, 0.0014960548070665131, 0.10470169794690731, 0.17936656189601807, 0.22358895571521015,
0.24316045516510607, 0.22414609038411615, 0.23099012731082397, 0.24044416432300283, 0.23496802299453273,
0.2080098564508869, 0.18044080303797405, 0.17171440118161724, 0.07021453590551424, 0.04599709783206941,
0.12795920463749427, 0.18175272894681246, 0.10763168567512764, 0.08838451746828015, 0.04050156794611525,
0.02090665207177152, 0.055423224766913054, 0.009966498080287054, 0.053956721883657674, 0.0020845532393903615,
0.01834272603880363, 0.03352340674378461, 0.03587250129324944, 0.0649882069005877, 0.047138494305119676,
0.2578789220617092, 0.2832557690208054, 0.3881802287244382, 0.3301100730391961, 0.27860786304148916,
0.22289429102322464, 0.10061797030783769, 0.003261508144693994, 0.0549073878045971, 0.010856128536178476,
0.004521098291086018, 0.022682737296431773, 0.05700251737062417, 0.06514744374649883, 0.057745675791496455,
0.013443717727723788, 0.009233569824764025, 0.04445392747041022, 0.08177259666558621, 0.20524243703314546],
[0.3057405455715629, 0.283180001363881, 0.05496392588727379, 0.4259686570605452, 0.1924140556181919,
0.24617422655433707, 0.030552486580611448, 0.33879567198650606, 0.16159609738423664, 0.14046960637901829,
0.2604715134569859, 0.21319746655931054, 0.10896232526342656, 0.15300950453726234, 0.1953211341014369,
0.19744765993334415, 0.011202204970289132, 0.11340878167367038, 0.009499426251356875, 0.02191864123833962,
0.40591325505994147, 0.3024627934427591, 0.2395832068330811, 0.4716159537058954, 0.08530958409092898,
0.06958601077671757, 0.029741536586087945, 0.010398044535822744, 0.18199864562932197, 0.05727150576582839,
0.26801373441273996, 0.20039442561260623, 0.35702889264292315, 0.0343904505556081, 0.030076780814192274,
0.04723663602462895, 0.12490932479793165, 0.12232591993989347, 0.2243872595662793, 0.12433958886868238,
0.21307320623172604, 0.033661628516368274, 0.23016734852651283, 0.02560549783051659, 0.013091410611102295,
0.02439997183169258, 0.09804271436031475, 0.2546554333147036, 0.05118181470308469, 0.016065591586708484,
0.08697220067391832, 0.050967423190320604, 0.002695288125741386, 0.017203402727404758, 0.08221139256281172,
0.10971412455051405, 0.04954108821268581, 0.03518734166281004, 0.08608727767504946, 0.007925484440421718,
0.0528853611157842, 0.17914352232510405, 0.16964147496147972, 0.1589977772177102, 0.15134025605604742,
0.142031456706442, 0.050284664400304116, 0.0014501729518057852, 0.013099822392181128, 0.0365700350064199,
0.03909285420664558, 0.060500802435164945, 0.11618202564532813, 0.22790212895225248, 0.17115785576877354,
0.07190709708876766, 0.08409703079156035, 0.12964402516527723, 0.12301684845876404, 0.10938964898400629,
0.11596739653932613, 0.11171594149746483, 0.11590009448260043, 0.12783012377548633, 0.12010600275307812,
0.04655930095952318, 0.06534618127136936, 0.0303780812719036, 0.010487556667068293, 0.11502918775291995,
0.19639834461552727, 0.24699473715003423, 0.09706984872464654, 0.013109854722358678, 0.04904267819258208,
0.02070354911048403, 0.04346437655048405, 0.04301864174385073, 0.07838693323632277, 0.02658353558684631,
0.029249537453776196, 0.07408823606039126, 0.10575440024244374, 0.039799286515611314, 0.0947153149643744,
0.19267341423808018, 0.16835874085884722, 0.29352833209832774, 0.2105449453984405, 0.15680324101452622,
0.11726911940321548, 0.04909826087617114, 0.0109640674468596, 0.08729419219535546, 0.03350876394295345,
0.010543456283452945, 0.012335522888480784, 0.03132930609422536, 0.0664868843571143, 0.07214712466185551,
0.08552874521722033, 0.08863646195798634, 0.11207218492371446, 0.14518642033886006, 0.06181248312406916],
[0.35049164953672307, 0.1175427918365461, 0.05590145678852562, 0.3469560549955182, 0.13477738759169763,
0.13577901591665, 0.01470526997237595, 0.23333052767422247, 0.17196643135004708, 0.029274212678280214,
0.19515311191404847, 0.08994985543257399, 0.23915163289644534, 0.3448319811257364, 0.2224523618775338,
0.141959263291998, 0.02680155314724447, 0.10801110016561272, 0.012697323005748383, 0.06677668560338583,
0.42883200080433687, 0.2739454701039391, 0.18016896464587004, 0.18901132459265133, 0.12289595790186875,
0.11759953756036352, 0.10482664258842415, 0.13541680971678194, 0.22602506550218965, 0.009759461795630092,
0.06317956073361408, 0.21457045715342754, 0.3038779346384961, 0.008018493121527411, 0.0945740174755651,
0.1448406793272076, 0.16149938072187112, 0.02826253846625332, 0.19848840744160642, 0.0998711959768002,
0.13490960924584472, 0.10066439178458111, 0.09998798416547065, 0.08507478422358268, 0.04798849307728618,
0.061054608179559745, 0.06243814725630332, 0.19998748926592833, 0.0003802494139402369, 0.010378125868111298,
0.0970665335348517, 0.055362314667446406, 0.009009515911547784, 0.02608180709732729, 0.0035307839180045964,
0.02250174138551394, 0.029338771064608977, 0.04002812597255159, 0.05158346048294347, 0.035689718714957464,
0.055782219455372865, 0.1352556413215152, 0.19338355971445823, 0.22848479303074015, 0.23499852597581544,
0.21290949692664984, 0.14666729490293504, 0.07592488076539138, 0.07092298684756719, 0.015886326207965894,
0.020966448745642907, 0.09788949041872433, 0.20304722444474083, 0.3114171972938826, 0.2662028031775618,
0.15412272264221788, 0.12341074306141882, 0.049941833105948116, 0.012346632342916268, 0.0559377601873102,
0.010521984608405286, 0.007941869601652267, 0.08336597138297423, 0.14010064589566032, 0.1362095873456068,
0.13640866418033862, 0.15895202484274631, 0.1518929347655661, 0.09351958874014484, 0.0331334782909971,
0.08300193402357586, 0.14034716462368182, 0.08953890015879487, 0.04936850498455126, 0.02711532057179193,
0.007770198326667233, 0.0318370121282187, 0.03239459796893293, 0.04594627609078142, 0.00899959820517184,
0.053083468566718015, 0.015331945345866044, 0.04599370416006648, 0.14613197513481938, 0.12429331226450077,
0.054409602390884576, 0.061932975076753334, 0.2808755160947356, 0.23157676998096344, 0.2845811979814107,
0.3705069625867675, 0.24273365250543494, 0.09298832708956528, 0.016055621653121417, 0.07495084861715408,
0.047860315128033196, 0.07598007485810955, 0.04332904979291104, 0.0030839058497895908, 0.04594437496784492,
0.06348334537065345, 0.01240065362902285, 0.09548414027941299, 0.17042958880678677, 0.20379778030290824],
[0.20365200871363529, 0.11626372027417323, 0.038277969691742304, 0.4771908735675695, 0.02208903681586717,
0.21777032431192272, 0.11268927824159646, 0.3346810820822587, 0.0670791772839345, 0.02304920007219664,
0.07119264619660026, 0.15943441317429372, 0.25553790851818803, 0.37366439985486133, 0.18854107367250222,
0.0022276783569627947, 0.018699362718372147, 0.15400763479391863, 0.05500406997455429, 0.07749438170898838,
0.40824696352104667, 0.3902240895188313, 0.34222027010263156, 0.4622499888520136, 0.058484858294133524,
0.008326419213455227, 0.15749360344950092, 0.05996629461362673, 0.21554018562712038, 0.03332546790780356,
0.050312757176919444, 0.28029078345248293, 0.38271753956426025, 0.0226467149779481, 0.052233561834211795,
0.12214986156715225, 0.020700571632382623, 0.18638847221450683, 0.2662155764416838, 0.11594794474409607,
0.14869248004173063, 0.048401640352405344, 0.1191452560190316, 0.010116723222563241, 0.03765181521979012,
0.030571110018373645, 0.017692117527621412, 0.19434587905337575, 0.028125498090935966, 0.01344203421511443,
0.08507677998181398, 0.10926197304365921, 0.025700047227596258, 0.013441804937213718, 0.02558267054576346,

0.014421972735878247, 0.034444803614437663, 0.0038798451887292486, 0.07684964318579633,
2.7818173334719364e-05, 0.027143453946034724, 0.1009478508518321, 0.1392788584770962, 0.15069135683562054,
0.15663295125948834, 0.127494028404006, 0.050975302585892834, 0.004506882431006718, 0.01291316687506099,
0.05080927034370655, 0.0651898870000526, 0.08612912617822102, 0.06218018302377807, 0.07786363133852144,
0.017350771997622465, 0.011430155125643741, 0.03663619830085432, 0.03918415867815315, 0.10408637509114402,
0.07281676018647393, 0.09516680264915967, 0.11809507464542252, 0.15338726905785122, 0.18689420390513467,
0.17819506388436707, 0.1474894987932527, 0.17370129102486462, 0.17833558979987585, 0.11196747676231947,
0.029051062103975074, 0.07368597768479436, 0.16570643245568226, 0.09830823115196566, 0.07376226640912051,
0.0028125979877998315, 0.025468309148806823, 0.08137998046556033, 0.10280076899592783, 0.007149184853489737,
0.04217170595031474, 0.05419967075366156, 0.02981555757282615, 0.09551912088333092, 0.23471532843924023,
0.22113404233176615, 0.08903193915045977, 0.11421791163784101, 0.40826601414750124, 0.3927666479662863,
0.419113747108812, 0.464548597950747, 0.29369189197805373, 0.09559607825086838, 0.05433853762907657,
0.13305004598327902, 0.13492278255517598, 0.1435792350787963, 0.07270210807815397, 0.04670463102347642,
0.033071362097095436, 0.007325955012085452, 0.054643729426607474, 0.07516959131001136, 0.22592346463812804,
0.16348130146251144]], [0.3356711401377877, 0.12680159578430333, 0.14529044540099204, 0.14235767528478074,
0.13649818406637645, 0.07521893304642754, 0.21507898410165346, 0.14513489716346573, 0.14338055647458423,
0.004087445704987144, 0.029565834006837353, 0.06883024269947928, 0.28706032410821425, 0.5264610279983923,
0.3463030714778516, 0.14521707965509173, 0.10696846287140097, 0.14646012800840513, 0.010250480979179306,
0.07454991497626128, 0.45915262030569176, 0.3185653672141057, 0.27700246726187194, 0.3699552110168354,
0.14990330664151477, 0.15091708087763728, 0.050556223711592546, 0.041750448489548106, 0.06484870048024337,
0.03075385304862669, 0.014056960761142455, 0.04674766058400973, 0.25038769602998084, 0.09669365412290089,
0.1037924539591307, 0.2781434408863248, 0.18553869146199234, 0.060416644733961106, 0.23808713125440364,
0.13950274766331724, 0.2720709646339882, 0.17715286481788836, 0.07050345084023465, 0.0494310244113636,
0.01477548637752936, 0.04188028303886979, 0.04485775742388912, 0.20227298102546598, 0.0003783138114599651,
0.043028812864241, 0.033672038226767946, 0.08828021668594646, 0.09583912593379923, 0.03696269687440391,
0.1137359096112591, 0.11565898562128141, 0.07418225704044308, 0.021294064536764884, 0.04553656378712795,
0.0395356079501309, 0.11445093707208018, 0.14074333467492206, 0.16043975661467008, 0.14642462435179468,
0.10705005118612408, 0.10148311053356304, 0.025042339724931948, 0.02128744189160542, 0.05611400481061307,
0.11613036574358494, 0.13724866363585314, 0.1632634184181966, 0.15229479982193964, 0.1298050884572359,
0.10523441137593414, 0.10359077060679958, 0.06955744655588346, 0.07766583566972114, 0.08910205654373099,
0.08098090684633313, 0.043391174726808014, 0.15177414202138378, 0.2096836533692994, 0.20915086177877285,
0.19493743222126145, 0.1693245238589084, 0.17440808813389258, 0.11447624584613494, 0.04602436293173518,
0.09304862531188954, 0.14787672304563826, 0.18832410213512357, 0.06887743519734157, 0.06478900660666798,
0.01574507789291457, 0.01704989984559248, 0.028745306282221113, 0.0145407216983002, 0.04241836183888207,
0.04680476624335365, 0.0514594629998728, 0.13912329085143052, 0.1650564391116719, 0.18945495090836453,
0.22897449322331984, 0.29318855491206547, 0.2270421975135361, 0.32690653671489606, 0.17092246580664813,
0.14815308607740957, 0.19547416983957489, 0.14746359058570999, 0.09010942198745808, 0.05671971340225434,
0.08234453115327602, 0.08401483517494936, 0.09238561893241651, 0.09154300305738616, 0.05224131707435521,
0.068503553020743, 0.019862223261785522, 0.08062460016668993, 0.16846574612050264, 0.2752823654819479,
0.28266887925211215]], [0.32625430975632885, 0.049434144844141416, 0.2211592266787564, 0.10751463357051737,
0.2909144293737297, 0.44593808759647113, 0.10955512507675937, 0.07393696534843582, 0.15258593935858217,
0.16558859923730618, 0.1517121191426629, 0.022164665374594144, 0.2940992737440238, 0.2648148783403012,
0.43449033642822416, 0.05016587051647978, 0.057109501605520224, 0.19082762793678879, 0.005516149466676469,
0.14418631611624924, 0.40888697389852074, 0.3684917798406463, 0.1816367171340858, 0.3540465938361963,
0.042396756776573195, 0.09376013774140939, 0.003840700694538237, 0.13143725144827373, 0.22569085737661937,
0.0812162334129705, 0.015362721613574141, 0.15780110506843956, 0.3179493147356135, 0.1903606513195203,
0.24976375955562066, 0.26069670952755536, 0.0390839450085215, 0.0888265069152937, 0.2921168042090326,
0.07754658035784691, 0.2010754320701368, 0.141891523940125, 0.01638441823641696, 0.14413796857856068,
0.1870330888746777, 0.25792858161029514, 0.024679132416207443, 0.14394672481835377, 0.029192378298450852,
0.0384119311523875, 0.11641741690945509, 0.11409412594598062, 0.04811343994410754, 0.029036479055199484,
0.06675608532346346, 0.06706482952536823, 0.03748912153316675, 0.04625604953084938, 0.07525454856447479,
0.08044793542705901, 0.07811382825595468, 0.014759321531532124, 0.09329431426901605, 0.14540957559096776,
0.12643773387423438, 0.13292168105000943, 0.0913369593253359, 0.05295201806943964, 0.05011746465424025,
0.0456069571226371, 0.0756014797319987, 0.016224470236577554, 0.06876177874507883, 0.137660826156588,
0.12301842247303436, 0.10666608470154247, 0.05813764191456304, 0.12534339100754238, 0.19469394948336272,
0.048728166345286796, 0.027621532794431525, 0.019006064989304666, 0.04376255555968458, 0.10827414561415069,
0.1407688377861714, 0.1737310326205703, 0.15906607226648573, 0.08892961962536099, 0.0520499390009261,
0.2509669353509823, 0.30969345369283424, 0.3840351067129484, 0.2179479059772851, 0.09299389317300384,
0.03424794166137405, 0.1444242953293676, 0.2700983503532775, 0.19712585408076772, 0.06293914748595181,
0.12348407382731565, 0.13178411116655508, 0.14200453762199616, 0.17522806480445893, 0.054251172152959076,
0.01250592633193244, 0.009329974359644553, 0.14717165756549658, 0.38807826234520304, 0.3124058715376006,
0.2453081063364489, 0.21387896387755195, 0.05256092518025452, 0.050561509498157256, 0.14044937885631037,
0.17216587370303385, 0.07953952580596534, 0.10076121526589489, 0.02811191298043623, 0.0069052982131480745,
0.06714373449178813, 0.009366996101778595, 0.013817690141213868, 0.02220414231864283, 0.17448215048725918,
0.22985416242814422]], [0.6612718252471717, 0.1352476661473441, 0.08405719330695857, 0.33332161169150526,
0.42628333388846934, 0.24616883979016202, 0.015722029694562576, 0.29983741783865825, 0.0412952212294947,
0.1377557806519869, 0.04595616839902517, 0.40571811315778494, 0.38303512600563433, 0.48102814636185215,
0.39408168114404496, 0.03189466628432356, 0.06574182092102525, 0.23488039448929812, 0.0460305270827816,
0.12133074665926993, 0.37089254919547915, 0.4453398715544558, 0.2086209890124479, 0.39856934446957804,
0.12126335661402841, 0.07070705935959241, 0.09573552195512659, 0.00620849776896266, 0.2945643178260967,
0.017234094459257488, 0.09757707597728664, 0.05007797557705277, 0.2528029749444719, 0.11018165097344965,
0.02677183928842308, 0.3679037957143092, 0.1796304947755118, 0.014545563483791754, 0.24729899759102697,
0.10750253132627156, 0.25244027154080634, 0.22495851809495565, 0.028563117321272345, 0.024817000356187495,
0.027778242570896013, 0.03685422917690557, 0.03422015207133121, 0.1853599049782101, 0.01668418269958743,
0.008728072997689556, 0.09981670179239042, 0.09403606218547983, 0.045471709812379565, 0.04368388193507731,
0.032165593654413835, 0.05104570642269582, 0.015341114567719465, 0.05432520268432875, 0.01680748185928887,
0.006877809176696849, 0.062278112040794975, 0.14494803257310904, 0.2129068309018582, 0.24516158164311078,
0.18360955277391106, 0.16299537472703904, 0.10814926718721846, 0.049074535208292365, 0.0016475949740138375,
0.059403613919866434, 0.05925737085949045, 0.08764758354439652, 0.11933327060566509, 0.15155466204492415,
0.1398099680722147, 0.1121236460386983, 0.06308644811806341, 0.06119986584330909, 0.09360755810893924,
0.016662766053313696, 0.01492230704227453, 0.04801577921463153, 0.1374883300957783, 0.19126196140496085,
0.13660502371369954, 0.12659676686274657, 0.13964411527453086, 0.10192167932610648, 0.06091286881141691,
0.07050787689842307, 0.09239839310492129, 0.19395726069260388, 0.10267002764348943, 0.0878924578193623,

0.07195644305715838, 0.03696764176987227, 0.02830169792989704, 0.03482424147526492, 0.05549415404088839,
0.10082472224004567, 0.1313662264703836, 0.15215023462091276, 0.177383739542183, 0.026186829721441277,
0.03402811107266202, 0.0757921543445904, 0.04468276915618813, 0.2320654783243456, 0.13332092913504912,
0.211345671991866, 0.2624595036379246, 0.2307535935392048, 0.11043509462883018, 0.050867339670265986,
0.020083406434819234, 0.022527012397419697, 0.06206176127832441, 0.023727704501046963, 0.011850798701642575,
0.0636611247174552, 0.11269473751281707, 0.10999167471536016, 0.19331407114607987, 0.23268233577363903,
0.06619132923067945], [0.48242404750937856, 0.011467893588178985, 0.04065906272408884, 0.42194063915246827,
0.2196945052972142, 0.2174911874887361, 0.20735375874212564, 0.30021399235640633, 0.14629696153425398,
0.06651400865926552, 0.13916872966923727, 0.23487503348331987, 0.2780810906655646, 0.23867225693462965,
0.40072248971550245, 0.019538852268110116, 0.007421484070425523, 0.2653357556894597, 0.059651822030288536,
0.09919967447391699, 0.36927574317619344, 0.4364186148568386, 0.34782132428819257, 0.09261104966522091,
0.10335668246271584, 0.1481172151948012, 0.0981879257259228, 0.07168019145028455, 0.1723220740089459,
0.023738797303261384, 0.032617532514327224, 0.021940224042213477, 0.32022410162487036, 0.2213085563445546,
0.09929789919455627, 0.2805032477450103, 0.12026732968536659, 0.07434974218792381, 0.26910288843424995,
0.06672205092359257, 0.1236557071547497, 0.1168359681388474, 0.03180260434489392, 0.08118429820278833,
0.04639876602599095, 0.03418493574769266, 0.05983631067754209, 0.14651994334911267, 0.007902775744168419,
0.044085605912398525, 0.12288352412067179, 0.11238864144699157, 0.07160980215545694, 0.015274050786145086,
0.044091853975044275, 0.0782494418215229, 0.06019224264469556, 0.014405817347658237, 0.04142305460825723,
0.002471624523994912, 0.01914730560346896, 0.10914126462283769, 0.1704537179821004, 0.20282130519814368,
0.17188899855066248, 0.12220818027456332, 0.051303494602920385, 0.002685099344567735‍7, 0.034473117696110875,
0.08154448715548017, 0.08097826952765119, 0.06713457168928016, 0.08678273037374937, 0.14195925082185631,
0.14417376142404825, 0.12801008431008679, 0.1169102844602301, 0.0735172987099299, 0.02238350625190826,
0.018467158670961117, 0.017716435024746107, 0.09884691569030885, 0.16127714496453718, 0.18040204345444452,
0.16253397188565397, 0.1948187509596857, 0.21434260314882134, 0.1674545683112199, 0.114441618613465‍4,
0.03483627212840834, 0.10447105115855944, 0.21011692755847614, 0.07834613670359944, 0.10531300834586316,
0.03517542076866961, 0.009830840992291307, 0.05369724140416177, 0.006404861169554905, 0.04221533984493705,
0.08750313606305492, 0.09523739616880166, 0.09572576441599176, 0.04788265721160137, 0.03249389990743508,
0.07472913607143945, 0.17174569721106894, 0.22569038677818995, 0.36638520583050427, 0.24442716286869068,
0.2657407264394268, 0.2794110089456389, 0.09744084788113497, 0.02247007267560619, 0.0515876107118384‍3,
0.07852883527066586, 0.054573423368377984, 0.0832411996258231, 0.019869566272085085, 0.007276715546415803,
0.076019779603539‍2, 0.1322150571006099, 0.06559702309654931, 0.15378006218031‍3, 0.223984016429732‍8,
0.18148788248702633], [0.3520733468180326, 0.17278589560333296, 0.03719643234244233, 0.2737426204589652,
0.26669628035356446, 0.2276260951060979, 0.4223655008580218, 0.027873028075357003, 0.17196955615765083,
0.017980857890688814, 0.050607160956566206, 0.044429400929868156, 0.18874737514738052, 0.31422705064607087,
0.5444839602597814, 0.05027499044940875, 0.003315788797809469, 0.1537148566971587, 0.10114216554594865,
0.13635218761558557, 0.41174295781809156, 0.4074089769445181, 0.11569059851647208, 0.14985736015868156,
0.07397005841209142, 0.007663941948522801, 0.07999858703364482, 0.06295414309536043, 0.2487423912626219,
0.03458476113549236, 0.017312197894786555, 0.21045261426729317, 0.23763220199236243, 0.2116859886492399,
0.02118213436642303, 0.04140104846465519, 0.10367493959007665, 0.12952939520691886, 0.140505906147713,
0.01348612362495972, 0.12517953000742196, 0.14244011092042344, 0.0384578151744934, 0.00634095360359937,
0.07803767798935572, 0.06970529994848448, 0.10228466060303405, 0.3051899611557444, 0.062150482881933,
0.0065382954154135494, 0.009424330109220141, 0.03188115695833248, 0.011223186509518356, 0.015001337075836948,
0.07727610830186225, 0.09123737110972113, 0.05378146145507942, 0.032080936221016355, 0.013005261168039278,
0.0067686667204196185, 0.0756393953597405, 0.17956645057219592, 0.22701197662726272, 0.20355754790026015,
0.15631117224350463, 0.10593764397306595, 0.0047847226453815285, 0.0638993032013592, 0.08546180389423226,
0.12976214258349494, 0.12429206390873306, 0.1285744233532176, 0.12426964260498206, 0.13302356176368618,
0.13273971613177535, 0.09284369666325692, 0.04124562564965308, 0.002633710957030138, 0.09476101351379539,
0.1558685064992605, 0.1499793880594929, 0.019494898289559193, 0.14810200602444057, 0.23887561647576377,
0.26771698490309687, 0.3018874654512851, 0.29178073209035643, 0.1874457499413155, 0.07375441810394558,
0.07301870789582512, 0.17776681123457982, 0.28579229899682496, 0.14689366710743373, 0.10788972408119864,
0.038055618685409985, 0.011802565005370635, 0.03080152598226531, 0.01500384210035854, 0.07601088135163611,
0.026711832633132288, 0.004291465524721346, 0.035435048757257316, 0.020919957886727014, 0.029174283919515195,
0.04807071671565217, 0.14562891666137803, 0.1952403357830954, 0.38536079232142795, 0.2729746819941042,
0.2664258549934131, 0.2266360539459262, 0.11563361586650463, 0.0011162957334283324, 0.07185610017223601,
0.11577703779355583, 0.05383108735926615, 0.09349886102115137, 0.015789448949836887, 0.018586464003345907,
0.02204899517590188, 0.021593664596323305, 0.1372380059142791, 0.1770706910625044, 0.21883208121993114,
0.14281860184746967], [0.10298693545512726, 0.18574514563169786, 0.195507732414973, 0.09011297567313128,
0.1116896862337328, 0.2577179034647478, 0.17266964084133385, 0.27743340531651417, 0.06802648372619427,
0.19525324397667887, 0.22797898365872774, 0.1788321852913077, 0.1512561498279942, 0.148127879767178,
0.3721952847669038, 0.04599747286769409, 0.05371506789799601, 0.21347336790848898, 0.14684114862402542,
0.1376466971633582‍3, 0.1520229854288104, 0.496008291241662, 0.20493971979724415, 0.32180437850314547,
0.09490334375050698, 0.11841053976679904, 0.06788115917525839, 0.22524350190689818, 0.06234087782073107,
0.05425258183113417, 0.04324547037125051, 0.253999956669033, 0.2899612805113186, 0.22949478259443187,
0.20393434402146277, 0.17462986539444691, 0.09473522719545227, 0.08354168908984927, 0.1328259352801816,
0.11632869111502665, 0.18267055717686512, 0.06817551389695826, 0.07852201440093881, 0.0621626691244895,
0.09816912073575153, 0.05718075605491252, 0.0538828029216676‍7, 0.16705623875885559, 0.053267713219159255,
0.05129786169712447, 0.00031361542457788495, 0.026937165940590816, 0.026841095045571372, 0.07798556499982129,
0.0844492687821303, 0.1043368338528215, 0.0709133895983045, 0.06088798369977662, 0.06870344819454638,
0.004537820199051604, 0.033594719314062826, 0.11709904237872151, 0.14525730731191605, 0.15247681494085413,
0.12633997811357495, 0.09374751321513745, 0.009246904871438389, 0.03344523907474006, 0.07364810907388314,
0.11471865402774047, 0.09602549894250972, 0.09265727928914506, 0.08360749617427997, 0.0827352634420996,
0.05917317547259891, 0.03003209383273778, 0.052026368124174, 0.01730076788157566, 0.012375362442756836,
0.038679270669184855, 0.0025674281048031385, 0.030850680781593885, 0.08422548580393131, 0.1337663837656032,
0.15996199510540146, 0.15616608995630826, 0.12742219856651255, 0.10666875602427688, 0.006046174435134545,
0.12490807659749407, 0.20156158258687287, 0.24924736274531709, 0.15361686074618502, 0.10762368130412141,
0.020655724292212626, 0.011342142299706894, 0.11760349559219807, 0.056923473740383326, 0.011234628833004481,
0.062282551813136654, 0.08728416871123885, 0.06613815034708803, 0.006054560741083938, 0.001695409585170473,
0.0732693275963205, 0.12535490131149082, 0.05594636655124604, 0.23549313255858753, 0.21242226672141948,
0.21653073554996133, 0.23353574066653865, 0.10798632420882552, 0.015466897210941315, 0.10972118821080099,
0.0707665466031597, 0.07712761076160884, 0.1174730676160727, 0.007602821444082549‍4, 0.060508086428806444,
0.0669124947945193, 0.1297579126716312, 0.09719445355114455, 0.023045229820228434, 0.12044264703048023,
0.18640233423269542], [0.29742094373392514, 0.008499307109214062, 0.07553022989809895, 0.3359223267938637,
0.2830618112358981, 0.17227573990000614, 0.012662410264857317, 0.05957962016512525, 0.023090013346231435,

0.0889710611089646, 0.09635021939128219, 0.13151690407642955, 0.24361154162480858, 0.39936564444704314,
0.2727394336286001, 0.05556563378447151, 0.019732431868127717, 0.08856170539434297, 0.12389160506533656,
0.25549845764437545, 0.4529435781295536, 0.27249092920789714, 0.361760840676965, 0.3076738976413794,
0.23220823496482, 0.09939128394947962, 0.10150858700935016, 0.08101831343198469, 0.256861839411061,
0.005874156929211517, 0.00992161438028563, 0.20080809832305171, 0.2891521473089744, 0.08051297942406448,
0.057353612227101454, 0.2554997926781431, 0.11035483396283016, 0.08224526135951586, 0.19346487267454032,
0.07392806670514031, 0.20286877892023722, 0.12163745160393659, 0.012445410066428233, 0.027586235332344285,
0.023159950744172684, 0.04443141971964565, 0.11079645238198102, 0.18575662561268272, 0.01102571389502701,
0.03279732075636875, 0.025046962650693762, 0.07777210875881294, 0.07175456029569688, 0.07206533252121712,
0.08779207715076494, 0.02569656973762865, 0.01940384608288871, 0.03573108649730881, 0.04584691477316825,
0.029632580498683468, 0.07986949775658436, 0.10863855415141885, 0.1464975953163708, 0.1621075531758037,
0.1260500118662784, 0.10600738146768802, 0.04943776744415442, 0.0528919949370813, 0.05182879358218028,
0.004532196589534832, 0.007683869799258769, 0.047089365526111926, 0.10631040921228999, 0.17730194047937745,
0.1698158307174527, 0.09436672547419545, 0.07870164930063031, 0.10883465780794399, 0.11351754665846696,
0.056296212012975614, 0.08106454294469344, 0.09948750779529657, 0.15037207607201944, 0.19062253585480393,
0.14540207117160878, 0.122606663475354, 0.12473352554451275, 0.09533386132246072, 0.056163133101888585,
0.037221090138863135, 0.09479463295223092, 0.13249601758719193, 0.013042038992721758, 0.01549008566394577,
0.014802487699664585, 0.034511772744963136, 0.007850130952845558, 0.01881804156973166, 0.06700153868712619,
0.03043580990594509, 0.08439979817113337, 0.0973833098742213, 0.09234889960039683, 0.0434341346767814,
0.05462226332703594, 0.17977753295972373, 0.17079836835876744, 0.29805146916188585, 0.1846424037137482,
0.16190917244694533, 0.23531590128914628, 0.15237980748935892, 0.07320010552529489, 0.04860745261364736,
0.011316782960832505, 0.0641679500092982, 0.02410400096865496, 0.09322190409610376, 0.10271113396488758,
0.11514097974471683, 0.09037977198681954, 0.11428993698206742, 0.20014204771606606, 0.269468465989644,
0.23880842533437166], [0.5655649935562048, 0.1714406222150879, 0.13966045933111665, 0.33634409457328285,
0.21879112617073107, 0.10896440607465695, 0.05990291717793807, 0.42477116808284304, 0.029238242346130035,
0.027920966377233666, 0.04728394278090114, 0.19413013009724772, 0.28205386035420377, 0.40003473239715454,
0.39362216182862797, 0.04134623726123393, 0.03449638561396614, 0.16147094607156756, 0.227360524883539,
0.08872493230607391, 0.569160685163172, 0.10723902090769433, 0.18287452295276574, 0.36927897136307725,
0.16140199968382019, 0.03464757352204002, 0.16971358347194218, 0.13493145043814217, 0.20148643058403506,
0.005338563796143699, 0.08972780741866938, 0.03707193603746275, 0.3604846205836836, 0.17708361895277314,
0.03141545507065068, 0.26884314273849585, 0.1119365549145316, 0.044183609710363565, 0.21526960681241145,
0.12161470270147039, 0.30007187279703385, 0.18936157596802258, 0.037431948222428534, 0.0594200787252179,
0.05383516514937356, 0.0034453302833364454, 0.015367580354129287, 0.17952352105921848, 0.06714021960053301,
0.06936532364742158, 0.024292445913250527, 0.05477632996693742, 0.024130686709195742, 0.09161250283192376,
0.02813944862331056, 0.022302893737459917, 0.01592694582097282, 0.010669275913345459, 0.0035870747027531946,
0.053285089591137244, 0.05412607800955586, 0.11602346501236946, 0.14783223435336007, 0.20852139401087486,
0.18111041912095927, 0.14431549426932172, 0.0647189679703095, 0.03728043596717788, 0.062245437058125075,
0.10001712721794108, 0.07581066244308401, 0.07134128887918467, 0.06642235040870036, 0.08404607724628475,
0.023444843786363627, 0.011769001080915562, 0.04484099880303352, 0.0022026099495033695, 0.09815737030609253,
0.032969949706305665, 0.021414094389290605, 0.009747947436817657, 0.049999975059246596, 0.09112676929714497,
0.08919368234173183, 0.10036971190780705, 0.15230693692112385, 0.12145599279911155, 0.07119442204578635,
0.08893610289610722, 0.15007965138292637, 0.23895849081017576, 0.07126228696128593, 0.03761345544838987,
0.020270498377192542, 0.07331027578854545, 0.04660130620617257, 0.09581970941257045, 0.05742966616102667,
0.025486088486845644, 0.02051748213462214, 0.023761259476465554, 0.10951553411218926, 0.2500883373579288,
0.14445621468712708, 0.04409483926327595, 0.15047357660405497, 0.45978383184090893, 0.3938219484931491,
0.3167427461961294, 0.3116177998523038, 0.02776099483282675, 0.07475711947014292, 0.20516020174844077,
0.18446429276378554, 0.1158484514688028, 0.11343886089062312, 0.013541865332448078, 0.04164937501602452,
0.13317631250829842, 0.024429411510780024, 0.009018727322862605, 0.08668587869353972, 0.23288616815476637,
0.1549335615585084], [0.24176983487514334, 0.14853369787419338, 0.1395195534716059, 0.37876788017701635,
0.3441035872681215, 0.11156602760249387, 0.16220894471336053, 0.03149947318250505, 0.348506479691959,
0.00721752887983351, 0.0707365228880556, 0.33292757046802135, 0.3886031787953216, 0.41975596008008464,
0.25674143496622315, 0.08072146270319719, 0.16510675908338515, 0.3384391563103737, 0.28169947197188244,
0.08546587346839644, 0.24205416405790597, 0.2066117258322293, 0.37494954814806236, 0.3513654235184157,
0.11269081675188006, 0.28335111049095557, 0.03790407469004339, 0.08695308933761059, 0.24846523855240352,
0.20052824637123134, 0.009578893456956684, 0.18223383665561335, 0.4040354784909404, 0.08204013473403496,
0.06520992036737368, 0.23680826311264527, 0.06184181476075684, 0.13915498244996666, 0.32906373781557885,
0.19437825729709513, 0.13823137213916356, 0.1450625108971492, 0.04709037611355009, 0.21078105780218886,
0.08597443353248255, 0.1176440061037537, 0.107183072395221, 0.2019412515545689, 0.035405289915670074,
0.08274756267444472, 0.09035599523637534, 0.04604717601833025, 0.052944834063527534, 0.08410383281987087,
0.03962353280870317, 0.02886197896819333, 0.0403991287293151, 0.08756507143595342, 0.06112297730511981,
0.009926784211939826, 0.015987304079707346, 0.13670706559921086, 0.18150406160917346, 0.18835999720438962,
0.17432820150381212, 0.1435667208587957, 0.07082131480194243, 0.04485169538291272, 0.016770449222438876,
0.03263299563168094, 0.0008697022044521853, 0.016332725851264995, 0.1218416446536436, 0.1994826025631603,
0.15367147751413388, 0.15856081527298277, 0.19321992843364216, 0.11643061343405575, 0.05323689013910385,
0.0027682223479106854, 0.007408731705755834, 0.018058864349950222, 0.024544631929750985, 0.061623683744627,
0.050228971910092804, 0.06763878391804137, 0.12805360669744745, 0.07178315231999753, 0.01348621330294127,
0.15496257953819562, 0.21157974862800272, 0.2527253725815564, 0.1469982759449111, 0.0666495761168578,
0.030487452423367867, 0.02322696880479262, 0.11666466834645742, 0.09253364016308574, 0.05860166771262239,
0.16412679687649634, 0.13758209554527373, 0.09835520157942246, 0.018290633011469038, 0.048160615850489016,
0.05555227377242027, 0.20110446494914422, 0.22319009236112713, 0.29914896785316347, 0.15411081992634615,
0.11338557763279651, 0.11689808869015503, 0.02089148307489627, 0.17551653708917053, 0.14819149782407876,
0.1348420967851232, 0.11576369630659512, 0.12914198882112657, 0.04054706217963935, 0.014227941950975435,
0.03049104959617112, 0.036482949932100814, 0.03265848539270407, 0.02441766644908042, 0.0893447969743226,
0.11856442111020106], [0.25432763905150924, 0.12527103273904783, 0.05468324655161736, 0.17605834717744806,
0.09758882778268942, 0.13226380003414492, 0.08167599471784207, 0.059199790106022485, 0.04769590635961221,
0.015510243942355355, 0.19543098084290988, 0.07157665444724273, 0.3026465929667456, 0.15771078671900102,
0.1517108924556638, 0.03941712421679017, 0.03625160012110797, 0.027973801137719988, 0.15635564778920752,
0.137341248544169, 0.42848572453106915, 0.23763083936042947, 0.5157843363910543, 0.45110823903561637,
0.2517038273472497, 0.06467070449007332, 0.0005785812126051439, 0.0753390279537193, 0.2054943380631094,
0.08505906798407353, 0.026404725967084836, 0.1956863232701136, 0.3086489618157298, 0.06534407395921692,
0.05725155801949708, 0.21115307017378593, 0.2208008389405845, 0.04544532388189482, 0.0800224984700047,
0.0021253265196061774, 0.0937145588025646, 0.19394342956696448, 0.07790276100391297, 0.12328291825635612,
0.011140495924530773, 0.03344375208656902, 0.06788250107729486, 0.13322246374343977, 0.045441417361167816,

0.049200845004130814, 0.012960001930999422, 0.06151679955262011, 0.05555438140652595, 0.07676123311233644,
0.09487282629879273, 0.04284540350509582, 0.00603899577179375, 0.036956836447626096, 0.05264329055108919,
0.02321836332687756, 0.009567880140468738, 0.08713927436473452, 0.14275037261382945, 0.12633455532991275,
0.09048122512997873, 0.05672566345558643, 0.021630003355357273, 0.014121926402247641, 0.02343612116910511,
0.008087583395189474, 0.036819052284947376, 0.0028563554684280217, 0.06340318863832946, 0.11142083266391216,
0.05874593205440862, 0.04710679605264561, 0.05341117275318702, 0.06718276284040921, 0.19089392328443808,
0.19075555600392816, 0.16038289008547604, 0.10921480857033587, 0.18943152052367007, 0.2454391411957137,
0.22291217431572166, 0.1844664471113714, 0.16674694973922458, 0.14765538669758352, 0.036491868324653766,
0.08275313810731758, 0.1686467015959084, 0.22447941584915165, 0.08979077668376423, 0.04021775947018716,
0.0047620990267466376, 0.010498611802101005, 0.018950345620450212, 0.06488060187165569, 0.04731324932440685,
0.022275391754538947, 0.04354477201555599, 0.02950021623623444, 0.05527113189065942, 0.1038624573234498,
0.10512466573703842, 0.1360774469958596, 0.2212648534838901, 0.4264145506936073, 0.3027096297537257,
0.2568050383328624, 0.33922216825834456, 0.1918875142356138, 0.045484601751099404, 0.04889222860743564,
0.10858183786200659, 0.0712485250366114, 0.07508381235406224, 0.024209591891023865, 0.012265397482525751,
0.04124096795558889, 0.06524098532710605, 0.07059878154307726, 0.18219011972342178, 0.2325888604023124,
0.19654146666936195], [0.3657343058126263, 0.06565577686415829, 0.0015546155913509245, 0.14922559325520102,
0.27807903880810975, 0.27038382032384184, 0.18257593802661448, 0.09808066249203533, 0.17890494456583908,
0.13759610969169875, 0.15536282830166162, 0.2631815076833804, 0.15694540292958, 0.5379504000806083,
0.35115714956471744, 0.07560465241711851, 0.08425052658133472, 0.06364107441610156, 0.010183861422830908,
0.24608021519799572, 0.46560337039320265, 0.3427618622877664, 0.3383929482969315, 0.21298191584814252,
0.038255989032144895, 0.1389041653929605, 0.09853373686724361, 0.06660225103085012, 0.12126923868406572,
0.052156330682640674, 0.010596664142971819, 0.08685200893003843, 0.23814578906423953, 0.062208996629216105,
0.021388603806563027, 0.19051016134914353, 0.21527099183382187, 0.09217967073501342, 0.2078657049377866,
0.04411846497269794, 0.22093950109509344, 0.20118378593450864, 0.017070307707249228, 0.09151668700778084,
0.023897457446102503, 0.013833521606997858, 0.03685621136943804, 0.1811717226999468, 0.048910751796586986,
0.061518184017562164, 0.13480914447417822, 0.16335727713475687, 0.06662720499654379, 0.002423796969390138,
0.04427017464727773, 0.0925850633915065, 0.033579830970657316, 0.011408441249102225, 0.05902431000041901,
0.05591873946805743, 0.012200198254417233, 0.09495440629666066, 0.17994602434413026, 0.20378748555216933,
0.18724704807466686, 0.17419958750583475, 0.125277889027267, 0.0827435550136447, 0.06449481731218853,
0.04050882210443501, 0.025346008103168675, 0.024085796201385706, 0.1288711049324171, 0.2067971239000694,
0.1692160821625897, 0.12168825046199724, 0.17571433265307093, 0.1643005997576389, 0.15085965429945047,
0.15642574594664208, 0.1465692728586429, 0.17667754084429693, 0.19745211786935252, 0.23450769644150243,
0.22442162416643432, 0.16810113169002092, 0.19612359465843057, 0.17455176808005335, 0.11787775201522124,
0.014158501972965183, 0.09218530627165802, 0.14544920176685147, 0.050872966762275165, 0.04107021203659448,
0.009537058142291454, 0.01452417355815831, 0.013157895005710586, 0.049463140169333425, 0.022285211808513457,
0.0736338631094657, 0.14369780581365266, 0.13679484589637544, 0.1385967759587702, 0.024519205894722385,
0.05285231894139239, 0.1362157241174588, 0.18175760432917204, 0.3934729205459303, 0.32290492903015844,
0.31638327554790435, 0.3572194189011921, 0.23133997777823026, 0.13656807043150015, 0.04223441113362223,
0.04755022925257813, 0.024224746180777378, 0.010435314443089103, 0.014964024623556024, 0.03616925993477595,
0.025896219941137954, 0.023646008698016568, 0.037456122665368685, 0.04881582018057474, 0.17081148127773582,
0.23057472241431885], [0.40643420496954297, 0.06827011426216945, 0.10759888204091081, 0.30400534871208384,
0.13364206982267623, 0.29841049547151005, 0.11626216093378758, 0.4966889579827883, 0.06992542860840244,
0.18719093747927945, 0.203633124517894, 0.07793984203347588, 0.24987916060681856, 0.35331463210459646,
0.4309834456171245, 0.10534403246795022, 0.12925885861195732, 0.18947432881807402, 0.12856247082363803,
0.218236514575856, 0.2635520356313481, 0.34889228262136335, 0.1898111884736423, 0.27701615317903433,
0.135403321771204, 0.045968090742771944, 0.17475613996893477, 0.04546460384865761, 0.1157277159693237,
0.010211512667315244, 0.14760808357941801, 0.10817165013057704, 0.1935396402408531, 0.008551785133490983,
0.08061887221268835, 0.20387332916852907, 0.038845906138847835, 0.027747687477713452, 0.058035676096456476,
0.03394338091000693, 0.1964104742098073, 0.13589109537820224, 0.05373612740496773, 0.00039616826584443795,
0.01876912228927724, 0.02200729686770851, 0.05600754253248005, 0.2284956468893883, 0.07028493667467896,
0.05834731860007997, 0.11258342314613211, 0.18349953428644009, 0.11687830132632823, 0.0917815641230425,
0.13233308293381169, 0.07815494988335864, 0.03669779679200785, 0.023759580640237525, 0.026807253111576612,
0.024941350446421814, 0.0673121036149261, 0.12553582994799894, 0.1324829886755639, 0.14056851945179863,
0.08013002875016695, 0.0822361751938127, 0.012777797770969504, 0.07430234889931356, 0.10974279201953985,
0.1534358213527395, 0.15788070714052682, 0.16233741123721435, 0.08516055766220501, 0.025847361794919085,
0.0011405698824656198, 0.022346378689195076, 0.02306577927289772, 0.0696946960270604, 0.14326232666153946,
0.15800607418144108, 0.1702563911937426, 0.19758526165723375, 0.23155341200277227, 0.2636328768677696,
0.22991276652350828, 0.2406468699441157, 0.22707313348019165, 0.12528135989457231, 0.09468653587040597,
0.028677240467084964, 0.09937580534070822, 0.15065125743457505, 0.013894578236045466, 0.03955142146933985,
0.03557051678983855, 0.05549520120827765, 0.04468529288441058, 0.03228776429877513, 0.0407765976242995,
0.020955610042826282, 0.013154379162079184, 0.08576792819734698, 0.10447647422403736, 0.03328823951783286,
0.12561732459917369, 0.2723166135502273, 0.2570243781722319, 0.3895050090597824, 0.25640711918835263,
0.18025865768570293, 0.24868609064273306, 0.1715297594683967, 0.07323686471236335, 0.05832226820922972,
0.06503493437291723, 0.09198633148236987, 0.08154959015864487, 0.08883198535708593, 0.11713198352168792,
0.10610399422751822, 0.11770680419823525, 0.09194288406376601, 0.2000159794813008, 0.23508759366994633,
0.1881792801754761], [0.41813427697790356, 0.10638456813323585, 0.21233155058007877, 0.3815855096141117,
0.2020290251247387, 0.33230563715232775, 0.0704836107966313, 0.058276875666528956, 0.04948411244498648,
0.15308035438845813, 0.06319579311692879, 0.1638330866765779, 0.39335721725748796, 0.5379736995378817,
0.4265588058504394, 0.019111985358470487, 0.05801201404220585, 0.17805144607367915, 0.10698528302750178,
0.27397930221597905, 0.4158331428032717, 0.392796557708995, 0.2662028195728244, 0.33235857928729823,
0.20952443363329215, 0.06307107301596739, 0.001580009912601363, 0.001858149221399557, 0.16285470742364175,
0.03559008957108974, 0.019173123861526666, 0.1470538474054319, 0.3778987613502669, 0.19157490570909938,
0.0691187180764134, 0.19288994087669892, 0.1383241939692935, 0.032654556895098115, 0.10309257742555501,
0.10326104873315188, 0.2774575957586805, 0.19841232585563157, 0.008236404969308095, 0.05527727708335367,
0.01695008098630092, 0.00735668867183198, 0.00207311443581612, 0.1635852273764418, 0.07973063429688605,
0.08903620329694818, 0.012771129733567014, 0.051074182046692014, 0.04168687602223443, 0.05487037556256279,
0.07529775618040954, 0.05574239519823908, 0.0809456252193655, 0.06814469611292641, 0.05149131377159829,
0.0019555283174013272, 0.04093916769852522, 0.12912156948548756, 0.16135892199446655, 0.1815800722281255,
0.14658178182049922, 0.0967493118528587, 0.027183454369143118, 0.06839211579517622, 0.10438303357473579,
0.1509727068599618, 0.16920859381307693, 0.15483699592306321, 0.11669812657682842, 0.11050160376521656,
0.09121985301129659, 0.07102087292580278, 0.05759348050532373, 0.00812738644717887, 0.05064270375350985,
0.12175927489801168, 0.1047902078847257, 0.006911381586145886, 0.1342211444983509, 0.218328601027318,
0.1976890148964487, 0.15670545717338683, 0.12034835601702891, 0.07913513264346017, 0.02713432804626626,

0.11627990305892155, 0.1780843664387155, 0.2795008693508003, 0.1861231868578127, 0.16089140648406977,
0.03517249137714237, 0.03170195488519582, 0.08285030139079311, 0.06785227834135332, 0.018019208052621683,
0.14232912972146528, 0.18431053041380796, 0.16867019568943933, 0.10571216463522087, 0.00330848990943375,
0.09411176956469847, 0.030336535020468843, 0.06365684500979513, 0.1994841914996558, 0.17972826051821608,
0.24034555078608938, 0.3056189446897869, 0.2453916379166239, 0.06432141632478258, 0.020814676775993107,
0.03267830097420454, 0.02718728932629231, 0.09554725464892344, 0.047393302358948695, 0.06995324759561414,
0.031521396297207384, 0.04359355093652537, 0.13026233857498665, 0.14057396506730832, 0.130854601466148,
0.0015244246322316313]], "wl": [908.1, 914.294, 920.489, 926.683, 932.877, 939.072, 945.266, 951.46, 957.655,
963.849, 970.044, 976.238, 982.432, 988.627, 994.821, 1001.015, 1007.21, 1013.404, 1019.598, 1025.793,
1031.987, 1038.181, 1044.376, 1050.57, 1056.764, 1062.959, 1069.153, 1075.348, 1081.542, 1087.736, 1093.931,
1100.125, 1106.319, 1112.514, 1118.708, 1124.902, 1131.097, 1137.291, 1143.485, 1149.68, 1155.874, 1162.069,
1168.263, 1174.457, 1180.652, 1186.846, 1193.04, 1199.235, 1205.429, 1211.623, 1217.818, 1224.012, 1230.206,
1236.401, 1242.595, 1248.789, 1254.984, 1261.178, 1267.373, 1273.567, 1279.761, 1285.956, 1292.15, 1298.344,
1304.539, 1310.733, 1316.927, 1323.122, 1329.316, 1335.51, 1341.705, 1347.899, 1354.094, 1360.288, 1366.482,
1372.677, 1378.871, 1385.065, 1391.26, 1397.454, 1403.648, 1409.843, 1416.037, 1422.231, 1428.426, 1434.62,
1440.814, 1447.009, 1453.203, 1459.398, 1465.592, 1471.786, 1477.981, 1484.175, 1490.369, 1496.564, 1502.758,
1508.952, 1515.147, 1521.341, 1527.535, 1533.73, 1539.924, 1546.119, 1552.313, 1558.507, 1564.702, 1570.896,
1577.09, 1583.285, 1589.479, 1595.673, 1601.868, 1608.062, 1614.256, 1620.451, 1626.645, 1632.839, 1639.034,
1645.228, 1651.423, 1657.617, 1663.811, 1670.006, 1676.2]}, "occlusion": {"base_f1": 0.7766933066847855,
"base_auc": 0.8478519209637033, "delta": {"top5": [-0.0005453120014990986, -0.004585163172284856], "top10":
[-0.015565614025676022, -0.011140773310169938], "top20": [-0.00924458537263051, -0.01634755642303365],
"rand5": [0.002723984170910909, -0.00015021326513864341], "rand10": [0.0026779487032012605,
-0.001615620732906664], "rand20": [0.0014235076059639917, -0.0016261249575864056], "contig5":
[-0.00037069255009891666, 0.0023403804292678743], "contig10": [-0.008650060745846733, -0.003141427701874866],
"contig20": [-0.02774203097493244, -0.016827280685573376]}}}

#### S10. results/cnn_nested.json

{
 "f1": [
  0.7708333333333333,
  0.6838978015448604,
  0.7270037839697282,
  0.6825031928480204,
  0.7886728655959425
 ],
 "acc": [
  0.8181818181818182,
  0.7593984962406015,
  0.7578125,
  0.7079646017699115,
  0.8076923076923077
 ],
 "bacc": [
  0.7708333333333333,
  0.6806122448979592,
  0.7315671432576395,
  0.6800804828973843,
  0.7872670807453416
 ],
 "auc": [
  0.860207100591716,
  0.8119533527696793,
  0.8474908887019905,
  0.7250167672702883,
  0.8242753623188407
 ],
 "chosen": [
  4,
  4,
  4,
  4,
  4
 ],
 "oof": [
  0.912235677242279,
  0.5352888107299805,
  0.42406702041625977,
  0.42288562655448914,
  0.8482723832130432,
  0.49874114990234375,
  0.16946901381015778,
  0.12643076479434967,
  0.22806574404239655,
  0.49385127425193787,
  0.22340726852416992,
  0.24513137340545654,
  0.21661055088043213,
  0.14815300703048706,
  0.6913468241691589,
  0.9021247029304504,

0.9608449339866638,
0.5679032802581787,
0.6854732632637024,
0.9175105094909668,
0.6657916903495789,
0.18615387380123138,
0.7904355525970459,
0.881935179233551,
0.4373694956302643,
0.7706968188285828,
0.7395179271697998,
0.2248474359512329,
0.1500377506017685,
0.7064242362976074,
0.9193460941314697,
0.894326388835907,
0.6752803325653076,
0.9062268137931824,
0.3080817759037018,
0.9487812519073486,
0.7527703642845154,
0.390617698431015,
0.2726147472858429,
0.5976419448852539,
0.19730181992053986,
0.17781014740467072,
0.24113018810749054,
0.6337423324584961,
0.37589481472969055,
0.8589146137237549,
0.5828068852424622,
0.7392234206199646,
0.4078977108001709,
0.5547127723693848,
0.3410223424434662,
0.4336985647678375,
0.26852285861968994,
0.42217543721199036,
0.2575649321079254,
0.6875438094139099,
0.2546204626560211,
0.14936529099941254,
0.20780962705612183,
0.22203849256038666,
0.13100211322307587,
0.26212820410728455,
0.2918812334537506,
0.2876181900501251,
0.197454035282135,
0.18829528987407684,
0.34821367263793945,
0.4944651424884796,
0.8670843243598938,
0.33505842089653015,
0.4351203441619873,
0.4636898934841156,
0.6470350623130798,
0.9610324501991272,
0.9488540291786194,
0.7300184369087219,
0.23399393260478973,
0.5508382320404053,
0.14365246891975403,
0.2121496945619583,
0.2947409152984619,
0.15639148652553558,
0.2829073965549469,
0.25493618845939636,
0.3004200756549835,
0.15631155669689178,
0.22293721139431,
0.13589543104171753,
0.47168049216270447,
0.2911128103733063,
0.4040151536464691,
0.3981952369213104,
0.25353220105171204,
0.14467547833919525,
0.17195479571819305,
0.7427659034729004,
0.37582388520240784,
0.2286853790283203,
0.228105366230011,

0.4234970808029175,
0.24189573526382446,
0.3289047181606293,
0.5604727268218994,
0.12951795756816864,
0.15669448673725128,
0.9756783843040466,
0.8184430003166199,
0.8474096655845642,
0.20738613605499268,
0.4296388626098633,
0.1171373799443245,
0.1403571367263794,
0.09918294101953506,
0.11798278242349625,
0.38688352704048157,
0.24459223449230194,
0.4651125967502594,
0.44810542464256287,
0.3460364043712616,
0.34019652009010315,
0.7370522022247314,
0.21231569349765778,
0.1719709187746048,
0.2977849543094635,
0.1223016083240509,
0.13541264832019806,
0.4223537743091583,
0.18372468650341034,
0.2188168615102768,
0.13040240108966827,
0.10695245116949081,
0.14060094952583313,
0.19879911839962006,
0.10834074020385742,
0.11710634082555771,
0.2425532191991806,
0.48204776644706726,
0.17924325168132782,
0.4433024823665619,
0.17234401404857635,
0.39820849895477295,
0.08714141696691513,
0.1278916895389557,
0.4373248517513275,
0.16009168326854706,
0.8791257739067078,
0.8406462669372559,
0.29759886860847473,
0.341080904006958,
0.1985132247209549,
0.28816041350364685,
0.35450586676597595,
0.6157390475273132,
0.15114560723304749,
0.12657764554023743,
0.1574731320142746,
0.1687072515487671,
0.15460552275180817,
0.22569775581359863,
0.8857032656669617,
0.3716946542263031,
0.3676643371582031,
0.7827057838439941,
0.9187837243080139,
0.5734401345252991,
0.24705272912979126,
0.8104484677314758,
0.19690650701522827,
0.7395266890525818,
0.9242346286773682,
0.2013085037469864,
0.2373397797346115,
0.10864558070898056,
0.26465728878974915,
0.1722126603126526,
0.608863353729248,
0.11305954307317734,
0.10575159639120102,
0.097658671438694,
0.237263485789299,
0.982361376285553,
0.9919511675834656,

0.5037593245506287,
0.1625005304813385,
0.6223734021186829,
0.3325887620449066,
0.21885544061660767,
0.7752450108528137,
0.6837332844734192,
0.298445463180542,
0.671741783618927,
0.8058114051818848,
0.6495905518531799,
0.8031098246574402,
0.6361735463142395,
0.16988319158554077,
0.1683114767074585,
0.4159318208694458,
0.36635562777519226,
0.4665358066558838,
0.38294515013694763,
0.8697472214698792,
0.2415706366300583,
0.3726697862148285,
0.6634839177131653,
0.7039268612861633,
0.21625065803527832,
0.32415592670440674,
0.8872676491737366,
0.6620327830314636,
0.6905840039253235,
0.6055311560630798,
0.4864296019077301,
0.38807153701782227,
0.27078017592430115,
0.8418116569519043,
0.5292231440544128,
0.18677622079849243,
0.3806075155735016,
0.19330255687236786,
0.283549427986145,
0.17941009998321533,
0.222410187125206,
0.2847503125667572,
0.7276201248168945,
0.3014388382434845,
0.34394264221191406,
0.26463642716407776,
0.864586353302002,
0.14919202029705048,
0.3429912030696869,
0.12567120790481567,
0.17767637968063354,
0.22668540477752686,
0.326905220746994,
0.506157398223877,
0.5154132843017578,
0.5439662933349609,
0.32453635334968567,
0.8070268630981445,
0.6682340502738953,
0.8952894806861877,
0.6566715836524963,
0.30787524580955505,
0.6489213109016418,
0.11896568536758423,
0.22299480438232422,
0.19724102318286896,
0.25550779700279236,
0.5831210017204285,
0.5861282348632812,
0.6051906943321228,
0.6930370926856995,
0.502907931804657,
0.4172346591949463,
0.36030682921409607,
0.41936931014060974,
0.6746469140052795,
0.44294849038124084,
0.6079841256141663,
0.5375500321388245,
0.6879844665527344,
0.9133524298667908,
0.9946210384368896,
0.38522812724113464,

0.23940761387348175,
0.36899563670158386,
0.8636379837989807,
0.45316410064697266,
0.77582186460495,
0.3101705014705658,
0.4219491481781006,
0.23632657527923584,
0.23355723917484283,
0.21650023758411407,
0.09729480743408203,
0.13274240493774414,
0.38717666268348694,
0.2538749873638153,
0.29069098830223083,
0.2615428566932678,
0.7384927868843079,
0.44321396946907043,
0.3571355640888214,
0.274640828371048,
0.3061624765396118,
0.16666175425052643,
0.15006397664546967,
0.41592851281166077,
0.3157552480697632,
0.4381936490535736,
0.5748047232627869,
0.11425558477640152,
0.5116636753082275,
0.3282740116119385,
0.27831050753593445,
0.28304150700569153,
0.22788113355636597,
0.3761732280254364,
0.14759491384029388,
0.19212917983531952,
0.3595069944858551,
0.11333045363426208,
0.19297318160533905,
0.23242665827274323,
0.19193415343761444,
0.23157711327075958,
0.22874432802200317,
0.19442182779312134,
0.5710344314575195,
0.24086345732212067,
0.8429651260375977,
0.26142773032188416,
0.19488467276096344,
0.5596016049385071,
0.29264771938323975,
0.8946542739868164,
0.34955522418022156,
0.665142297744751,
0.8807886242866516,
0.32620880007743835,
0.988658607006073,
0.49711874127388,
0.3051191568374634,
0.1734507530927658,
0.29858505725860596,
0.3275974988937378,
0.866740882396698,
0.5948695540428162,
0.3034181296825409,
0.5329996943473816,
0.40152859687805176,
0.24358844757080078,
0.405716210603714,
0.184679314494133,
0.17465849220752716,
0.1815280169248581,
0.17071974277496338,
0.29512637853622437,
0.1528937816619873,
0.11109455674886703,
0.5540714859962463,
0.3402808904647827,
0.3152247667312622,
0.13000164926052094,
0.15946908295154572,
0.6016953587532043,
0.24458055198192596,

0.6085826754570007,
0.17060492932796478,
0.9006900191307068,
0.8658069968223572,
0.26468077301979065,
0.18790477514266968,
0.43344947695732117,
0.19848763942718506,
0.3955979645252228,
0.38152870535850525,
0.5431433320045471,
0.8316020369529724,
0.692549467086792,
0.6280663013458252,
0.3189980685710907,
0.37140461802482605,
0.29954853653907776,
0.16356579959392548,
0.2278807908296585,
0.247671440243721,
0.19885103404521942,
0.18797121942043304,
0.26851823925971985,
0.14846830070018768,
0.6654686331748962,
0.7438123822212219,
0.2486376017332077,
0.5880922675132751,
0.8489429354667664,
0.8899384140968323,
0.597862184047699,
0.5578811764717102,
0.14777980744838715,
0.15650510787963867,
0.1475447416305542,
0.1772444248199463,
0.10147512704133987,
0.11938204616308212,
0.15581883490085602,
0.8599610328674316,
0.4041874408721924,
0.315602570772171,
0.3219027519226074,
0.5524126887321472,
0.8710281252861023,
0.7748315930366516,
0.298008531332016,
0.30032864212989807,
0.34507179260253906,
0.3781697452068329,
0.9363909363746643,
0.2760910987854004,
0.3344489336013794,
0.49549493193626404,
0.232469379901886,
0.20187337696552277,
0.23844152688980103,
0.4799772799015045,
0.25644350051879883,
0.216167613863945,
0.9756185412406921,
0.8376967310905457,
0.6689596176147461,
0.28861260414123535,
0.41508230566978455,
0.33263257145881653,
0.2631232440471649,
0.31128501892089844,
0.2492796927690506,
0.43298494815826416,
0.8810500502586365,
0.3251393139362335,
0.4706767797470093,
0.6289206147193909,
0.20271170139312744,
0.36920633912086487,
0.15339933335781097,
0.7380859851837158,
0.40453001856803894,
0.2547636926174164,
0.3279312551021576,
0.4129045903682709,
0.351059228181839,

0.7731264233589172,
0.984722375869751,
0.38027188181877136,
0.6580765247344971,
0.7681059837341309,
0.1731688380241394,
0.8677369952201843,
0.6000593304634094,
0.8085033893585205,
0.693239688873291,
0.9381997585296631,
0.17113347351551056,
0.2549435794353485,
0.436025470495224,
0.30188438296318054,
0.5874067544937134,
0.8405582308769226,
0.4321999251842499,
0.6591577529907227,
0.39695656299591064,
0.7361722588539124,
0.8098778128623962,
0.2514493763446808,
0.287008136510849,
0.733496367931366,
0.49839329719543457,
0.5485402941703796,
0.1555848866701126,
0.4450456202030182,
0.46154436469078064,
0.6754866242408752,
0.23974181711673737,
0.4127570688724518,
0.18886272609233856,
0.19787681102752686,
0.2558024227619171,
0.4827265739440918,
0.9872972369194031,
0.3617742359638214,
0.5162996649742126,
0.9137219786643982,
0.35277533531188965,
0.9890778064727783,
0.971430778503418,
0.9772723317146301,
0.9710444808006287,
0.2554206848144531,
0.21847926080226898,
0.22769804298877716,
0.2901149094104767,
0.38949427008628845,
0.4300963580608368,
0.16174912452697754,
0.464021235704422,
0.22780804336071014,
0.47952982783317566,
0.31835734844207764,
0.7184333205223083,
0.13066482543945312,
0.15334896743297577,
0.28427132964134216,
0.2917316257953644,
0.26901015639305115,
0.30763310194015503,
0.268125981092453,
0.22223693132400513,
0.252068430185318,
0.6042850017547607,
0.4158106744289398,
0.5450453162193298,
0.6486290693283081,
0.291867733001709,
0.6641848087310791,
0.7492510676383972,
0.7910275459289551,
0.3434452712535858,
0.22435329854488373,
0.4490004777908325,
0.34390804171562195,
0.4534864127635956,
0.5052487254142761,
0.19408635795116425,
0.47239336371421814,

0.3252197206020355,
0.17692796885967255,
0.18767833709716797,
0.18518026173114777,
0.8671393990516663,
0.2310633659362793,
0.3954215347766876,
0.38217464089393616,
0.09168460220098495,
0.11389404535293579,
0.3364146649837494,
0.38176634907722473,
0.31279781460762024,
0.31318360567092896,
0.3090771734714508,
0.3312698304653168,
0.3623119294643402,
0.2842363119125366,
0.3973037004470825,
0.280169278383255,
0.22532467544078827,
0.15772952139377594,
0.1825304627418518,
0.3519998788833618,
0.29088321328163147,
0.355555921792984,
0.2901231348514557,
0.24229466915130615,
0.14823921024799347,
0.14098437130451202,
0.11897870898246765,
0.12061405181884766,
0.2359909862279892,
0.24134480953216553,
0.8911067843437195,
0.3044276535511017,
0.6721816062927246,
0.47871705889701843,
0.17048488557338715,
0.32830870151519775,
0.24694804847240448,
0.9025009274482727,
0.563519299030304,
0.4949101507663727,
0.27638497948646545,
0.30443689227104187,
0.2518887221813202,
0.23067934811115265,
0.7845829129219055,
0.4129364490509033,
0.17107422649860382,
0.344431608915329,
0.6868411898612976,
0.3962336480617523,
0.3609929382801056,
0.8816727995872498,
0.3877536356449127,
0.21364767849445343,
0.18196403980255127,
0.16322951018810272,
0.505506694316864,
0.7624855041503906,
0.6870854496955872,
0.6915146708488464,
0.15038001537322998,
0.29878532886505127,
0.1849898248910904,
0.43934524059295654,
0.386318564414978,
0.9904179573059082,
0.5646533370018005,
0.20388595759868622,
0.301387220621109,
0.8903807997703552,
0.480864018201828,
0.49407902359962463,
0.9263588786125183,
0.48745131492614746,
0.47877785563468933,
0.32692909240722656,
0.873124897480011,
0.5438991189002991,
0.6425924897193909,

  0.4866810739040375,
  0.3939187824726105,
  0.6534101963043213,
  0.4002895653247833,
  0.2787160575389862,
  0.21072185039520264,
  0.21989955008029938,
  0.15701164305210114,
  0.12369212508201599,
  0.12359961122274399,
  0.12670792639255524,
  0.10426318645477295,
  0.14091989398002625,
  0.0971221849322319,
  0.4550880491733551,
  0.3908369839191437,
  0.12704579532146454,
  0.3252996504306793,
  0.8525951504707336,
  0.13613809645175934,
  0.34033170342445374,
  0.5891807675361633,
  0.2902188003063202,
  0.5640990138053894,
  0.9234650731086731,
  0.8885905146598816,
  0.8074442744255066,
  0.3217034637928009,
  0.3319821059703827,
  0.9452984929084778,
  0.10176157206296921,
  0.36579540371894836,
  0.36137518286705017,
  0.7612624764442444,
  0.580270528793335,
  0.44284871220588684,
  0.3270077109336853,
  0.2527984380722046,
  0.343576043844223,
  0.28853705525398254,
  0.398294597864151,
  0.5637757778167725,
  0.37727418541908264,
  0.5863168835639954,
  0.5760128498077393,
  0.29530686140060425,
  0.8156559467315674,
  0.2886416018009186,
  0.23251937329769135,
  0.5445767045021057
 ]
}


### S11. results/post_analysis.json


{
 "wilcoxon_svm_cnn": {
  "stat": 5.0,
  "p": 0.625,
  "diff": [
   -0.006936800587048264,
   0.13969731189318502,
   0.010342317261461953,
   0.025214787961324592,
   -0.02918323242209886
  ]
 },
 "diff_SVM_LogReg": [
  0.04657095135093625,
  -0.04745751814090193,
  -0.015434451549362627,
  0.03771380892782639,
  0.010439256438056876
 ],
 "diff_SVM_RF": [
  0.007877575400313419,
  0.03307392214365823,
  0.006479827051206977,
  0.033090609535632254,
  0.014391593958157367
 ],

"diff_SVM_PLS-DA": [
 0.05218145770708538,
 0.0865354441353855,
 0.07067943456452341,
 0.0640486067051883,
 0.009194124899612022
],
"auc_diff_ci": {
 "LogReg": [
  0.008383220340113792,
  -0.02176693910196684,
  0.04117612126894838
 ],
 "RF": [
  0.019873466834411737,
  -0.008281944045110077,
  0.04919041863374143
 ],
 "PLS-DA": [
  0.03569031652034435,
  0.002891107751788733,
  0.07028657201994898
 ],
 "1D-CNN": [
  0.029197334828816892,
  -0.005566648708370278,
  0.0635584659952349
 ]
},
"pooled_auc": {
 "SVM": 0.8434518262104469,
 "LogReg": 0.8342098255891358,
 "RF": 0.8229596591665557,
 "PLS-DA": 0.8071384192073846,
 "1D-CNN": 0.8134957617716239
},
"occlusion_perfold": {
 "top5": [
  [
   0.0,
   -0.005177514792899518
  ],
  [
   -0.0076577569578397675,
   -0.0011661807580175543
  ],
  [
   -0.010442578426878901,
   -0.002523128679562614
  ],
  [
   0.010720557396635133,
   -0.006036217303823088
  ],
  [
   0.0046532179805882645,
   -0.008022774327122173
  ]
 ],
 "contig5": [
  [
   0.020173430189940778,
   0.002218934911242698
  ],
  [
   0.0,
   -0.0011661807580174433
  ],
  [
   -0.010442578426878901,
   -0.001682085786374965
  ],
  [
   0.008076787802325014,
   0.012072434607645732
  ],
  [
   -0.01966110231588103,
   0.000258799171842683
  ]
 ],
 "top5_bands": [
  [

```
   908.1,
   988.627,
   1031.987,
   1038.181,
   1050.57
  ],
  [
   908.1,
   926.683,
   1031.987,
   1044.376,
   1050.57
  ],
  [
   994.821,
   1031.987,
   1038.181,
   1050.57,
   1570.896
  ],
  [
   926.683,
   988.627,
   1031.987,
   1050.57,
   1106.319
  ],
  [
   908.1,
   1031.987,
   1044.376,
   1050.57,
   1570.896
  ]
 ],
 "top10": [
  [
   0.0026842844674435096,
   0.0
  ],
  [
   -0.0076577569578397675,
   -0.0037900874635568016
  ],
  [
   -0.025378303834549865,
   -0.005606952621250438
  ],
  [
   -0.03771380892782639,
   -0.021462105969148215
  ],
  [
   -0.009762484875607047,
   -0.024844720496894457
  ]
 ],
 "contig10": [
  [
   0.0,
   -0.0017258382642998393
  ],
  [
   -0.010966493325786586,
   -0.008163265306122436
  ],
  [
   -0.01794224838800107,
   -0.015419119708438345
  ],
  [
   0.0,
   0.008048289738430525
  ],
  [
   -0.014341562015445009,
   0.0015527950310558758
  ]
 ],
 "top10_bands": [
  [
   908.1,
   926.683,
```

939.072,
988.627,
1031.987,
1038.181,
1050.57,
1570.896,
1583.285,
1589.479
],
[
908.1,
926.683,
994.821,
1031.987,
1044.376,
1050.57,
1106.319,
1570.896,
1583.285,
1589.479
],
[
908.1,
926.683,
988.627,
994.821,
1031.987,
1038.181,
1050.57,
1106.319,
1570.896,
1676.2
],
[
926.683,
939.072,
988.627,
994.821,
1031.987,
1038.181,
1050.57,
1100.125,
1106.319,
1570.896
],
[
908.1,
988.627,
1025.793,
1031.987,
1038.181,
1044.376,
1050.57,
1570.896,
1577.09,
1589.479
]
],
"top20": [
[
-0.010366826156299913,
-0.002712031558185446
],
[
-0.015223388207681543,
-0.019825072886297423
],
[
0.0,
-0.008410428931875491
],
[
-0.01087022762356371,
-0.027498323272971192
],
[
-0.009762484875607047,
-0.02329192546583858
]
],
"contig20": [
[
-0.01480486125505398,

-0.014053254437869755
],
[
-0.02607880958941866,
-0.021282798833819117
],
[
-0.04320684342644343,
-0.01878329128118872
],
[
-0.01087022762356371,
-0.009054325955734521
],
[
-0.0437494129801822,
-0.020962732919254656
]
],
"top20_bands": [
[
908.1,
914.294,
926.683,
939.072,
951.46,
988.627,
1031.987,
1038.181,
1044.376,
1050.57,
1100.125,
1106.319,
1155.874,
1162.069,
1471.786,
1570.896,
1577.09,
1583.285,
1589.479,
1595.673
],
[
908.1,
926.683,
932.877,
982.432,
988.627,
994.821,
1031.987,
1038.181,
1044.376,
1050.57,
1081.542,
1106.319,
1124.902,
1143.485,
1155.874,
1471.786,
1570.896,
1577.09,
1583.285,
1589.479
],
[
908.1,
926.683,
932.877,
982.432,
988.627,
994.821,
1031.987,
1038.181,
1044.376,
1050.57,
1106.319,
1143.485,
1155.874,
1360.288,
1422.231,
1471.786,
1564.702,
1570.896,

```
    1577.09,
    1676.2
   ],
   [
    908.1,
    926.683,
    939.072,
    982.432,
    988.627,
    994.821,
    1025.793,
    1031.987,
    1038.181,
    1044.376,
    1050.57,
    1081.542,
    1100.125,
    1106.319,
    1124.902,
    1143.485,
    1155.874,
    1471.786,
    1570.896,
    1589.479
   ],
   [
    908.1,
    988.627,
    1025.793,
    1031.987,
    1038.181,
    1044.376,
    1050.57,
    1106.319,
    1124.902,
    1131.097,
    1143.485,
    1416.037,
    1422.231,
    1428.426,
    1558.507,
    1570.896,
    1577.09,
    1583.285,
    1589.479,
    1676.2
   ]
  ]
 },
 "occlusion_sd": {
  "top5": [
   0.0077818333110056224,
   0.0024582528749177244
  ],
  "contig5": [
   0.013907703695979184,
   0.005050481440262067
  ],
  "top10": [
   0.014255530330757186,
   0.010030958679359948
  ],
  "contig10": [
   0.0073993364511951805,
   0.008064861695621072
  ],
  "top20": [
   0.0050075702501712685,
   0.009312169614514077
  ],
  "contig20": [
   0.013785435189995495,
   0.004666768291904774
  ]
 }
}
```
